# Clustering high-cost patients in England using machine learning: a population-based cohort study

**DOI:** 10.64898/2025.12.23.25342912

**Authors:** Shaolin Wang, Laura Anselmi, Matt Sutton, Evangelos Kontopantelis, Thomas Beaney, Michael Anderson

## Abstract

**Objective:** To identify clusters of high-cost patients in England based on diagnoses and sociodemographic characteristics to inform targeted population health management.

**Design:** A retrospective population-based cohort study using unsupervised machine learning.

**Setting:** English primary care electronic health records from the Clinical Practice Research Datalink, linked to Hospital Episode Statistics for hospital records and Office for National Statistics mortality data.

**Participants:** 10,119,490 adult patients aged 18 years or over registered with 1,397 general practices in England on 1 April 2018. High-cost patients were defined as the top 1% of total healthcare spending (n=101,195). Additional high-cost population were examined, including age-specific subgroups, patients who died during the year and patients in the top 1% of unplanned care costs.

**Main outcome measures:** Primary and secondary care costs in financial year 2018/19. Clusters of high-cost patients defined using unsupervised machine learning based on age, sex, area-level deprivation, ethnicity, and diagnoses recorded during 2006/07-2018/19.

**Results:** High-cost patients accounted for GBP1.8billion (26.8%) of GBP6.6billion population costs. Mean annual costs per high-cost patients were GBP17,485 (median GBP14,609; interquartile range: GBP12,028 to GBP19,633) compared with GBP653 (GBP103; GBP14 to GBP352) in the overall population. Hierarchical clustering identifying nine clusters was the optimal solution based on evaluation combining multiple validity and stability metrics. Across those clusters, mean age ranged from 56 to 79 years, and mean annual costs ranged from GBP15,792 (95%CI GBP15,629 to GBP15,955) to GBP19,107 (GBP18,784 to GBP19,430). Notable clusters produced across clustering approaches and high-cost populations, including younger people with liver disease and mental health conditions, patients with nodal metastases, patients with prostate cancer and hyperplasia, and older people with cardiovascular disease and dementia.

**Conclusion:** High-cost patients are a heterogeneous population with distinct clinical and sociodemographic profiles and utilization patterns. Clustering across multiple high-cost populations identified recurrent clusters, highlighting common pathways of high expenditure, while also revealing population-specific patterns of need. Incorporating cluster-based approaches into population health management may improve the targeting of case management programmes, optimise resource allocation, and support more effective and sustainable health system planning.

**What is already known on this topic:**

- A small proportion of patients account for a large share of healthcare costs, and are a priority for population health management.
- Previous clustering studies show heterogeneity among high-cost patients, but are often limited by scale, care settings, or lack of robustness assessment

**What this study adds:**

- Using linked English primary and secondary care data for over 10 million adults, the top 1% high-cost patients accounted for more than a quarter of total costs.
- By comparing multiple clustering methods across several high-cost populations, we identify recurrent, clinically interpretable subgroups, including younger adults with liver disease and mental health conditions, highly deprived, with heavy emergency use; oncology with nodal metastases, intensive planned pathways and high mortality; older men with prostate cancer or hyperplasia, sustained planned care; and older adults with cardiovascular disease and dementia, recurrent emergency admissions and high primary-care contact

**How this study might affect research, practice or policy:**

- Robust segmentation can complement risk prediction by supporting more tailored, multidisciplinary care for high-cost patients.
- Cluster profiles can inform population health management and service planning in universal healthcare systems.

## Introduction

Globally, there has been increased attention on characterising high-cost patients, reflecting concerns about growing healthcare budgets, and the disproportionate share of healthcare spending concentrated in a small number of patients. ^1–3^ Segmentation of patients into clinically distinct groups can help build an understanding of which specific patient groups are associated with high-costs, which in turn can inform targeted interventions and service design, enabling more efficient use of resources and better patient outcomes.^4,5^ This is a key strategy for population health management, which is understood as using a data-driven approach to identify at-risk groups and design targeted and personalized interventions.^6^

A systematic review, published in 2018, found 55 articles on characteristics of high-cost patients.^7^ Most studies were from the US (38/55), with other studies from Canada, the Netherland, Denmark, Germany, and Taiwan. They found a high prevalence of mental illness in high-cost patients, and while age was a strong predictor of being high-cost, half of high-cost patients were younger than 65. Most studies used descriptive or regression analyses, with only three studies employing clustering approaches. Subsequent research has increasingly used data-driven segmentation to move beyond descriptive characterisation of high-cost populations, applying a range of approaches to clustering including latent class analysis,^8–10^ k-means,^8,11^ hierarchical,^11,12^ and density-based.^11,13,14^ Most analyse single care settings at regional or institutional level, with two notable exceptions use linked primary and secondary care data. Wick et al. (2022) used linked inpatient, emergency, outpatient physician and pharmacy data for 3·9 million adults in Alberta to identify 21,115 persistently high-cost users between 2014 and 2019, defined as top 1% of healthcare spenders in at least two consecutive fiscal years.^9^ Trenaman et al. (2025) analysed 224,285 high-need, high-cost patients (top 5% of total health-system costs in 2017) within a cohort 5·4 million British Columbians using linked records for hospital, day surgery, physician services, prescription drugs, and home and community care.^15^ Common clusters recurred across studies, including those characterised by mental health and substance misuse, cancer, neurological conditions, cardiometabolic and renal disease, complex multimorbidity and frailty, and advanced cardiovascular disease.

Examining high-cost patients in England has important advantages, including the use of comprehensive linked, nationally representative datasets and a public health system where access is based on clinical need rather than ability to pay, meaning there are no insurance-related barriers to service use. Despite this, there is limited evidence on the characteristics of high-cost patients from England. An analysis from the Health Foundation using data from the financial year 2014/15 discovered that 5% of patients in England accounted for approximately half of the healthcare costs in primary and secondary care settings.^1^ The high-cost group of patients were older and suffered from a higher-level morbidity, with 61·4% of patients above 60 years old, and 55·9% having more than three conditions. The most common pre-existing diagnoses in the high-cost group included cancer, heart failure, Parkinson’s disease, atrial fibrillation, peripheral vascular disease, and bronchiectasis. Soley-Bori et al 2023 clustered high-cost individuals with multiple long term conditions (MLTCs), classified as within the top 20% of primary care costs in on borough of London (Lambeth).^10^ They identified five common disease combinations, with the trio of anxiety, chronic pain, and depression representing the highest share of costs.

We aimed to focus on total primary and secondary care costs to define and cluster high-cost patients in the English National Health Service (NHS). Our paper contributes to the literature on high-cost patients by using a large nationally representative dataset, testing multiple clustering algorithms, and using a broad range of information on patient diagnoses and sociodemographic characteristics. In doing so, we hope to establish whether high-cost patients can be categorised into distinct clusters that could be targeted with case-management programmes to improve quality and coordination of care and help manage overall healthcare costs.

## Methods

### Study design and participants

We used data from the Clinical Practice Research Datalink (CPRD) Aurum,^16^ linked with Hospital Episode Statistics (HES),^17^ Office for National Statistics (ONS) death registration,^18^ and 2019 English Index of Multiple Deprivation (IMD) decile.^19^ These linkages have been used in prior research on diagnostic coding^20^ and healthcare costs.^21,22^

CPRD covers a broadly nationally representative sample of 20 percent of the general practice population in England, and contains pseudonymised data on age, sex, ethnicity, diagnoses, test results, referrals, prescriptions, general practice registration and general practice ID.^16^ Data were processed and retained for 10,119,490 adult patients aged 18 years or older registered with 1,397 general practices in England on 1 April 2018, following established cleaning procedures described in prior work.^22^ The study protocol was approved by the CPRD Independent Scientific Advisory Panel (protocol 21_000693) and a summary is available online.

### Patient characteristics

Information on patient characteristics was used as of 1^st^ of April 2018. Binary indicators were generated for sex(self-reported at registration), four age bands (18-44, 45-65, 66-79 and 80+), five deprivation quintiles of the patient’s area of residence based on the 2019 English IMD, and five ethnic categories (White, Mixed, Asian, Black, Other), with ethnicity derived from the standard CPRD-HES linkage approach described previously. ^20^ Patients with unknown ethnicity or unknown deprivation were included in separate unknown categories.

We identified diagnoses using the CALIBER phenotyping algorithms, ^23,24^ which defines physical and mental health conditions based on ICD-10 codes in HES and Read codes in the CPRD GOLD. We ascertained 209 LTCs mapped to CPRD Aurum^25^ using records between 2006/07 and 2018/19. The observation period began in 2006/07, two years after the launch of the Quality and Outcomes Framework programme in 2004 and its first major revision, to ensure more stable and consistent diagnostic coding.^20^ To capture conditions most directly associated with costs in the study year, we additionally mapped 91 acute conditions to CPRD Aurum and identified diagnoses recorded in 2018/19 (code lists available on a github data repository).

### Healthcare costs

Following the approach in previous research on healthcare utilisation costing, ^21^ ^26^ we estimated primary and secondary care costs for each registered patient in the subsequent financial year (2018/19), which was the most recent complete year unaffected by the COVID-19 pandemic. Primary care activity was deduplicated (same date, staff role and appointment mode), and costed by staff role and appointment mode using 2018/19 Personal Social Services Research Unit (PSSRU) unit costs,^27^ calculated as the product of the hourly contact cost and the mean length of appointment. Where a direct unit cost was unavailable, we scaled to the relevant salary band and used the latest length estimates from PSSRU or UK General Practice Workload Survey.^28^ Secondary care costs were estimated by assigning 2018/19 National Tariff prices to each inpatient stay, outpatient attendance, and emergence department (ED) attendance, supplemented by NHS Reference Costs or specialty average where unavailable.^21^ To remove outliers and likely also specialised high-cost cases, total annual costs were truncated at £100,000 per patient-year, consistent with the NHS General and Acute funding formula.^29,30^

Total care costs were partitioned into planned care (scheduled GP appointments, elective admissions, surgery, or hospital transfers, and outpatient care)^31^ and unplanned care (emergency admissions and emergency department (ED) attendances). We also calculated contact counts by service: emergency admissions, ED attendances, elective admissions, outpatient attendances, and GP appointments.

### High-cost patients

We examined clustering among top 1% high-cost patients defined by total healthcare costs across primary and secondary care from 1 April 2018 to 31 March 2019, as the main analysis. We additionally assessed five subgroups: four age groups (18-44, 45-65, 66-79, and 80+ years), and patients who died in 2018/19. Finally, we examined clustering among high-cost patients based specifically on unplanned care costs.

### Statistical analysis

Figure 1 summaries the analytical pipeline from data processing to cluster profiling: (i) dimension reduction of high-dimensional diagnostic and sociodemographic variables; (ii), clustering using alternative algorithms; (iii) selection of optimal clusters based on validity and stability metrics; and (iv) cluster profiling to characterise sociodemographic, diagnostic, and utilisation patterns. Hierarchical and k-medoids clustering require computation of an all-pairs dissimilarity matrix, which scales quadratically with sample size. For approximately 100,000 high-cost patients, this entails more than10 billion pairwise distances, exceeding the memory capacity of the “cluster” and “fclust” packages in R even on high-performance computing. To ensure computational feasibility, we drew a 50% random sample for the main analysis and five subgroup analyses (i.e. aged 18–44 years, aged 45-65 years, aged 66–79 years, aged 80 years and over, and in-year descents). A second independent 50% random sample of total-cost high-cost patients was drawn to assess robustness for our main analysis. A third independent 50% random sample was drawn focusing exclusively on unplanned costs. Full methodological details, including dimension reduction, clustering algorithms, cluster evaluation and selection, are provided in Appendix text S1-3.

**Figure 1:**
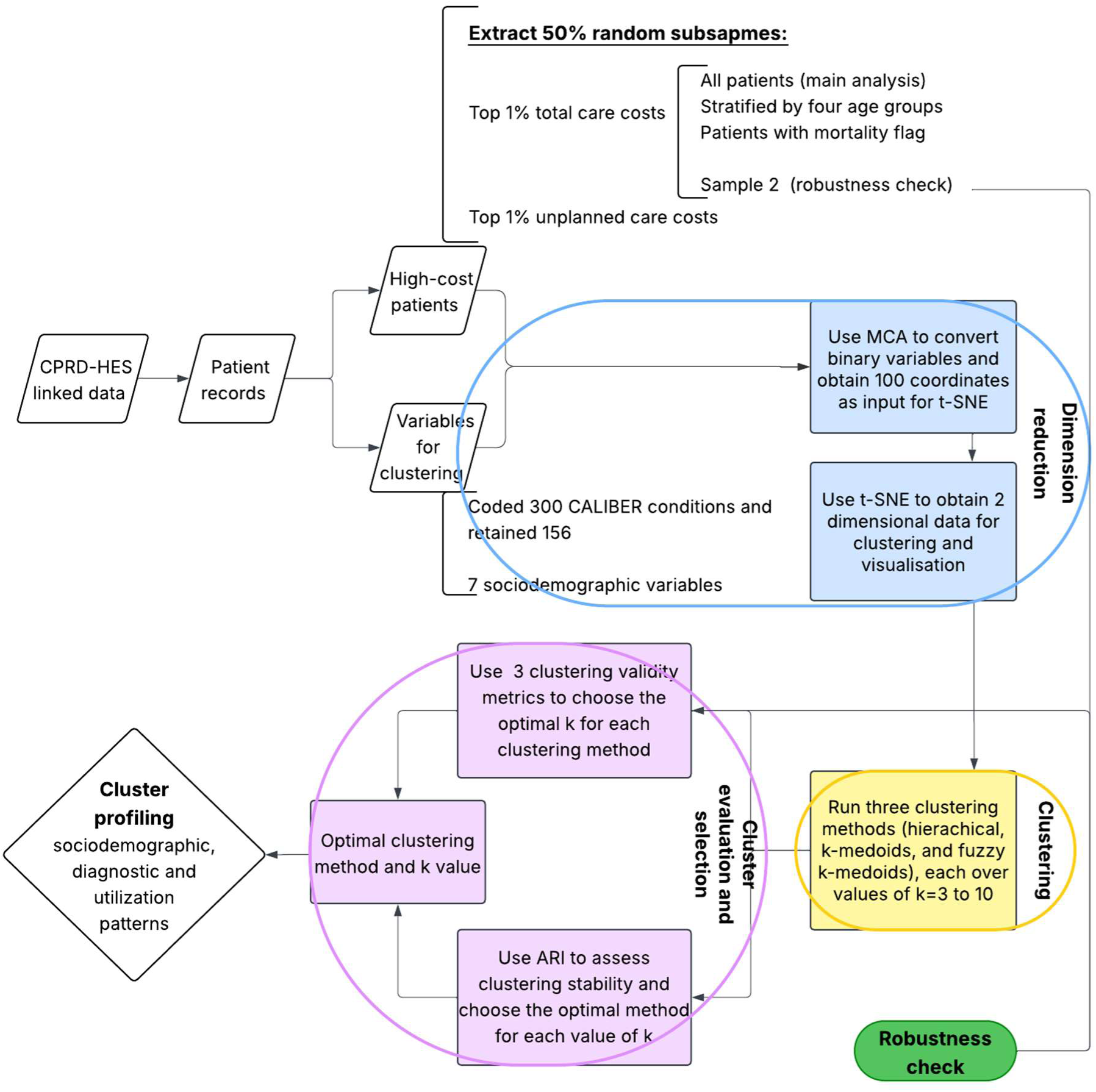
Steps of our methodology from data processing to clustering.

Because diagnostic data are high-dimensional and correlated, we first reduced dimensionality to improve computational feasibility and cluster interpretability. We excluded diagnoses with prevalence below 1%^11^ and four common blood test abnormalities (low HDL-C, raised LDL-C, raised total cholesterol, raised triglycerides). Binary diagnostic indicators were then mapped to a continuous latent space using multiple correspondence analysis (MCA), retaining components explaining 90% of variation. We further applied t-distributed stochastic neighbour embedding (t-SNE) following Yan et al (2019)^11^ to project the MCA embedding to two dimensions, enabling visualisation of complex data patterns and exploration of potential clustering structure.

We applied three unsupervised clustering algorithms to the reduced data. First, agglomerative hierarchical clustering with Ward’s criterion^32,33^ starts with each patient as a single cluster and iteratively merges the most similar clusters, producing a dendrogram that can be cut at different levels to yield a desired number of clusters. Second, k-medoids partitions patients into a prespecified number of groups, k, by selecting representative patients as medoid for each cluster and then assigning others to their nearest medoid^34^; this approach is robust to outliers and irregular cluster shapes. Third, fuzzy k-medoids extends k-medoids by assigning partial membership (weights) to multiple clusters (via membership weights)^35^, accommodating the clinical reality that patient profiles can overlap. See a detailed description of three clustering algorithms in Appendix Table S1.

Each method was run over k = 3-10 (pre-specified for interpretability). We assessed clustering validity using the Silhouette, Davies–Bouldin, and Calinski–Harabasz indices^36^, and stability using the Adjusted Rand Index (ARI). Validity and stability metrics were standardised and combined into a composite score to select the optimal method-k solution.

Clusters were profiled using within-cluster statistics of sociodemographic characteristics, in-year mortality, healthcare utilization (total costs, planned vs. unplanned costs, and contact counts by service), key diagnoses and multimorbidity (number of CALIBER diagnoses in total, and for LTCs and chronic conditions separately). Key diagnoses were defined by relative prevalence (O/E) ≥ 2^37^, or otherwise the highest relative prevalence across clusters to capture distinctive but moderate enrichments. For visualisation, radar charts summarised disease patterns by system, restricted to approximately 80 conditions highly relevant to the first two MCA coordinates for readability. Full descriptive tables of patient characteristics, utilization patterns and the top 20 diseases by relative prevalence are provided in Supplement.

We assessed robustness of high-cost clusters produced from the main analysis in two ways: (i) cross-method comparison of the optimal solutions from hierarchical, k-medoids and fuzzy k-medoids; and (ii) cross-sample comparison using two independently drawn 50% samples for our main analysis. Clusters were aligned by disease profile similarity and classified as matched (same profile; label retained), reproduced (core profile preserved with additional or shifted features; label refined), or dropped (no standalone recover; features dispersed into one or more other clusters, including any newly formed classes). Analyses were undertaken using R (version 4.4.2).

### Patient and public involvement

Patient representatives were involved in the design and interpretation of this research through consultation with members of the Primary Care Research in Manchester Engagement Resource (PRIMER) group, University of Manchester. Findings will be communicated to the public through multiple media channels.

## Results

In total, the top 1% high-cost patients accounted for £1.8M (26·8%) of £6.6M overall primary and secondary care costs (Table 1; Appendix Figure S1). Median costs for the 1% group were £14,609 (Interquartile range (IQR): £12,028 to 19,633), compared to £103 (IQR: £ 14 to 352) in the full study population. Costs were predominantly driven by secondary care (median £14,077, IQR: £11,561 to 19,055, vs £0, IQR: £0 to 144), and by unplanned care (median £10,872, IQR: £5,462 to 14,924, vs £0, IQR: £0 to 0).

**Table 1:** Descriptive Statistics for full study population and top 1% high-cost patients.

|  | Full study population<br>(N=10,119,490) |  | Top 1% high-cost patients<br>(N=101,195) |  |
| --- | --- | --- | --- | --- |
| **Utilization in 2018/19** |  |  |  |  |
| **per person** | <u>Mean (s.d.)</u> | <u>Median (IQR)</u> | <u>Mean (s.d.)</u> | <u>Median (IQR)</u> |
| Total care costs (£) | 653 (2,248) | 103 (14-352) | 17,485 (8,921) | 14,609 (12,028-19,633) |
| Secondary care costs (£) | 530 (2,180) | 0 (0-144) | 16,941 (8,915) | 14,077 (11,561-19,055) |
| Primary care costs (£) | 123 (187) | 68 (0-165) | 544 (529) | 406 (202-721) |
| Planned care costs (£) | 373 (1270) | 82 (0-261) | 6,199 (7,949) | 3,039 (831-10,299) |
| Unplanned care costs (£) | 279 (1592) | 0(0-0) | 11,288 (8,587) | 10,872 (5,462-14,924) |
| N(emergency admissions) | 0.1 (0.5) | 0 (0-0) | 2.8 (2.7) | 2 (1-4) |
| N(ED attendances) | 0.4 (1.4) | 0 (0-0) | 4.0 (7.4) | 2 (1-5) |
| N(elective admissions) | 0.2 (2.1) | 0 (0-0) | 3.6 (11.7) | 1 (0-3) |
| N(outpatient attendances) | 1.7 (4.4) | 0 (0-2) | 13.0 (16.1) | 8 (3-17) |
| N(GP appointments) | 4.8 (6.7) | 3 (0-7) | 17.4 (16.4) | 14 (7-23) |
| **Multimorbidity** |  | <u>Mean (s.d.)</u> |  | <u>Mean (s.d.)</u> |
| N(CALIBER conditions) |  | 4.8 (5.0) |  | 19.4 (8.2) |
| N(long term conditions) |  | 4.6 (4.7) |  | 15.6 (7.0) |
| N(acute conditions) |  | 0.2 (0.8) |  | 3.7 (2.7) |
| N(disease systems) |  | 2.8 (2.4) |  | 8.0 (2.4) |
| **Socio-demographics** |  |  |  |  |
| **Age** |  | 47.7 (18.8) |  | 70.3 (16.7) |
| **Age group** |  | <u>Number (%)</u> |  | <u>Number (%)</u> |
| 18-44 |  | 4,767,330 (47.1%) |  | 8,653 (8.6%) |
| 45-65 |  | 3,327,290 (32.9%) |  | 24,284 (24.0%) |
| 66-79 |  | 1,414,167 (14.0%) |  | 33,601 (33.2%) |
| 80+ |  | 610,703 (6.0%) |  | 34,657 (34.2%) |
| **Sex** |  |  |  |  |
| Male |  | 5,046,041 (49.9%) |  | 52,099 (51.5%) |
| Female |  | 5,073,449 (50.1%) |  | 49,096 (48.5%) |
| **Ethnicity** |  |  |  |  |
| White |  | 7,524,093 (74.4%) |  | 89,933 (88.9%) |
| Mixed |  | 84,272 (0.8%) |  | 272 (0.3%) |
| Asian |  | 679,652 (6.7%) |  | 4,324 (4.3%) |
| Black |  | 395,812 (3.9%) |  | 3,200 (3.2%) |
| Other |  | 225,871 (2.2%) |  | 690 (0.7%) |
| Unknown |  | 1,209,790 (12.0%) |  | 2,776 (2.7%) |
| **IMD quintile** |  |  |  |  |
| 1 (least deprived) |  | 2,042,741 (20.2%) |  | 19,282 (19.1%) |
| 2 |  | 2,061,284 (20.4%) |  | 20,017 (19.8%) |
| 3 |  | 2,007,416 (19.9%) |  | 19,742 (19.5%) |
| 4 |  | 2,110,361 (20.9%) |  | 20,566 (20.3%) |
| 5 (most deprived) |  | 1,886,234 (18.7%) |  | 21,495 (21.3%) |
| unknown |  | 11,454 (0.1%) |  | 93 (0.1%) |
| **In-year mortality flag** |  | 91,166 (0.9%) |  | 20,335 (20.1%) |
| **New registration** |  | 848,117 (8.4%) |  | 9,086 (9.0%) |
| **In-year deregistration** |  | 683,342 (6.8%) |  | 11,315 (11.2%) |

As expected, utilisation was markedly higher for this population both in primary (medians: GP appointments 14 (IQR 7 to 23) vs 3 (IQR 0 to 7)) and secondary care settings (emergency admissions 2 (IQR 1 to 5) vs 0 (IQR 0 to 0); elective admissions 1 (IQR 0 to 3) vs 0 (IQR 0 to 0); outpatients 8 (IQR 3 to 17) vs 0 (IQR 0 to 0); ED attendances 2 (IQR 1 to 4) vs 0 (IQR 0 to 0)). High-cost patients were older (mean 70·3 years vs 47·7 years), had greater multimorbidity (mean 15·6 vs 4·6 long-term conditions; mean 3·7 vs 0·2 acute conditions), and much higher in-year mortality (20,335 [20·1%] of /101,195 vs 91,166 [0·9%] of 10,119,490). They were more likely to be White (89,933 [88·9%] vs 7,524,093[74·4%]) and to live in the most deprived quintile (21,495[21·3%] vs 1,886,234[18·7%]). Sex distribution was similar.

### t-SNE projection

Based on the combined validity-stability score, the hierarchical solution with k=9 ranked best overall, followed by k-medoids (k=6) and fuzzy k-medoids (k=7). Detailed results on dimension reduction and cluster evaluation are provided in Appendix text S4 and Tables S2-3. Figure 2 shows the three clustering solutions on the 2D t-SNE map (each point represents a patient; distances approximate similarity; colours denote cluster assignment). The clustering methods yielded distinct partitions, although the main body of patients was consistently split into a few dominant clusters; differences arose primarily in how edge-case profiles were handled. The hierarchical solution (k=9), identified as optimal overall, provides the most granular segmentation, dividing the space into nine groups and isolating several small peripheral clusters alongside the central ones. This finer resolution is consistent with its stronger CHI and DBI performance.

**Figure 2.**
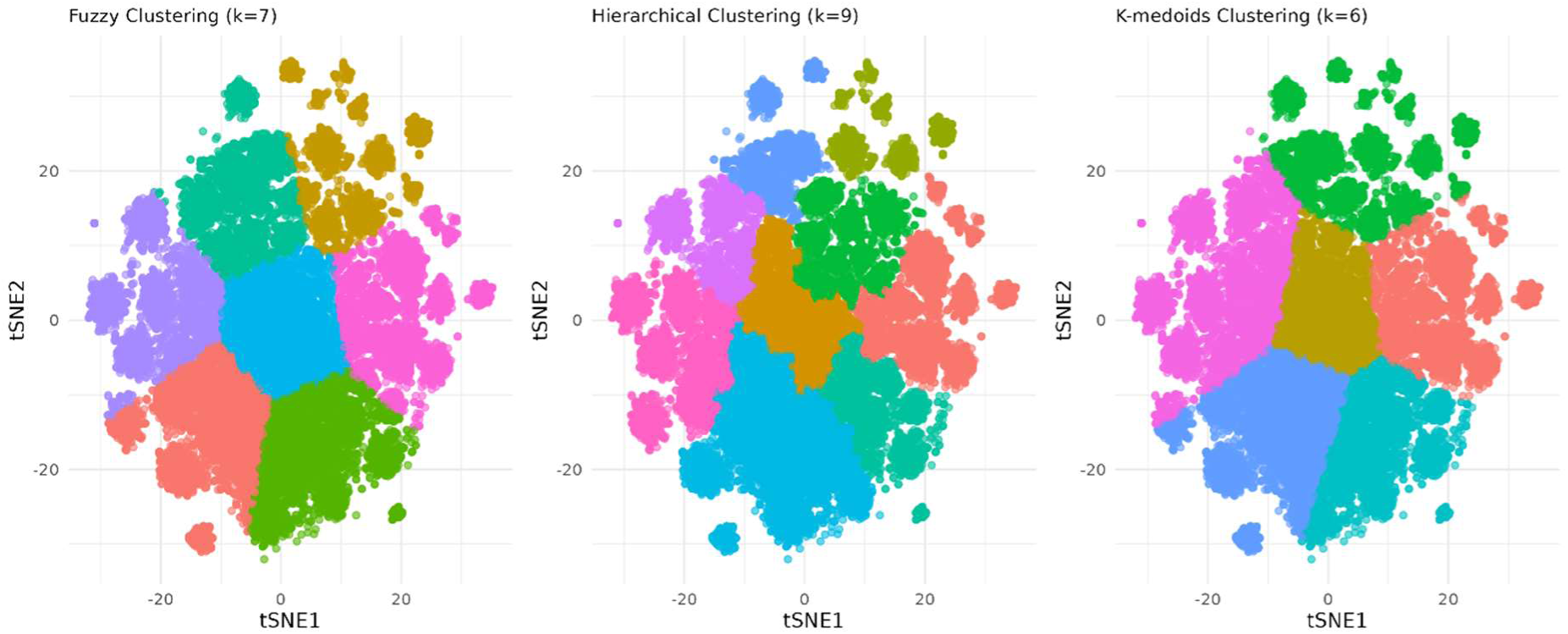
Cluster configurations for hierarchical, k-medoids, and fuzzy k-medoids clustering

### Cluster profiles: main analysis

We characterised nine clusters from the optimal hierarchical solution among those who were high-cost patients in the subsequent year, using socio-demographic, diagnostic, and utilization patterns (Table 2; full statistics in Appendix Table S4). Clusters are ordered by increasing mean age. A radar chart (Appendix Figure S2) displays O/E prevalence ratios, colour-coded by disease system, and provides a visual cross-check of distinct cluster signatures.

**Table 2:** Cluster profiles of high-cost patients, 2018/19, based on 50% random sample (N=52,175; optimal hierarchical 9-cluster solution)

| Cluster | Mean age | Demographics | Feature diagnoses (O/E prevalence ratio) | Service use | Mortality | Profile summary |
| --- | --- | --- | --- | --- | --- | --- |
| 1. Liver disease & mental health (N=6,523 [12.5%]) | 55.8 | Younger, more deprived (mean age: 55.8; IMD=4-5: 3,319 [50.9%]) | alcoholic liver disease (6.8), portal hypertension (6.4), personality disorders (5.8), acne (5.7), cirrhosis (5.5), bipolar (5.2), substance misuse (3.9), schizophrenia (3.9), hypersplenism (3.7), pancreatitis (3.6), fatty liver (2.8), alcohol misuse (2.7), epilepsy (2.3), cholelithiasis (2.2), secondary thrombocytopaenia (1.8), depression (1.7), anxiety disorders (1.7), oesophagitis and oesophageal ulcer (1.7), irritable bowel syndrome (1.6) | Very high unplanned care (£12,476; 3.8 emergency admissions; 7.3 ED attendances) | 882 (13.5%) | Young adults with alcohol-related liver disease and severe mental health/substance misuse; heavy emergency care use. |
| 2. Sleep apnoea, gastro-reflux and diabetes (N=4,589 [8.8%]) | 65.6 | Mid-older, more deprived (IMD=4-5: 2,442 [53.2%]), higher minority ethnicity (1,926 [42.0%]) | sleep apnoea (2.8), peptic ulcer (2.7), diabetic eye disease (1.7), diabetes (1.4), unstable angina (1.4), respiratory failure (1.3) | High secondary care (£17,391) | 704 (15.3%) | Ethnically diverse patients with sleep apnoea, peptic ulcer disease, and diabetes; high secondary care use. |
| 3. Chronic fatigue, immunological and haematological disorders (N=5,464 [10.5%]) | 65.6 | Ethnicity under-recorded (1,129 [20.7%] unknown) | chronic fatigue syndrome (3.8), hypersplenism (2.1), secondary thrombocytopaenia (1.9), infections of digestive system (1.3) | Highest total and planned care (£19,107 and £8,684); frequent elective admissions (5.9) | 929 (17.0%) | Patients with chronic fatigue, immunological, and haematological conditions; highest total and planned care costs. |
| 4. Low multimorbidity (N=6,852 [13.1%]) | 66.2 | Least deprived (IMD=4-5: 4,460 [35.9%]) | Highest O/E prevalence was 1.1. | High planned care; balanced planned/unplanned (means £7,982 vs. £7,810) | 715 (10.4%) | Patients in the least deprived areas with relatively low multimorbidity; costs are equally distributed between planned and unplanned care |
| 5. Nodal metastases (N=3,514 [6.7%]) | 67.3 | Predominantly female (2,371 [67.5%]), less deprived (IMD=4-5: 36.4%) | secondary malignancy in lymph nodes (5.4), primary malignancy skin (1.4) | Most frequent elective admissions (6.7) and outpatient (21.5) | 1,128 (32.1%) | Mostly female patients with advanced cancers with lymph node metastases; frequent elective care and highest mortality. |
| 6. Complex multimorbidity | 68.3 | Predominantly female (1,905 [67.0%]), most | skin cancer (1.5), spondylosis (1.5) | Most frequent GP appointments | 475 (16.7%) | Older, mainly female patients with complex multiple morbidity; high |
| (N=2,842<br>[5.4%]) |  | multimorbid (21<br>diagnoses) |  | (19.4); frequent<br>ED visits (4.8) |  | primary and emergency care<br>use. |
| 7. Prostate<br>cancer (with<br>nodal<br>metastasis)<br>(N=3,850<br>[7.4%]) | 72.8 | Predominantly<br>male (2,645<br>[68.7%]) | Prostate cancer (5.0), secondary malignancy in lymph<br>nodes (3.0), COPD (1.4), diverticular disease of intestine<br>(1.3) | Frequent elective<br>admissions (4.1),<br>outpatient (16.0) | 1,201<br>(31.2%) | Older men with prostate<br>cancer and hyperplasia;<br>intensive planned care use<br>and high mortality. |
| 8. Prostate,<br>stroke &<br>sensory<br>impairment<br>(N=6,138<br>[11.8%]) | 78.9 | Predominantly<br>male (3,892<br>[63.4%]) | prostate hyperplasia (1.8), stroke (1.7), macular<br>degeneration (1.7), visual impairment (1.6), transient<br>ischaemic attack (1.6), glaucoma (1.5), PAD (1.5),<br>diaphragmatic hernia (1.4) cataract (1.4), hearing loss<br>(1.4), urinary tract infections (1.4), osteoporosis (1.4) | Frequent<br>emergency<br>admissions (3.0) | 1,568<br>(25.5%) | Oldest men with prostate<br>cancer, prostate hyperplasia,<br>and sensory impairment |
| 9.<br>Cardiovascular<br>disease and<br>dementia<br>(N=12,403<br>[23.8%]) | 79.2 | Eldest, mixed<br>sex (mean age:<br>79.2) | cardiomyopathy (2.5), rheumatic valve disorders (2.3),<br>pulmonary hypertension (2.1), left bundle branch block<br>(2.1), dementia (2.0), multiple valve disorder (1.9),<br>atrioventricular blocks (1.9), nonrheumatic mitral valve<br>disorders (1.9), diabetic neuropathy (1.9), heart failure<br>(1.7), macular degeneration (1.7), right bundle branch<br>blocks (1.7), Ischaemic stroke (1.7), delirium (1.7), AF<br>(1.6), nonrheumatic aortic valve disorders (1.6), Transient<br>ischaemic attack (1.6), myocardial infarction (1.5) | Highest<br>unplanned and<br>primary care<br>(£13,375 and<br>£606) | 3,157<br>(25.5%) | Elderly with complex<br>cardiovascular disease and<br>dementia; highest unplanned<br>and primary care demand. |

**Table 3:**
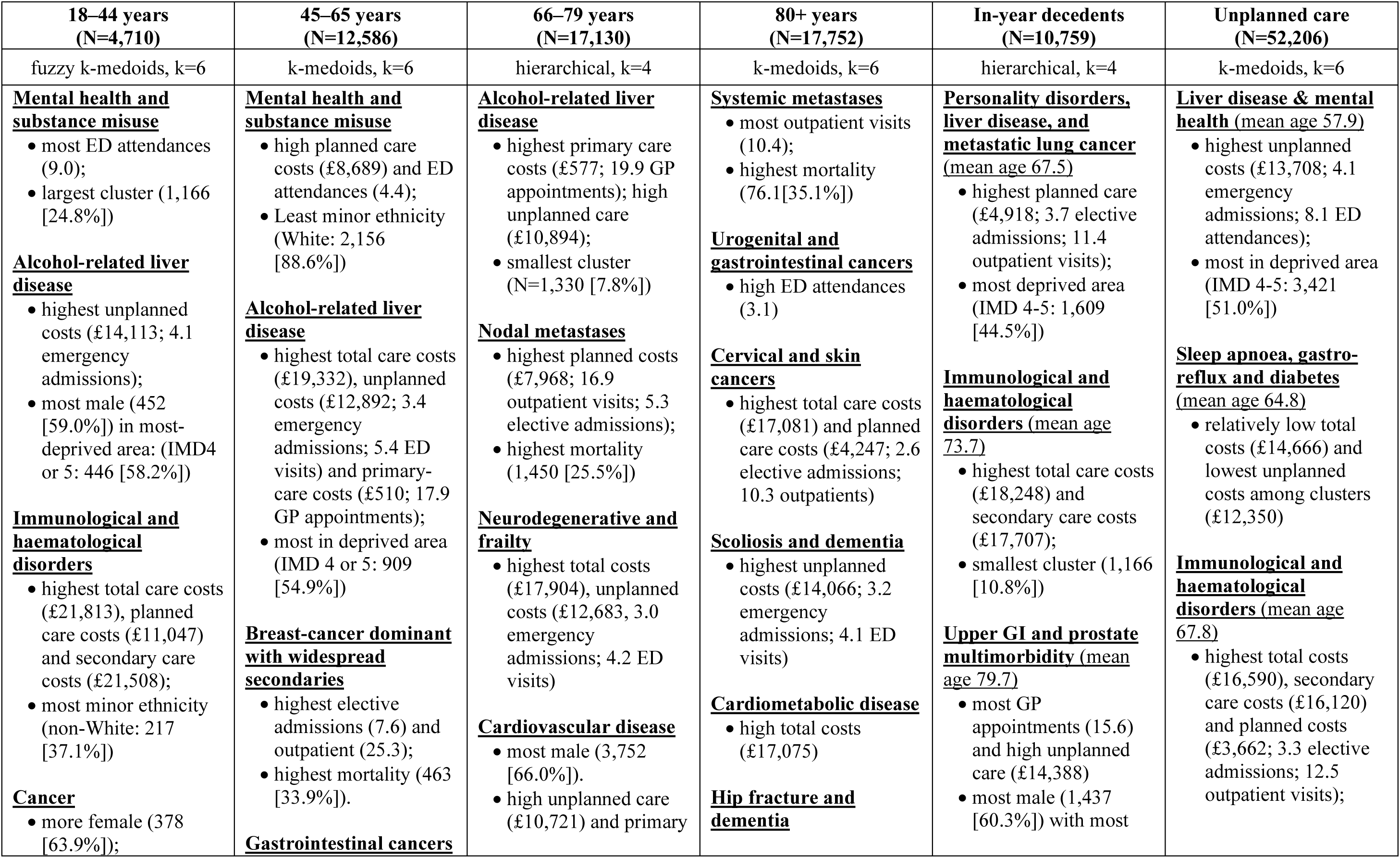
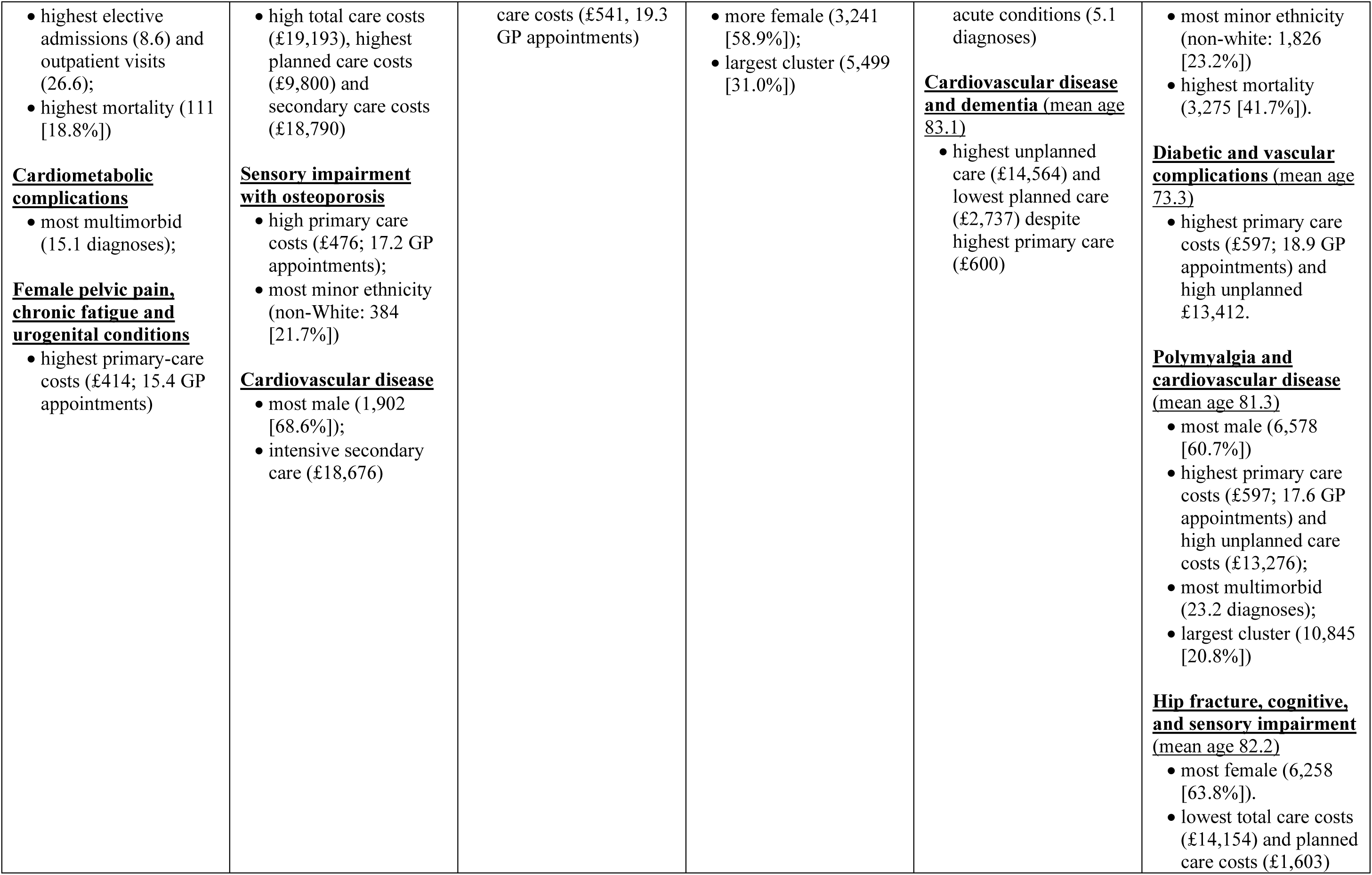
Cluster profiles of high-cost patients by age groups, 2018/19.

| **18–44 years**<br>**(N=4,710)** | **45–65 years**<br>**(N=12,586)** | **66–79 years**<br>**(N=17,130)** | **80+ years**<br>**(N=17,752)** | **In-year decedents**<br>**(N=10,759)** | **Unplanned care**<br>**(N=52,206)** |
| --- | --- | --- | --- | --- | --- |
| fuzzy k-medoids, k=6 | k-medoids, k=6 | hierarchical, k=4 | k-medoids, k=6 | hierarchical, k=4 | k-medoids, k=6 |
| **<u>Mental health and substance misuse</u>**<br>• most ED attendances (9.0);<br>• largest cluster (1,166 [24.8%])<br>**<u>Alcohol-related liver disease</u>**<br>• highest unplanned costs (£14,113; 4.1 emergency admissions);<br>• most male (452 [59.0%]) in most-deprived area: (IMD4 or 5: 446 [58.2%])<br>**<u>Immunological and haematological disorders</u>**<br>• highest total care costs (£21,813), planned care costs (£11,047) and secondary care costs (£21,508);<br>• most minor ethnicity (non-White: 217 [37.1%])<br>**<u>Cancer</u>**<br>• more female (378 [63.9%]); | **<u>Mental health and substance misuse</u>**<br>• high planned care costs (£8,689) and ED attendances (4.4);<br>• Least minor ethnicity (White: 2,156 [88.6%])<br>**<u>Alcohol-related liver disease</u>**<br>• highest total care costs (£19,332), unplanned costs (£12,892; 3.4 emergency admissions; 5.4 ED visits) and primary-care costs (£510; 17.9 GP appointments);<br>• most in deprived area (IMD 4 or 5: 909 [54.9%])<br>**<u>Breast-cancer dominant with widespread secondaries</u>**<br>• highest elective admissions (7.6) and outpatient (25.3);<br>• highest mortality (463 [33.9%]).<br>**<u>Gastrointestinal cancers</u>** | **<u>Alcohol-related liver disease</u>**<br>• highest primary care costs (£577; 19.9 GP appointments); high unplanned care (£10,894);<br>• smallest cluster (N=1,330 [7.8%])<br>**<u>Nodal metastases</u>**<br>• highest planned costs (£7,968; 16.9 outpatient visits; 5.3 elective admissions);<br>• highest mortality (1,450 [25.5%])<br>**<u>Neurodegenerative and frailty</u>**<br>• highest total costs (£17,904), unplanned costs (£12,683, 3.0 emergency admissions; 4.2 ED visits)<br>**<u>Cardiovascular disease</u>**<br>• most male (3,752 [66.0%]).<br>• high unplanned care (£10,721) and primary | **<u>Systemic metastases</u>**<br>• most outpatient visits (10.4);<br>• highest mortality (76.1[35.1%])<br>**<u>Urogenital and gastrointestinal cancers</u>**<br>• high ED attendances (3.1)<br>**<u>Cervical and skin cancers</u>**<br>• highest total care costs (£17,081) and planned care costs (£4,247; 2.6 elective admissions; 10.3 outpatients)<br>**<u>Scoliosis and dementia</u>**<br>• highest unplanned costs (£14,066; 3.2 emergency admissions; 4.1 ED visits)<br>**<u>Cardiometabolic disease</u>**<br>• high total costs (£17,075)<br>**<u>Hip fracture and dementia</u>** | **<u>Personality disorders, liver disease, and metastatic lung cancer</u>**<br>(mean age 67.5)<br>• highest planned care (£4,918; 3.7 elective admissions; 11.4 outpatient visits);<br>• most deprived area (IMD 4-5: 1,609 [44.5%])<br>**<u>Immunological and haematological disorders</u>** (mean age 73.7)<br>• highest total care costs (£18,248) and secondary care costs (£17,707);<br>• smallest cluster (1,166 [10.8%])<br>**<u>Upper GI and prostate multimorbidity</u>** (mean age 79.7)<br>• most GP appointments (15.6) and high unplanned care (£14,388)<br>• most male (1,437 [60.3%]) with most | **<u>Liver disease & mental health</u>** (mean age 57.9)<br>• highest unplanned costs (£13,708; 4.1 emergency admissions; 8.1 ED attendances);<br>• most in deprived area (IMD 4-5: 3,421 [51.0%])<br>**<u>Sleep apnoea, gastro-reflux and diabetes</u>**<br>(mean age 64.8)<br>• relatively low total costs (£14,666) and lowest unplanned costs among clusters (£12,350)<br>**<u>Immunological and haematological disorders</u>** (mean age 67.8)<br>• highest total costs (£16,590), secondary care costs (£16,120) and planned costs (£3,662; 3.3 elective admissions; 12.5 outpatient visits); |
| • highest elective admissions (8.6) and outpatient visits (26.6);<br>• highest mortality (111 [18.8%])<br>**<u>Cardiometabolic complications</u>**<br>• most multimorbid (15.1 diagnoses);<br>**<u>Female pelvic pain, chronic fatigue and urogenital conditions</u>**<br>• highest primary-care costs (£414; 15.4 GP appointments) | • high total care costs (£19,193), highest planned care costs (£9,800) and secondary care costs (£18,790)<br>**<u>Sensory impairment with osteoporosis</u>**<br>• high primary care costs (£476; 17.2 GP appointments);<br>• most minor ethnicity (non-White: 384 [21.7%])<br>**<u>Cardiovascular disease</u>**<br>• most male (1,902 [68.6%]);<br>• intensive secondary care (£18,676) | care costs (£541, 19.3 GP appointments) | • more female (3,241 [58.9%]);<br>• largest cluster (5,499 [31.0%]) | acute conditions (5.1 diagnoses)<br>**<u>Cardiovascular disease and dementia (mean age 83.1)</u>**<br>• highest unplanned care (£14,564) and lowest planned care (£2,737) despite highest primary care (£600) | • most minor ethnicity (non-white: 1,826 [23.2%])<br>• highest mortality (3,275 [41.7%]).<br>**<u>Diabetic and vascular complications (mean age 73.3)</u>**<br>• highest primary care costs (£597; 18.9 GP appointments) and high unplanned £13,412.<br>**<u>Polymyalgia and cardiovascular disease (mean age 81.3)</u>**<br>• most male (6,578 [60.7%])<br>• highest primary care costs (£597; 17.6 GP appointments) and high unplanned care costs (£13,276);<br>• most multimorbid (23.2 diagnoses);<br>• largest cluster (10,845 [20.8%])<br>**<u>Hip fracture, cognitive, and sensory impairment (mean age 82.2)</u>**<br>• most female (6,258 [63.8%]).<br>• lowest total care costs (£14,154) and planned care costs (£1,603) |

Cluster 1 (“Liver disease & mental health”) comprised younger patients in more deprived areas (mean age: 55·8; IMD quintiles 4 and 5: 3,319 [50·9%] of 6,523) with high relative prevalence of alcohol-related liver disease, substance misuse, personality disorders, bipolar affective disorder, and schizophrenia. They had the second highest unplanned care costs (mean £12,476 per person; 3·8 emergency admissions; 7·3 ED attendances) following the eldest Cluster 9.

Cluster 2 (“Sleep apnoea, gastro-reflux and diabetes”) included mid-older and ethnically diverse patients in more deprived area (mean age: 65·6; IMD=4-5: 3,442 [53·2%] of 4,589; non-White: 1,926 [42.0%]) with sleep apnoea, peptic ulcer disease, diabetes, and diabetic eye disease, and high secondary care utilisation (mean £17,391).

Cluster 3 (“Chronic fatigue, immunological and haematological disorders”), with a similar mean age (65·6), comprised patients with immune and haematological conditions and chronic fatigue. This cluster had the highest total and planned care costs (mean £19,107 and £8,684, respectively), reflecting frequent elective admissions (mean 5·9).

Cluster 4 (“Low multimorbidity”) represented patients from the least deprived areas (mean age: 66·2; IMD=4-5: 2,460 [35·9%] of 6,852) and low relative prevalence of major chronic diseases. Their costs were more evenly split between planned and unplanned care (means £7,982 vs. £7,810), and had the lowest mortality across all clusters (715 [10.4%]).

Cluster 5 (“Nodal metastases”) comprised predominantly older women (mean age: 67·3; female: 2,371 [67·5%] of 3,514) with secondary malignancy in lymph nodes. They had the most frequent elective admissions (mean 6·7) and outpatient attendances (mean 21·5), and the highest mortality (1,128 [32.1%]).

Cluster 6 (“Complex multimorbidity”), the smallest cluster (2,842 [5·4%] of 52,175), included mainly older women (mean age: 68·3; female 1,905 [67·0%] of 2,842) with complex multimorbidity including no obvious patterns across specialty but the highest number of comorbidities (mean 21·2 CALIBER diagnoses). They were the heaviest users of primary care (mean 19·4 GP appointments) and frequent ED users (mean 4·8).

Cluster 7 (“Prostate cancer with nodal metastasis”) were predominantly older men (mean age: 72·8; male: 2,645 [68·7%] of 3,850) with prostate cancer, prostate hyperplasia, and secondary malignancy in lymph nodes. This cluster also had a higher relative prevalence of COPD, and diverticular disease. This cluster was characterised by intensive planned care (mean 4·1 elective admissions and 16·0 outpatients) and high mortality (1,201 [31.2%]).

Cluster 8 (“Prostate, stroke, and sensory impairment”) included mainly older men (mean age: 78·9; male: 3,892 [63·4%] of 6,138) with prostate hyperplasia, stroke, macular degeneration, and visual impairment, and had frequent emergency admissions (mean 3·0).

Cluster 9 (“Cardiovascular disease and dementia”), the largest cluster (12,403 [23·8%] of 52,175), captured elderly patients (mean age: 79·2) with cardiomyopathy, valve disorders, pulmonary hypertension, left bundle branch block, and dementia. This group showed the highest unplanned and primary care utilisation (mean: £13,375 and £606, respectively).

### Cluster profiles stratified by age

We observed many similar clusters across age bands, but their expression split in younger adults and combined in older age (Table 4; radar charts of prevalent conditions and detailed statistics in Appendix Figure S3 and Table S5).

Among those aged 18–44, Liver disease and mental health separated into a large mental-health and substance-misuse cluster (1,166 [24·8%] of 4,710) with the highest emergency-department use (mean 9·0), and an alcohol-related liver disease cluster with the highest unplanned costs (mean £14,113), emergency admissions (mean 4·1), more men (452 [59.0%] of 766), and high deprivation (IMD=4-5: 446 [58·2%]). Other clusters included immunological and haematological disorders with the highest secondary-care costs (mean £21,813) and most non-White patients (217 [37·1%] of 585); a cardiometabolic complications cluster with the greatest multimorbidity (mean 15·1 conditions); and a female pelvic-pain, chronic-fatigue and urogenital cluster with the highest primary-care use (mean £414; 15·4 GP contacts). A cancer cluster also appeared, predominantly female (378 [63·9%] of 592) with the highest elective admissions (mean 8·6), outpatient visits (mean 26·6), and mortality (111 [18·8%]).

In mid-life (aged 45–65), the mental-health and liver clusters remained distinct. The liver disease cluster showed the highest total costs (mean £19,332), unplanned costs (mean £12,892), emergency admissions (mean 3·4), ED attendances (mean 5·4), highest primary-care costs (mean £510), most GP appointments (mean 17·9), and high deprivation (IMD=4-5: 909 [54·9%] of 1,654). Mental health and substance misuse continued to show high emergency department utilisation (mean 4·4 ED attendances). Oncology split into a breast-cancer cluster (highest mean elective admissions 7·6; most mean outpatient visits 25·3; highest mortality (463 [33·9%] of 1,365) and a gastrointestinal-cancer cluster (high mean total costs £19,193; highest mean planned secondary-care costs £18,790). A sensory-impairment with osteoporosis cluster had high primary-care use (mean £476; 17·2 GP contacts) and the largest non-White share (384 [21·7%] of 1,769). A cardiovascular-disease cluster also emerged, mostly male (1,902 [68·6%] of 2,772) with high secondary-care use (mean £18,676).

For ages 66–79, several clusters from the main analysis combined. Alcohol-related liver disease persisted but was the smallest cluster (1,330 [7·8%] of 17,130) and remained primary-care heavy, with the highest primary-care costs (mean £577). A broad nodal-metastases cluster re-emerged, showing the highest planned costs (mean £7,968), most outpatient visits (mean 16·9), most elective admissions (mean 5·3), and the highest mortality (1,450 [25·5%] of 5,686). A neurodegenerative and frailty cluster carried the acute burden, with the highest total costs (mean £17,904), highest unplanned costs (mean £12,683), and highest unplanned care activity (mean 3·0 admissions; 4·2 ED visits). The cardiovascular-disease cluster, mostly male (3,752 [66.0%] of 5,687), showed high unplanned care (mean £10,721) and primary-care use (£541; 19·3 GP appointments).

In the 80+ group, clusters existed covering advanced cardiovascular disease, oncology, and frailty. The cardiometabolic cluster showed high total costs (mean £17,075). The scoliosis and dementia cluster carried the greatest unplanned burden with the highest unplanned costs (mean £14,066), emergency admissions (mean 3·2), and ED visits (mean 4·1). Oncology appeared as a systemic-metastases cluster with the most outpatient visits (mean 10·4) and the highest mortality (716 [35·1%] of 2,037); a urogenital–gastrointestinal cancer cluster with high ED use (mean 3·1 visits); and a cervical–skin cancer cluster with the highest total (mean £17,081) and planned costs (mean £4,247) and high elective-admission (mean 2·6) and outpatient activity (mean 10·3). A hip-fracture and dementia cluster was the largest (5,499 [31·0%] of 17,752) and mainly female (3,241 [58·9%] of 5,499).

### Cluster profiles: in-year decedents

Among high-cost patients who died in-year, the optimal hierarchical clustering yielded four age-graded clusters with similar total spend (mean total costs £17,296–£18,248; Table 5; Appendix Figure S4 and Table S6). A younger (mean age: 67·5 years) personality disorders, liver disease, and metastatic lung cancer cluster showed the highest planned care costs (mean £4,918; 3·7 elective admissions; 11·4 outpatient visits). A high proportion of this group lived in deprived area or minor ethnic group (IMD 4-5: 1,609 [44·5%] of 3,616; non-White: 295 [8.2%]). An immunological and haematological disorder cluster (mean age: 73·7) was the smallest group (1,166 [10.8%] of 10,759) carried the highest total (mean £18,248) and secondary-care costs (mean £17,707). An upper GI and prostate multimorbidity cluster were older (mean age: 79·7) and more often male (1,437 [60·3%] of 2,383). This cluster had the most GP appointments (mean 15·6) and most acute conditions (mean 5·1 diagnoses) alongside high unplanned spend (mean £14,388). Finally, a cardiovascular disease and dementia cluster (mean age: 83·1) concentrated the highest unplanned care (mean £14,564) while having the lowest planned costs (mean £2,737) despite the highest primary care spend (mean £600; 15·5 GP appointments).

### Cluster profiles: high-cost patients defined by unplanned care

Among high-cost patients defined by unplanned care, a six-cluster k-medoids output was the optimal solution. Most clusters were similar to the main analysis (Table 5; Appendix Figure S5 and Table S7), except the low multimorbidity, complex multimorbidity, and prostate diagnoses driven clusters were not present.

The liver disease and mental health group (mean age: 57·9) shows the highest unplanned use (mean £13,708; 4·1 emergency admissions; 8·1 ED attendances) and mostly lived in deprived areas (IMD quintiles 4 and 5: 3421 [51.0%] of 6703). The sleep apnoea, gastro-reflux and diabetes cluster (mean age: 64·8) shows relatively low total care costs (mean 14,666) and lowest unplanned spend (mean £12,350). The immunological and haematological cluster (mean age: 67·8) had highest total care costs (mean £16,590) with a sizeable planned-care footprint (mean £3,662; 3·3 elective; 12·5 outpatients) and had the highest mortality (3,275 [41·7%] of 7,860). The diabetic and vascular complication cluster (mean age: 73·3) carries the greatest ongoing management burden (mean primary care costs: £597; 18·9 GP appointments) and high unplanned care costs (£13,412). Two older-age clusters appear. First, a polymyalgia and cardiovascular disease cluster (mean age: 81·3; most male: 6,578 [60·7%] of 10,845) with high levels of multimorbidity (mean 23·2 CALIBER diagnoses). Second, a hip fracture, cognitive and sensory impairment cluster (mean age: 82·2; most female: 6,258 [63·8%] of 9,804) with the lowest planned-care footprint (mean £1,603).

### Robustness checks

The across-method comparison shows stable core clinical structure (see cluster comparison and detailed statistics in Appendix Tables S8-9). Both alternative approaches to clustering generate a matched liver disease and mental health cluster, two oncology driven clusters (nodal metastases and prostate-led), and a low multimorbidity cluster. The cardiovascular and dementia cluster was also reproduced using these two alternative approaches but with more dementia and sensory impairment diagnoses. Compared with the hierarchical nine-cluster solution, the k-medoids clustering produced broader and more integrated groupings, merging the sleep apnoea, gastro-reflux, and diabetes cluster into a new “cardiometabolic disease” cluster, embedding chronic fatigue, immunological, and haematological disorders within “nodal metastases,” and splitting the “prostate, stroke, and sensory impairment” cluster between “prostate cancer (with nodal metastasis)” and “dementia, cardiovascular disease, and sensory impairment”. Compared with the six-cluster K-medoids solution, the seven-cluster fuzzy solution produced similar clusters except chronic fatigue, immunological and haematological disorders was retained as a distinct cluster, bringing it closer to the main nine-theme layout.

Using a second independent 50% random sample (Appendix Tables S10-11), the preferred solution was four clusters from fuzzy k-medoids. The liver disease and mental health cluster was reproduced, comprising younger patients (mean age 57·4 years) higher levels of deprivation (IMD quintiles 4 and 5: 6,129 [48·0%] of 12,761), the highest total care costs (mean £17,853) and the most emergency admissions (mean 3·0) and ED attendances (mean 5·4). Three clusters were reproduced with additional or shifted feature diagnoses. A nodal metastases and chronic-fatigue cluster was characterised by high rates of secondary malignancy in the lymph nodes and chronic fatigue and showed the highest planned-care costs (mean £7,516), alongside the highest in-year mortality (2,986 [25·3%] of 11815). The dementia and cardiovascular-disease cluster comprised older patients (mean age 79·3) with a high prevalence of dementia, stroke, and other cardiovascular conditions, and exhibited the highest unplanned-care costs (mean £12,717) but the lowest planned-care costs (mean £4,297). The prostate and cardiometabolic-complications cluster had increased rates of prostate cancer, prostate hyperplasia, and diabetes- and vascular-related complications, and showed the highest multimorbidity burden (mean 22·4 CALIBER diagnoses) and frequent primary-care use (mean 19·2 GP appointments).

## Discussion

### Summary of Findings

This study examined high-cost patients within the English NHS, using linked primary and secondary care data for over 10 million adults in 2018/19. The top 1% of high-cost patients had average annual costs of £17,485 per person, accounting for £1.8billion (26·8%) of £6.6billion overall healthcare costs, compared with £653 for the full study population. Compared to the full population, high-cost patients were generally older, had more comorbidities, greater deprivation, and higher mortality within the year. Proportionally, fewer high-cost patients were of minority ethnicities when compared to the full population. This may reflect differences in access or recording, warranting further investigation.

Multiple clustering algorithms were tested, with hierarchical clustering and nine clusters demonstrating optimal validity and stability. The resulting clusters identified distinct patient groups with differing demographic, clinical, and socioeconomic profiles, although different forms of clustering and sub-group analyses produced similar results. Notable clusters that were consistently produced across different clustering approaches included: younger people with liver disease and mental health; patients with nodal metastases; patients with prostate cancer and hyperplasia; and older people with cardiovascular disease and dementia.

Our approach to examining high-cost patients extends previous evidence by applying multiple clustering methods, enabling direct comparison of cluster stability and structure, and identifying distinct groups of high-cost patients that are consistently reproduced across different methods and sample selections. Identifying these clusters offers opportunities for more targeted case-management interventions, which may improve care quality and system efficiency.

### Strengths and Limitations

This study has several key strengths. First, our analysis improves on existing literature by clustered high-cost patients into distinct patient groups based on their diagnoses and socioeconomic characteristics, providing a more comprehensive understanding of patient complexity and the multiple drivers of healthcare costs. Second, it utilises one of the largest nationally representative datasets available in England, linking over 10 million adult patient records from primary care (CPRD) with secondary care (HES), deprivation indices, and mortality data, thereby enabling robust, population-level insights that are generalisable across the health system. Third, by incorporating a detailed set of over 300 acute and chronic CALIBER diagnoses, the analysis captures multimorbidity in far greater depth and precision than previous studies that relied on a narrow list of long-term conditions.^38–40,40^ Finally, the use of multiple clustering approaches, combined with systematic evaluation of validity and stability indices, enhances confidence in the clusters identified and their potential use to inform targeted interventions and service design.

However, several limitations should be acknowledged when interpreting the results. First, the calculation of healthcare costs relied on applying national unit costs to recorded service use, which may not capture the full complexity of actual resource use. This is particularly relevant for specialised care and while certain uplifts to tariffs do exist for patient episodes subject to specialised commissioning,^41^ it is not possible to estimate the full costs associated with these episodes as many involve high-cost medicines or devices which are centrally procured by NHS England with confidential pricing arrangements.^42^ Second, we focused our analysis on healthcare costs in primary and secondary care and did not include costs in other NHS settings (i.e. community health or mental health, long-term care), or privately funded care, potentially underestimating true total costs for some patients. Third, while clustering algorithms identified distinct groups, these reflect patterns of co-occurrence and do not imply causality or distinct etiological subtypes. There is also the concern that some individuals may not fit neatly into any one cluster, and despite this all patients are forcibly assigned to clusters even when their profiles are atypical. This may explain how certain clusters produced did not have distinctive individual characteristics or diagnoses when compared to other clusters.

Finally, our decision to limit clusters to three to ten clusters to improve interpretability could have restricted the production of more detailed clusters and insights.

### Future research and policy implications

Our findings have several implications for research and policy. Further research is needed to explore the longitudinal trajectories of these patient groups.^7,43,44^ This is important as patients with recurrent and episodic high costs may experience exacerbations of long-term conditions that could be averted with appropriate secondary prevention interventions. There is also potential to incorporate further socioeconomic characteristics within clustering algorithms such as functional status, social care needs, and patient-reported outcomes, which are often not captured in routine healthcare data but may strongly influence costs and care needs.

Although, the risk of overfitting with high-dimensional data must be considered,^45^ and it will be important to consider both theory-informed and data-driven approaches to prioritise which socioeconomic characteristics should be included within clustering approaches.

The current policy agenda in the England is focused on three priorities including investment in disease prevention, moving care from hospitals to the community, and strengthening digital health.,^46^ and strategies to address the needs of high-cost patients are aligned with all three priorities. However, there remains limited capacity and capabilities to identify and support high-cost patients at the local level. Existing risk-stratification tools, such as the Johns Hopkins Adjusted Clinical Groups (ACG) system that has been applied in the NHS over the last decade,^47^ are typically based on predefined condition lists or utilisation patterns. In contrast, the clustering methods used in our analysis take a data-driven approach that can be applied by local data analysts to uncover context specific subgroups of high-cost patients based on their diagnostic and sociodemographic profiles, offering greater transparency and adaptability to local contexts. Delivering this in practice will require sustained investment in data analytical capacity, including enhanced secure data environments and better integration of datasets across sectors to support service planning and delivery. ^48^

Crucially, once high-cost patients are identified, services must be adapted to their specific needs rather than applying a single model of care. Our findings indicate that a uniform approach to managing high-cost patients is unlikely to be effective, as we identified multiple distinct clusters with differing MLTC profiles and care needs. For example, interventions such as outreach and recovery teams during admissions have been shown to reduce subsequent hospital admissions for patients with substance misuse and mental health issues.^49,50^ Case-management programmes have also been shown to prevent hospital admissions for patients with cardiovascular disease, and diabetes. ^4,5^ Several interventions can also be used to support elderly patients with frailty such as muscle strength training and nutritional supplementation, home visits, and health education.^51^

## Conclusion

In this large national study of primary and secondary care costs, we identified distinct clusters of high-cost patients using alternative clustering methods. These clusters demonstrate substantial diversity in clinical conditions, demographics, healthcare utilisation, and socioeconomic characteristics. These findings provide new insights into the diverse profiles that drive disproportionate healthcare costs and emphasise that high-cost patients cannot be treated as a uniform group. Incorporating cluster-based approaches into population health management approaches may improve the targeting of case management programmes, optimise resource allocation, and ultimately enhance care quality and health system sustainability. Future research should build on these findings to explore longitudinal patterns, integrate data from other healthcare settings and on further socioeconomic characteristics, and evaluate the impact of targeted interventions for these distinct patient populations.

## Data Availability

Access to research data from Clinical Practice Research Datalink (CPRD) is restricted and subject to protocol approval via CPRD research data governance process. Code lists of acute CALIBER diagnoses and statistical code are available on github.

https://github.com/ShaolinWang-codes/Codelist-for-Acute-CALIBER-conditions-used-in-High-Cost-Patient-study

## Acknowledgements

We are grateful to Geraldine Clarke, Joe Hewton, Kathryn Marszalek, Luisa Pettigrew, and Ruth Thorlby of The Health Foundation for their comments on the drafts.

## Contributors

SW, LA, MS, EK, TB, and MA contributed to conceptualisation, investigation, methodology, and review of manuscript; SW contributed data curation, software, and visualisation; SW and MA contributed to formal analysis, validation, and writing of manuscript; MA and LA contributed to funding acquisition; MA contributed to project administration, resource, and supervision. The corresponding author (SW) attests that all listed authors meet authorship criteria and that no others meeting the criteria have been omitted. SW is the guarantor. Transparency: The lead author (the guarantor) affirms that the manuscript is an honest, accurate, and transparent account of the study being reported; that no important aspects of the study have been omitted; and that any discrepancies from the study as planned (and, if relevant, registered) have been explained.

## Provenance and peer review

Not commissioned; externally peer reviewed.

