## Supplementary materials for "Clustering high-cost patients in England using machine learning: a population-based cohort study"

### Text S1 – Dimension reduction

Prior to applying clustering algorithms, we applied multiple strategies for dimension reduction recommended for high-dimensional data as high levels of dimensionality make it conceptually and computationally difficult to converge on a reasonable clustering solution.[1] In our case, the dimensions refer to diagnostic and patient characteristic variables, which are often highly correlated. An important first step in analysing such datasets is to reduce redundancies and correlations among variables, as these can obscure underlying structure and limit the ability to extract meaningful information. First, we discarded all diagnoses with a prevalence lower than one percent following Yan et al (2019) [1]. Second, we removed four diagnoses derived from blood test results, specifically Low HDL-C, Raised LDL-C, Raised total cholesterol and Raised triglycerides since they would have minimal influence on costs but significantly influence clustering due to high prevalence.

Third, we applied the t-distributed stochastic neighbour embedding (t-SNE) methodology, [2] following the methodology used by Yan et al (2019) to visualise complex patterns in the data and explore potential clustering structure.[1] This involves first using multiple correspondence analysis (MCA) to convert the high-dimensional binary data into low-dimensional continuous data.[3] Unlike Principal Component Analysis, which is designed for continuous variables and relies on a covariance/correlation matrix between diagnostic dummy variables, MCA is specifically designed for categorical/binary data, making it more appropriate for clustering diagnoses from binary or coded clinical variables. We retained sufficient MCA components that explained approximately 90% of the variation across the original sample, and used these as input to t-SNE. We did this instead of using the first two MCA dimensions alone because patients appeared randomly distributed in the 2D MCA space, offering limited visual separation of potential clusters, whereas supplying t-SNE with a richer embedding (up to100 dimensions) preserved more of the underlying data structure and yielded clearer neighbourhood patterns. The t-SNE[1] was implemented using the Barnes-Hut algorithm, which returns a solution that minimizes Kullback-Leibler divergence, a measure of the difference of probability distributions between the original data and the lower-dimensional representation.

[1] We run Barnes-Hut t-SNE for 1,000 iterations with the default perplexity parameter of 30 and the default speed/accuracy trade-off parameter (theta) of 0.5.

#### Text S2 – Clustering algorithms

We applied three clustering algorithms (Table S1) to the t-SNE embedding. First we applied connectivity-based clustering, using the agglomerative hierarchical clustering with Ward's criterion.[4,5] This treats each observation as a singleton cluster, and then merges the most similar clusters sequentially until the final optimal clusters are formed. At each step, the Ward's criterion merges the pairs of clusters that minimize the variance of the clusters being merged. The result is a tree-like dendrogram to visualize the entire cluster hierarchy, where cutting the hierarchy at different levels causes a different number of clusters.

Second, we applied centroid-based clustering with the k-medoids algorithm, which requires the number of clusters to be pre-specified (k), and then accordingly selects k observations as the medoid of each cluster and assign observations to the cluster based on the shortest path to a medoid.[6] The algorithm optimises the selection of medoids to create the shortest total distance from all data points to their nearest medoid. By using actual data points instead of taking the mean as cluster representatives, k-medoids is more flexible than hierarchical and k-means in handling non-spherical or irregularly shaped clusters, and less sensitive to noise and outlier.

Finally, we applied soft clustering, using fuzzy k-medoids, a clustering algorithm that assigns each data point to multiple clusters with varying degrees of membership, rather than placing it into just one group.[7] The algorithm iteratively updates both the fuzzy membership values and the medoids based on distances and a fuzziness parameter, allowing it to model overlapping clusters while maintaining stability in the presence of extreme values. Fuzzy k-medoids extends k-medoids to capture uncertainty and reflects the reality of overlapping groups.

### Table S1 – Descriptive of different clustering algorithms

| Method | Computational approach | Advantages | Limitations |
|---|---|---|---|
| **Hierarchical clustering** | Builds a hierarchy of clusters by successively merging or splitting based on a distance metric and linkage rule; results shown in a dendrogram. | - No need to pre-specify number of clusters (can "cut" dendrogram at different levels).<br>- Flexible choice of distance/linkage methods.<br>- Intuitive tree-like structure for visualization. | - Computationally intensive for large samples.<br>- Early mistakes in merging/splitting cannot be corrected.<br>- No direct "representative" observation for clusters.<br>- Sensitive to noise/outliers. |
| **K-medoids** | Partitions data into *k* clusters by selecting representative observations (medoids) that minimise total dissimilarity within clusters. | - More robust to outliers than hierarchical and k-means (medoids are real data points).<br>- Works with a variety of distance measure (e.g., Euclidean, Manhattan, Jaccard, Gower).<br>- Provides interpretable representative cases (medoids). | - Less efficient than k-means on very large datasets.<br>- Requires *k* to be pre-specified.<br>- Every observation is forced into a cluster, even borderline or noisy points. |
| **Fuzzy k-medoids** | Extension of k-medoids allowing each observation to have partial membership across clusters (membership weights sum to 1). | - Captures uncertainty and overlap between clusters.<br>- More realistic for heterogeneous populations.<br>- Membership weights can be used in further analyses. | - Requires *k* and fuzziness parameter to be set.<br>- Interpretation and reporting more complex (no single definitive cluster label).<br>- Computationally more demanding than hard clustering. |

#### Text S3 – Cluster evaluation and selection

We applied each clustering method over a range of cluster numbers (k = 3 to 10), which were selected to improve interpretability of our clusters. We selected the optimal method-k combination by combining clustering validity ranks and cross-method stability, prioritizing solutions that demonstrated both high quality and consistency.

Following Arbelaitz et al (2013)[8], we compared multiple validity indices that assess cluster quality based on cohesion (within-cluster variance) and separation (between-cluster variance):

- Silhouette index estimates cohesion via the distance between all the points in the same cluster and separation via the nearest neighbour distance.[9]
- Davies–Bouldin index (DBI) estimates cohesion via the distance from the points in a cluster to its centroid and separation via the distance between centroids.[10]
- Calinski–Harabasz Index (CHI) estimates cohesion via the distances from the points in a cluster to its centroid and separation via the distance from the centroids to the global centroid.[11]

Within each method, clustering solutions were ranked by Silhouette (descending; high is better), DBI (ascending; lower is better) and CHI (descending; high is better) across k. We computed a combined rank by summing the three individual metric ranks, which was subsequently negated and standardized to yield a composite validity score. Higher scores indicate better defined clustering structure.

For each value of k, we computed Adjusted Rand Index (ARI) from pairwise comparisons between clustering methods (hierarchical vs. k-medoids, hierarchical vs. fuzzy k-medoids, k-medoids vs. fuzzy k-medoids). ARI measures similarity between two clustering approaches, adjusted for chance agreement (range between -1 and 1; 1 indicating perfect agreement). For each clustering method we averaged its two ARIs (e.g. hierarchical to be compared with k-medoids and fuzzy k-medoids), and standardized to obtain a ARI score.

The final score for each method-k combination was computed as the equal-weigh average of the standardized validity score and the standardized ARI score. We used this approach to select the optimal clustering solution within each analysis that is both valid and stable across methods and values of k.

**Figure S1 – Distribution of healthcare costs**

Panel a. full study population

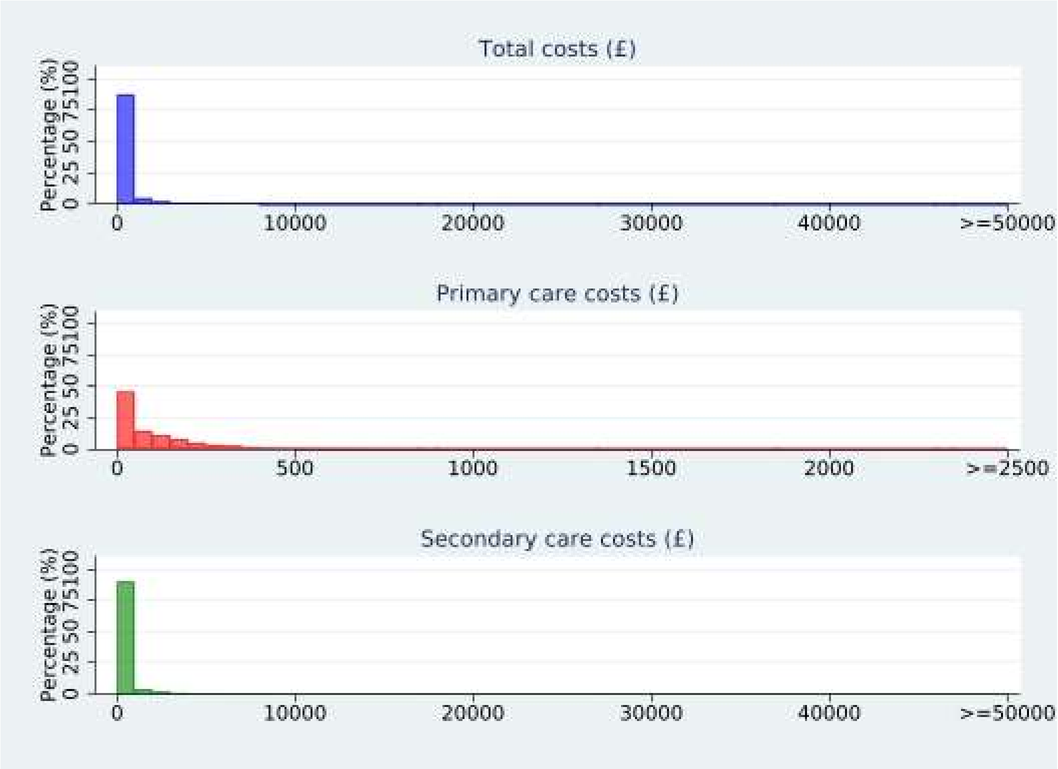

Panel b. top 1% high-cost patients

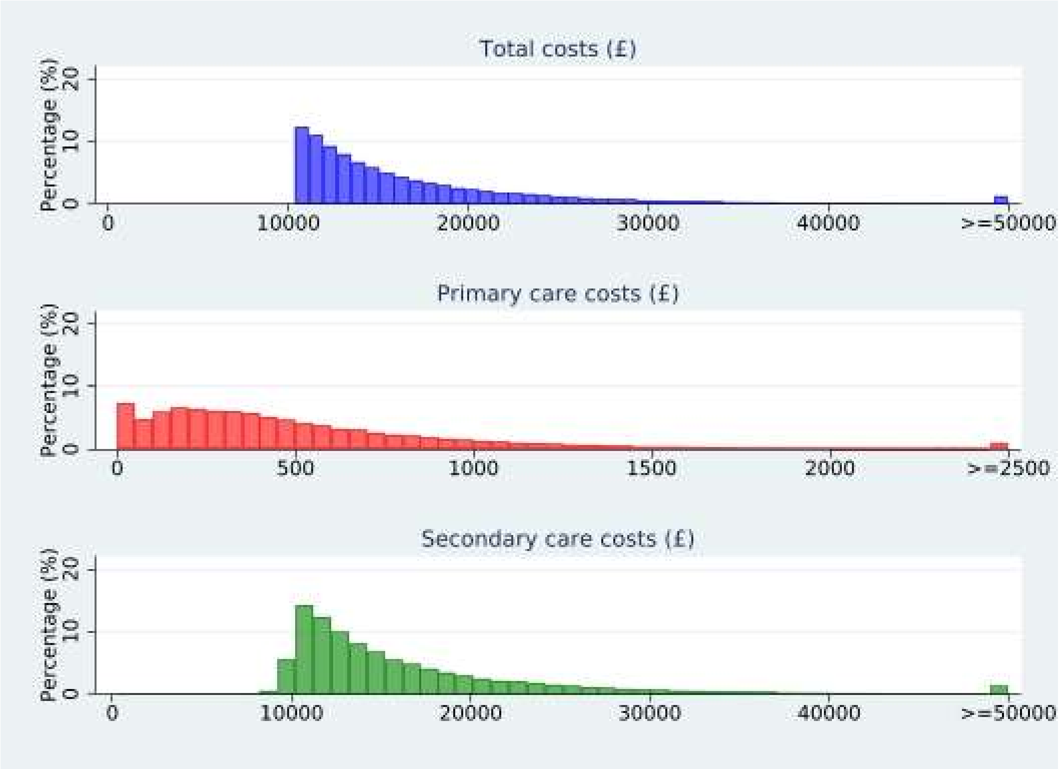

#### Text S4 – Dimension reduction and cluster evaluation and selection

MCA was used to transform categorical variables into continuous dimensions. The first two dimensions explained 17.2% and 5.5% of the total variance, respectively. A total of 82 conditions and age group variables either contributed more than expected or were better represented in these two dimensions (Table S2). An additional 13 conditions and the in-year-death indicator loaded mainly on the third dimension, capturing 4.1% of the variance. The top 100 MCA components, collectively explaining 86.5% of the variance in the original dataset, were then used as input to t-SNE to obtain a two-dimensional map for clustering and visualisation.

The optimal clustering solution was selected by combining clustering validity and stability (Table S3). The preferred number of clusters, *k,* were identified as 9 or 10 for hierarchical, 4 or 6 for k-medoids, and 7 for fuzzy k-medoids. For hierarchical, at $k = 9$ and 10, silhouette coefficients were 0.302 and 0.304 (ranked 3rd and 2nd), DBI were 0.934 and 0.999 (ranked 1st and 3rd), and CHI were 33,900 and 35,292 (ranked 2nd and 1st), yielding the same best combined rank (6) for both *k* values.

Stability, assessed by ARI peaked at $k = 9$ across methods: hierarchical and k-medoids 0.583, hierarchical and fuzzy 0.550, and k-medoids and fuzzy 0.553. Although k-medoids achieved marginally higher agreement with the other methods (mean ARI = 0.568) than hierarchical (0.566) and fuzzy k-medoids (0.551), the difference was small and therefore not decisive on their own. Considering validity and stability jointly support hierarchical clustering with nine clusters as the most robust and optimal solution overall, followed by k-medoids with six clusters and fuzzy k-medoids with seven clusters.

**Table S2 – Percent contributions and squared cosines of each condition for dimensions 1-3, derived from MCA.**

Highlighted values indicate contributions that are higher than expected, or good representation of that variable to the dimension.

| | % Contrib1 | % Contrib2 | % Contrib2 | Cos1 | Cos2 | cos3 |
|---|---|---|---|---|---|---|
| Abdominal aortic aneurysm | 0.17 | 0.03 | 0.01 | 0.12 | 0.01 | 0.00 |
| Abdominal hernia | 0.11 | 0.04 | 0.06 | 0.08 | 0.01 | 0.01 |
| Acne | 0.20 | 0.11 | 0.08 | 0.14 | 0.03 | 0.01 |
| Actinic keratosis | 0.03 | 0.08 | 0.01 | 0.02 | 0.01 | 0.00 |
| Acute kidney injury | 0.99 | 0.20 | 1.14 | 0.44 | 0.03 | 0.12 |
| Agranulocytosis | 0.13 | 0.01 | 0.84 | 0.09 | 0.00 | 0.13 |
| Alcohol misuse | 0.07 | 2.69 | 0.03 | 0.04 | 0.44 | 0.00 |
| Alcoholic liver disease | 0.04 | 3.77 | 0.16 | 0.02 | 0.50 | 0.02 |
| Allergic and chronic rhinitis | 0.00 | 0.06 | 0.08 | 0.00 | 0.02 | 0.02 |
| Anterior and intermediate uveitis | 0.01 | 0.01 | 0.01 | 0.01 | 0.00 | 0.00 |
| Anxiety disorders | 0.01 | 1.66 | 0.25 | 0.01 | 0.29 | 0.03 |
| Appendicitis | 0.03 | 0.00 | 0.00 | 0.03 | 0.00 | 0.00 |
| Aspiration pneumonitis | 0.06 | 0.00 | 0.06 | 0.05 | 0.00 | 0.01 |
| Asthma | 0.08 | 0.88 | 0.07 | 0.05 | 0.17 | 0.01 |
| Atrial fibrillation | 2.05 | 0.20 | 0.02 | 0.71 | 0.02 | 0.00 |
| Atrioventricular blocks | 0.60 | 0.06 | 0.03 | 0.36 | 0.01 | 0.00 |
| Bacterial diseases (excl TB) | 0.65 | 0.33 | 0.82 | 0.23 | 0.04 | 0.07 |
| Barrett's oesophagus | 0.05 | 0.12 | 0.01 | 0.04 | 0.03 | 0.00 |
| Benign neoplasm of colon, rectum, anus and anal canal | 0.00 | 0.02 | 0.02 | 0.00 | 0.00 | 0.00 |
| Benign neoplasm of ovary | 0.03 | 0.04 | 0.01 | 0.02 | 0.01 | 0.00 |
| Bipolar affective disorder and mania | 0.02 | 0.55 | 0.09 | 0.01 | 0.12 | 0.01 |
| Bronchiectasis | 0.15 | 0.12 | 0.04 | 0.10 | 0.03 | 0.01 |
| COPD | 0.64 | 0.57 | 0.18 | 0.31 | 0.09 | 0.02 |
| Carcinoma in situ_cervical | 0.02 | 0.07 | 0.01 | 0.01 | 0.01 | 0.00 |
| Carpal tunnel syndrome | 0.00 | 0.00 | 0.06 | 0.00 | 0.00 | 0.01 |
| Cataract | 1.58 | 0.18 | 0.05 | 0.61 | 0.02 | 0.00 |
| Cholecystitis | 0.00 | 0.07 | 0.00 | 0.00 | 0.02 | 0.00 |
| Cholelithiasis | 0.02 | 0.22 | 0.03 | 0.02 | 0.05 | 0.01 |
| Chronic fatigue syndrome | 0.01 | 0.27 | 0.16 | 0.00 | 0.06 | 0.03 |
| Chronic kidney disease | 2.50 | 0.16 | 0.00 | 0.77 | 0.02 | 0.00 |
| Chronic sinusitis | 0.00 | 0.07 | 0.03 | 0.00 | 0.02 | 0.01 |
| Cirrhosis | 0.01 | 4.13 | 0.24 | 0.00 | 0.51 | 0.02 |
| Collapsed vertebra | 0.07 | 0.00 | 0.04 | 0.05 | 0.00 | 0.01 |
| Coronary heart disease NOS | 2.58 | 0.01 | 0.21 | 0.70 | 0.00 | 0.01 |
| Crohn's disease | 0.02 | 0.11 | 0.01 | 0.01 | 0.03 | 0.00 |
| Delirium, not induced by alcohol and other substances | 0.57 | 0.01 | 0.13 | 0.31 | 0.00 | 0.02 |
| Dementia | 0.56 | 0.26 | 0.08 | 0.27 | 0.04 | 0.01 |
| Depression | 0.00 | 2.43 | 0.35 | 0.00 | 0.40 | 0.04 |
| Dermatitis | 0.08 | 0.10 | 0.01 | 0.06 | 0.02 | 0.00 |
| Diabetes | 0.99 | 0.40 | 0.02 | 0.39 | 0.05 | 0.00 |
| Diabetic eye disease | 0.73 | 0.19 | 0.06 | 0.31 | 0.03 | 0.01 |
| Diabetic neuropathy | 0.37 | 0.46 | 0.04 | 0.18 | 0.07 | 0.00 |
| Diaphragmatic hernia | 0.36 | 0.49 | 0.00 | 0.19 | 0.09 | 0.00 |
| Diverticular disease of intestine | 0.51 | 0.01 | 0.01 | 0.29 | 0.00 | 0.00 |
| Dysmenorrhoea | 0.11 | 0.14 | 0.11 | 0.07 | 0.03 | 0.02 |
| Endometriosis | 0.08 | 0.11 | 0.04 | 0.06 | 0.03 | 0.01 |
| Enthesopathies & synovial disorders | 0.09 | 0.04 | 0.17 | 0.06 | 0.01 | 0.03 |
| Epilepsy | 0.00 | 0.41 | 0.08 | 0.00 | 0.09 | 0.01 |
| Fatty Liver | 0.00 | 2.34 | 0.02 | 0.00 | 0.44 | 0.00 |
| Female genital prolapse | 0.00 | 0.00 | 0.03 | 0.00 | 0.00 | 0.01 |
| Fibromatoses | 0.01 | 0.00 | 0.00 | 0.01 | 0.00 | 0.00 |
| Folate deficiency anaemia | 0.07 | 0.16 | 0.00 | 0.06 | 0.04 | 0.00 |
| Fracture of hip | 0.07 | 0.21 | 0.08 | 0.04 | 0.05 | 0.01 |
| Gastritis and duodenitis | 0.22 | 1.76 | 0.00 | 0.12 | 0.30 | 0.00 |
| Gastro-oesophageal reflux disease | 0.08 | 0.58 | 0.08 | 0.05 | 0.12 | 0.01 |
| Glaucoma | 0.23 | 0.07 | 0.02 | 0.16 | 0.01 | 0.00 |
| Glomerulonephritis | 0.14 | 0.05 | 0.00 | 0.10 | 0.01 | 0.00 |
| Gout | 0.47 | 0.00 | 0.01 | 0.29 | 0.00 | 0.00 |
| Hearing loss | 0.62 | 0.15 | 0.02 | 0.35 | 0.03 | 0.00 |
| Heart failure | 2.75 | 0.00 | 0.02 | 0.78 | 0.00 | 0.00 |
| Hyperparathyroidism | 0.07 | 0.01 | 0.00 | 0.05 | 0.00 | 0.00 |
| Hyperplasia of prostate | 0.40 | 0.18 | 0.09 | 0.22 | 0.03 | 0.01 |
| Hypersplenism | 0.00 | 1.40 | 0.18 | 0.00 | 0.28 | 0.03 |
| Hypertension | 2.44 | 0.05 | 0.02 | 0.77 | 0.01 | 0.00 |
| Infection of skin and subcutaneous tissues | 0.18 | 0.29 | 0.08 | 0.12 | 0.06 | 0.01 |
| Infections of other or unspecified organs | 0.19 | 0.34 | 0.85 | 0.10 | 0.05 | 0.10 |
| Infections of the digestive system | 0.05 | 0.47 | 0.63 | 0.03 | 0.10 | 0.10 |

| | | | | | | |
|---|---|---|---|---|---|---|
| Intellectual disability | 0.03 | 0.19 | 0.03 | 0.02 | 0.04 | 0.00 |
| Intervertebral disc disorders | 0.02 | 0.12 | 0.19 | 0.01 | 0.02 | 0.03 |
| Intracerebral haemorrhage | 0.00 | 0.01 | 0.04 | 0.00 | 0.00 | 0.01 |
| Iron deficiency anaemia | 0.28 | 0.18 | 0.04 | 0.19 | 0.04 | 0.01 |
| Irritable bowel syndrome | 0.01 | 0.35 | 0.14 | 0.00 | 0.08 | 0.02 |
| Ischaemic stroke | 0.33 | 0.11 | 0.17 | 0.17 | 0.02 | 0.02 |
| Left bundle branch block | 0.62 | 0.04 | 0.01 | 0.35 | 0.01 | 0.00 |
| Leiomyoma of uterus | 0.03 | 0.01 | 0.00 | 0.02 | 0.00 | 0.00 |
| Lower respiratory tract infections | 1.06 | 0.30 | 2.17 | 0.37 | 0.03 | 0.18 |
| Macular degeneration | 0.49 | 0.12 | 0.03 | 0.29 | 0.02 | 0.00 |
| Menorrhagia and polymenorrhoea | 0.06 | 0.08 | 0.04 | 0.05 | 0.02 | 0.01 |
| Migraine | 0.08 | 0.22 | 0.24 | 0.06 | 0.05 | 0.04 |
| Multiple valve disorder | 1.03 | 0.01 | 0.00 | 0.46 | 0.00 | 0.00 |
| Myocardial infarction | 1.67 | 0.00 | 0.13 | 0.58 | 0.00 | 0.01 |
| Neuropahtic bladder | 0.01 | 0.06 | 0.09 | 0.01 | 0.01 | 0.02 |
| Non-Hodgkin lymphoma | 0.00 | 0.00 | 0.13 | 0.00 | 0.00 | 0.02 |
| Nonrheumatic aortic valve disorders | 0.73 | 0.07 | 0.03 | 0.39 | 0.01 | 0.00 |
| Nonrheumatic mitral valve disorders | 0.62 | 0.01 | 0.01 | 0.34 | 0.00 | 0.00 |
| Obesity | 0.21 | 0.46 | 0.07 | 0.12 | 0.09 | 0.01 |
| Obstructive and reflux uropathy | 0.01 | 0.00 | 0.58 | 0.01 | 0.00 | 0.09 |
| Oesophagitis and oesophageal ulcer | 0.01 | 0.52 | 0.05 | 0.00 | 0.12 | 0.01 |
| Osteoarthritis (excl spine) | 0.85 | 0.03 | 0.29 | 0.39 | 0.00 | 0.03 |
| Osteoporosis | 0.42 | 0.00 | 0.08 | 0.22 | 0.00 | 0.01 |
| Other anaemias | 0.80 | 0.33 | 0.35 | 0.40 | 0.05 | 0.04 |
| Other cardiomyopathy | 0.22 | 0.01 | 0.01 | 0.14 | 0.00 | 0.00 |
| Other or unspecified infectious organisms | 0.89 | 0.55 | 3.09 | 0.31 | 0.06 | 0.25 |
| Pancreatitis | 0.00 | 0.76 | 0.01 | 0.00 | 0.18 | 0.00 |
| Parkinson's disease | 0.06 | 0.03 | 0.03 | 0.04 | 0.01 | 0.01 |
| Peptic ulcer disease | 0.02 | 0.33 | 0.07 | 0.02 | 0.08 | 0.01 |
| Pericardial effusion | 0.05 | 0.01 | 0.11 | 0.04 | 0.00 | 0.02 |
| Peripheral arterial disease | 0.59 | 0.06 | 0.00 | 0.34 | 0.01 | 0.00 |
| Peripheral neuropathy | 0.16 | 0.57 | 0.01 | 0.09 | 0.11 | 0.00 |
| Peritonitis | 0.03 | 0.12 | 0.14 | 0.02 | 0.03 | 0.02 |
| Personality disorders | 0.07 | 0.97 | 0.20 | 0.04 | 0.19 | 0.03 |
| Pleural effusion | 0.51 | 0.11 | 1.46 | 0.26 | 0.02 | 0.18 |
| Pleural plaque | 0.10 | 0.00 | 0.10 | 0.08 | 0.00 | 0.02 |
| Polymyalgia rheumatica | 0.17 | 0.01 | 0.03 | 0.12 | 0.00 | 0.00 |
| Portal hypertension | 0.01 | 3.31 | 0.25 | 0.00 | 0.45 | 0.03 |
| Primary malignancy_bladder | 0.03 | 0.06 | 0.18 | 0.02 | 0.01 | 0.03 |
| Primary malignancy_bowel | 0.05 | 0.13 | 0.76 | 0.03 | 0.03 | 0.12 |
| Primary malignancy_breast | 0.09 | 0.05 | 0.23 | 0.06 | 0.01 | 0.04 |
| Primary malignancy_lung | 0.01 | 0.02 | 1.23 | 0.01 | 0.00 | 0.20 |
| Primary malignancy_melanoma | 0.00 | 0.06 | 0.11 | 0.00 | 0.01 | 0.02 |
| Primary malignancy_other | 0.08 | 0.05 | 1.60 | 0.05 | 0.01 | 0.25 |
| Primary malignancy_prostate | 0.03 | 0.26 | 0.44 | 0.02 | 0.06 | 0.07 |
| Primary malignancy_skin | 0.21 | 0.33 | 0.03 | 0.14 | 0.07 | 0.00 |
| Psoriasis | 0.01 | 0.13 | 0.00 | 0.00 | 0.03 | 0.00 |
| Pulmonary collapse (excl pneumothorax) | 0.02 | 0.09 | 0.50 | 0.02 | 0.02 | 0.09 |
| Pulmonary embolism | 0.00 | 0.00 | 0.50 | 0.00 | 0.00 | 0.09 |
| Pulmonary fibrosis | 0.15 | 0.02 | 0.06 | 0.11 | 0.01 | 0.01 |
| Raynaud's syndrome | 0.00 | 0.01 | 0.01 | 0.00 | 0.00 | 0.00 |
| Respiratory failure | 0.20 | 0.35 | 0.57 | 0.12 | 0.06 | 0.08 |
| Retinal vascular occlusions | 0.11 | 0.02 | 0.01 | 0.08 | 0.00 | 0.00 |
| Rheumatic valve disorder | 0.26 | 0.03 | 0.03 | 0.17 | 0.01 | 0.00 |
| Rheumatoid arthritis | 0.13 | 0.05 | 0.04 | 0.09 | 0.01 | 0.01 |
| Right bundle branch block combinations | 0.43 | 0.01 | 0.01 | 0.28 | 0.00 | 0.00 |
| Rosacea | 0.00 | 0.00 | 0.00 | 0.00 | 0.00 | 0.00 |
| Schizophrenia | 0.02 | 0.55 | 0.06 | 0.01 | 0.12 | 0.01 |
| Scoliosis | 0.00 | 0.00 | 0.02 | 0.00 | 0.00 | 0.00 |
| Seborrheic dermatitis | 0.01 | 0.05 | 0.01 | 0.01 | 0.01 | 0.00 |
| Secondary malignancy_bone | 0.10 | 0.28 | 3.82 | 0.05 | 0.04 | 0.44 |
| Secondary malignancy_liver | 0.21 | 0.17 | 3.69 | 0.10 | 0.03 | 0.42 |
| Secondary malignancy_lung | 0.15 | 0.17 | 3.54 | 0.07 | 0.03 | 0.42 |
| Secondary malignancy_lymph nodes | 0.32 | 0.18 | 2.95 | 0.16 | 0.03 | 0.34 |
| Secondary malignancy_other | 0.19 | 0.26 | 4.54 | 0.08 | 0.04 | 0.48 |
| Secondary pulmonary hypertension | 0.38 | 0.05 | 0.04 | 0.23 | 0.01 | 0.01 |
| Secondary thrombocytopaenia | 0.00 | 0.69 | 0.43 | 0.00 | 0.15 | 0.07 |
| Sleep apnoea | 0.08 | 0.39 | 0.05 | 0.05 | 0.08 | 0.01 |
| Spinal stenosis | 0.09 | 0.01 | 0.13 | 0.05 | 0.00 | 0.02 |
| Spondylolisthesis | 0.02 | 0.01 | 0.10 | 0.02 | 0.00 | 0.02 |
| Spondylosis | 0.32 | 0.04 | 0.20 | 0.18 | 0.01 | 0.03 |
| Stable angina | 1.66 | 0.00 | 0.28 | 0.57 | 0.00 | 0.02 |
| Stroke NOS | 0.52 | 0.06 | 0.18 | 0.25 | 0.01 | 0.02 |
| Substance misuse | 0.08 | 1.51 | 0.05 | 0.05 | 0.30 | 0.01 |
| Supraventricular tachycardia | 0.09 | 0.01 | 0.00 | 0.07 | 0.00 | 0.00 |

| | | | | | | |
|---|---|---|---|---|---|---|
| Thyroid disease | 0.22 | 0.01 | 0.04 | 0.15 | 0.00 | 0.01 |
| Tinnitus | 0.01 | 0.00 | 0.02 | 0.01 | 0.00 | 0.00 |
| Transient ischaemic attack | 0.47 | 0.05 | 0.15 | 0.27 | 0.01 | 0.02 |
| Tubulo-interstitial nephritis | 0.02 | 0.20 | 0.00 | 0.01 | 0.05 | 0.00 |
| Ulcerative colitis | 0.00 | 0.06 | 0.00 | 0.00 | 0.01 | 0.00 |
| Unstable angina | 0.58 | 0.03 | 0.18 | 0.31 | 0.01 | 0.02 |
| Urinary incontinence | 0.20 | 0.08 | 0.17 | 0.13 | 0.02 | 0.03 |
| Urinary tract infections | 0.66 | 0.01 | 0.33 | 0.27 | 0.00 | 0.03 |
| Urolithiasis | 0.00 | 0.03 | 0.02 | 0.00 | 0.01 | 0.00 |
| Venous thromboembolic disease (Excl PE) | 0.01 | 0.17 | 0.21 | 0.01 | 0.04 | 0.04 |
| Ventricular tachycardia | 0.15 | 0.00 | 0.01 | 0.10 | 0.00 | 0.00 |
| Viral diseases (excl chronic hepatitis/HIV) | 0.00 | 0.12 | 0.07 | 0.00 | 0.03 | 0.01 |
| Visual impairment and blindness | 0.30 | 0.00 | 0.05 | 0.20 | 0.00 | 0.01 |
| Vitamin B12 deficiency anaemia | 0.18 | 0.04 | 0.02 | 0.13 | 0.01 | 0.00 |

**Table S3 – Cluster evaluation results.**

a. Validity

| | k | Silhouette | DBI | CHI | Rank Silhouette | Rank DBI | Rank CHI | Rank combined |
|---|---|---|---|---|---|---|---|---|
| Hierarchical | 3 | 0.318 | 1.117 | 30,548 | 1 | 7 | 6 | 14 |
| | 4 | 0.286 | 0.999 | 28,827 | 4 | 4 | 8 | 16 |
| | 5 | 0.271 | 1.159 | 29,374 | 8 | 8 | 7 | 23 |
| | 6 | 0.285 | 0.999 | 31,876 | 5 | 5 | 5 | 15 |
| | 7 | 0.284 | 1.003 | 33,260 | 6 | 6 | 3 | 15 |
| | 8 | 0.280 | 0.993 | 33,097 | 7 | 2 | 4 | 13 |
| | **9** | **0.302** | **0.934** | **33,900** | **3** | **1** | **2** | **6** |
| | **10** | **0.304** | **0.999** | **35,292** | **2** | **3** | **1** | **6** |
| K-medoids | 3 | 0.358 | 1.028 | 35,348 | 1 | 5 | 6 | 12 |
| | **4** | **0.342** | **1.008** | **37,916** | **2** | **3** | **1** | **6** |
| | 5 | 0.323 | 0.958 | 35,672 | 4 | 2 | 4 | 10 |
| | **6** | **0.324** | **0.948** | **36,853** | **3** | **1** | **2** | **6** |
| | 7 | 0.295 | 1.029 | 34,842 | 8 | 6 | 7 | 21 |
| | 8 | 0.302 | 1.053 | 35,382 | 6 | 7 | 5 | 18 |
| | 9 | 0.300 | 1.076 | 34,488 | 7 | 8 | 8 | 23 |
| | 10 | 0.316 | 1.026 | 36,481 | 5 | 4 | 3 | 12 |
| Fuzzy k-medoids | 3 | 0.357 | 1.027 | 35,266 | 1 | 8 | 7 | 16 |
| | 4 | 0.332 | 0.998 | 36,663 | 7 | 7 | 6 | 20 |
| | 5 | 0.316 | 0.985 | 33,663 | 8 | 5 | 8 | 21 |
| | 6 | 0.350 | 0.900 | 39,457 | 3 | 1 | 5 | 9 |
| | **7** | **0.353** | **0.909** | **40,997** | **2** | **2** | **1** | **5** |
| | 8 | 0.343 | 0.938 | 40,211 | 4 | 4 | 2 | 10 |
| | 9 | 0.333 | 0.989 | 39,590 | 6 | 6 | 4 | 16 |
| | 10 | 0.336 | 0.920 | 40,039 | 5 | 3 | 3 | 11 |

b. Stability

| k | | ARI Hierarchical | K-medoids | Fuzzy k-medoids | average |
|---|---|---|---|---|---|
| 3 | Hierarchical | | 0.311 | 0.399 | 0.355 |
| | K-medoids | 0.311 | | 0.812 | 0.562 |
| | Fuzzy k-medoids | 0.399 | 0.812 | | **0.606** |
| 4 | Hierarchical | | 0.397 | 0.308 | 0.353 |
| | K-medoids | 0.397 | | 0.368 | 0.383 |
| | Fuzzy k-medoids | 0.308 | 0.368 | | 0.338 |
| 5 | Hierarchical | | 0.510 | 0.394 | 0.452 |
| | K-medoids | 0.510 | | 0.448 | 0.479 |
| | Fuzzy k-medoids | 0.394 | 0.448 | | 0.421 |
| 6 | Hierarchical | | 0.443 | 0.380 | 0.411 |
| | K-medoids | 0.443 | 0.380 | 0.619 | 0.481 |
| | Fuzzy k-medoids | 0.380 | 0.619 | | 0.499 |
| 7 | Hierarchical | | 0.479 | 0.470 | 0.475 |
| | K-medoids | 0.479 | | 0.491 | 0.485 |
| | Fuzzy k-medoids | 0.470 | 0.491 | | 0.480 |
| 8 | Hierarchical | | 0.560 | 0.445 | 0.503 |
| | K-medoids | 0.560 | | 0.520 | 0.540 |
| | Fuzzy k-medoids | 0.445 | 0.520 | | 0.483 |
| 9 | Hierarchical | | 0.583 | 0.550 | **0.566** |
| | K-medoids | 0.583 | | 0.553 | **0.568** |
| | Fuzzy k-medoids | 0.550 | 0.553 | | 0.551 |
| 10 | Hierarchical | | 0.507 | 0.588 | 0.548 |
| | K-medoids | 0.507 | | 0.521 | 0.514 |
| | Fuzzy k-medoids | 0.588 | 0.521 | | 0.554 |

c. Standardised validity and stability scores

| | k | Validity combined rank | ARI average | Final score |
|---|---|---|---|---|
| **Hierarchical** | **9** | **1.33** | **1.07** | **1.20** |
| Hierarchical | 10 | 1.33 | 0.82 | 1.13 |
| K-medoids | 6 | 1.33 | 0.60 | 1.04 |
| Fuzzy k-medoids | 7 | 1.51 | -0.07 | 0.88 |
| Fuzzy k-medoids | 10 | 0.44 | 0.91 | 0.63 |
| K-medoids | 3 | 0.27 | 1.01 | 0.56 |
| Fuzzy k-medoids | 3 | -0.44 | 1.59 | 0.37 |
| Fuzzy k-medoids | 6 | 0.80 | 0.18 | 0.55 |
| K-medoids | 10 | 0.27 | 0.37 | 0.31 |
| Fuzzy k-medoids | 8 | 0.62 | -0.04 | 0.36 |
| K-medoids | 5 | 0.62 | -0.09 | 0.34 |
| Fuzzy k-medoids | 9 | -0.44 | 0.87 | 0.08 |
| Hierarchical | 8 | 0.09 | 0.22 | 0.14 |
| K-medoids | 4 | 1.33 | -1.37 | 0.25 |
| K-medoids | 8 | -0.80 | 0.72 | -0.19 |
| Hierarchical | 7 | -0.27 | -0.15 | -0.22 |
| K-medoids | 9 | -1.68 | 1.09 | -0.57 |
| Hierarchical | 6 | -0.27 | -0.98 | -0.55 |
| K-medoids | 7 | -1.33 | -0.01 | -0.80 |
| Hierarchical | 3 | -0.09 | -1.73 | -0.75 |
| Hierarchical | 5 | -1.68 | -0.45 | -1.19 |
| Fuzzy k-medoids | 5 | -1.33 | -0.86 | -1.14 |
| Hierarchical | 4 | -0.44 | -1.76 | -0.97 |
| Fuzzy k-medoids | 4 | -1.15 | -1.96 | -1.47 |

**Figure S2: Disease-system profiles of high-cost clusters (optimal hierarchical 9-cluster solution)**

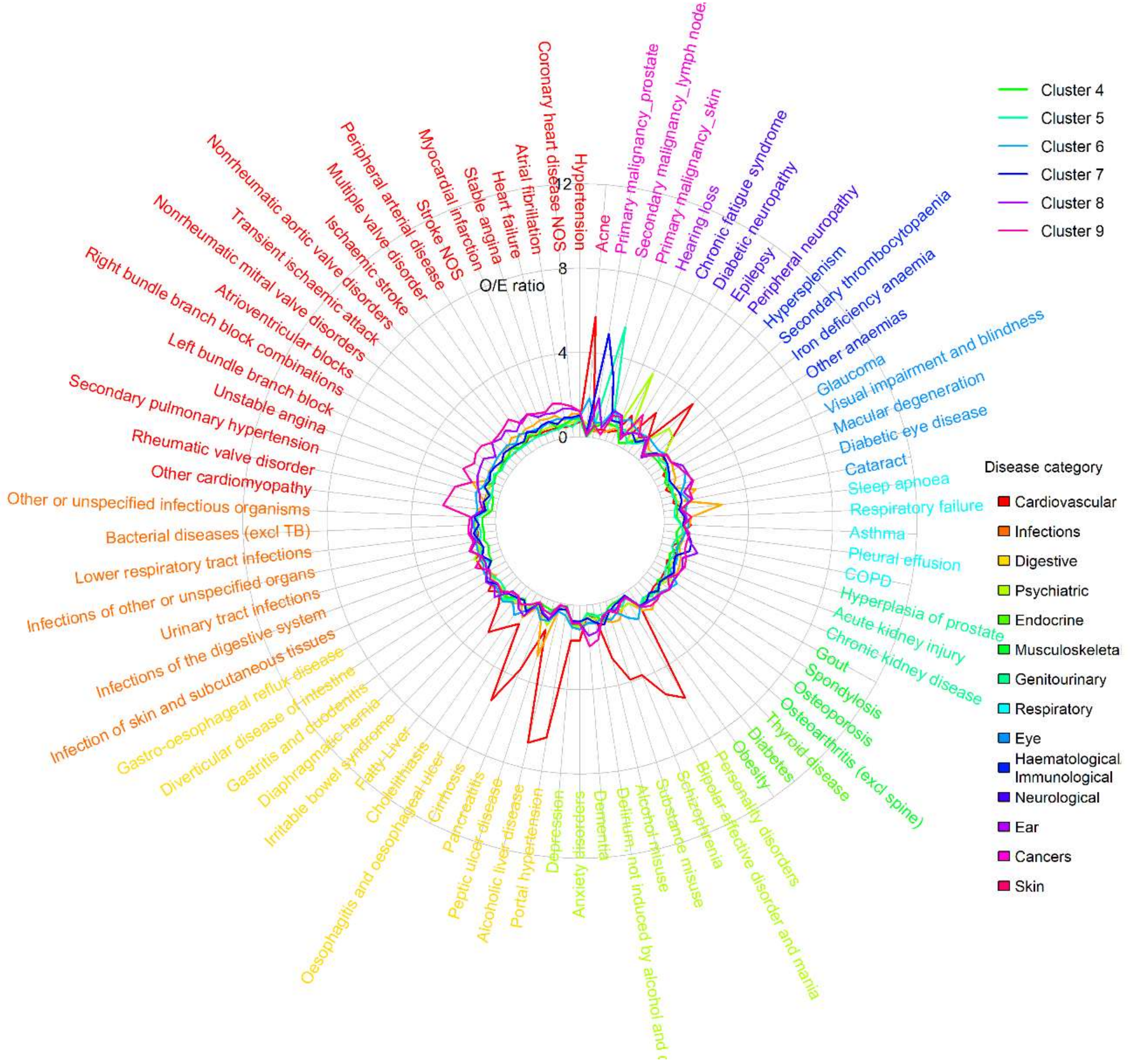

*Note: Relative prevalence is estimated according to relative prevalence to other patients in high-cost patient group*

**Table S4 – Descriptive statistics of high-cost clusters, based on 50% random sample (N=52,175; optimal hierarchical 9-cluster solution).**

| | (1) Liver disease & mental health | | (2) Sleep apnoea, gastro-reflux and diabetes | | (3) Chronic fatigue, immunological and haematological disorders | | (4) Low multimorbidity | | (5) Nodal metastases | | (6) Complex multimorbidity | | (7) Prostate cancer (with nodal metastasis) | | (8) Prostate, stroke & sensory impairment | | (9) Cardiovascular disease and dementia | |
|---|---|---|---|---|---|---|---|---|---|---|---|---|---|---|---|---|---|---|
| | N=6,523 (12.5%) | | N=4,589 (8.8%) | | N=5,464 (10.5%) | | N=6,852 (13.1%) | | N=3,514 (6.7%) | | N=2,842 (5.4%) | | N=3,850 (7.4%) | | N=6,138 (11.8%) | | N=12,403 (23.8%) | |
| Age | 55.8 | [18.2] | 65.6 | [18.1] | 65.6 | [16.6] | 66.2 | [15.4] | 67.3 | [14.7] | 68.3 | [19.3] | 72.8 | [11.4] | 78.9 | [11.1] | 79.2 | [12.5] |
| 333 | 3,221 | [49.4%] | 2,213 | [48.2%] | 2,871 | [52.5%] | 3,020 | [44.1%] | 2,371 | [67.5%] | 1,905 | [67.0%] | 1,205 | [31.3%] | 2,246 | [36.6%] | 6,294 | [50.7%] |
| Ethnicity | | | | | | | | | | | | | | | | | | |
| White | 5,818 | [89.2%] | 2,616 | [57.0%] | 3,998 | [73.2%] | 6,697 | [97.7%] | 3,185 | [90.6%] | 2,403 | [84.6%] | 3,615 | [93.9%] | 5,787 | [94.3%] | 11,261 | [90.8%] |
| Mixed | 0 | [0.0%] | 240 | [5.2%] | 0 | [0.0%] | 0 | [0.0%] | 0 | [0.0%] | 1 | [0.0%] | 1 | [0.0%] | 0 | [0.0%] | 0 | [0.0%] |
| Asian | 349 | [5.4%] | 932 | [20.3%] | 203 | [3.7%] | 94 | [1.4%] | 133 | [3.8%] | 144 | [5.1%] | 81 | [2.1%] | 179 | [2.9%] | 349 | [2.8%] |
| Black | 209 | [3.2%] | 754 | [16.4%] | 134 | [2.5%] | 42 | [0.6%] | 108 | [3.1%] | 174 | [6.1%] | 122 | [3.2%] | 126 | [2.1%] | 202 | [1.6%] |
| Other | 9 | [0.1%] | 0 | [0.0%] | 0 | [0.0%] | 0 | [0.0%] | 2 | [0.1%] | 4 | [0.1%] | 0 | [0.0%] | 0 | [0.0%] | 520 | [4.2%] |
| unknown | 138 | [2.1%] | 47 | [1.0%] | 1,129 | [20.7%] | 19 | [0.3%] | 86 | [2.4%] | 116 | [4.1%] | 31 | [0.8%] | 46 | [0.7%] | 71 | [0.6%] |
| IMD quintiles | | | | | | | | | | | | | | | | | | |
| 1 (least deprived) | 979 | [15.0%] | 566 | [12.3%] | 1,061 | [19.4%] | 1,446 | [21.1%] | 749 | [21.3%] | 578 | [20.3%] | 810 | [21.0%] | 1,302 | [21.2%] | 2,397 | [19.3%] |
| 2 | 1,057 | [16.2%] | 681 | [14.8%] | 1,054 | [19.3%] | 1,538 | [22.4%] | 730 | [20.8%] | 573 | [20.2%] | 742 | [19.3%] | 1,303 | [21.2%] | 2,602 | [21.0%] |
| 3 | 1,168 | [17.9%] | 900 | [19.6%] | 1,108 | [20.3%] | 1,408 | [20.5%] | 757 | [21.5%] | 544 | [19.1%] | 722 | [18.8%] | 1,203 | [19.6%] | 2,391 | [19.3%] |
| 4 | 1,456 | [22.3%] | 1,190 | [25.9%] | 1,140 | [20.9%] | 1,268 | [18.5%] | 650 | [18.5%] | 545 | [19.2%] | 742 | [19.3%] | 1,190 | [19.4%] | 2,451 | [19.8%] |
| 5 (most deprived) | 1,863 | [28.6%] | 1,252 | [27.3%] | 1,101 | [20.2%] | 1,192 | [17.4%] | 628 | [17.9%] | 602 | [21.2%] | 755 | [19.6%] | 1,140 | [18.6%] | 2,562 | [20.7%] |
| unknown | 0 | [0.0%] | 0 | [0.0%] | 0 | [0.0%] | 0 | [0.0%] | 0 | [0.0%] | 0 | [0.0%] | 79 | [2.1%] | 0 | [0.0%] | 0 | [0.0%] |
| Usage in 2018/19 | | | | | | | | | | | | | | | | | | |
| Total care costs (£) | 18,251 | [9,930] | 17,902 | [9,561] | 19,107 | [12,180] | 15,792 | [6,892] | 16,744 | [7,387] | 17,931 | [9,333] | 16,694 | [7,658] | 17,415 | [7,915] | 17,546 | [8,401] |
| Planned care (£) | 5,756 | [8,013] | 6,080 | [8,023] | 8,684 | [11,452] | 7,982 | [7,479] | 7,739 | [6,809] | 6,737 | [8,303] | 6,721 | [6,994] | 5,188 | [6,731] | 4,174 | [6,607] |
| Unplanned care (£) | 12,476 | [9,471] | 11,823 | [9,372] | 10,462 | [8,632] | 7,810 | [7,428] | 8,993 | [7,598] | 11,192 | [8,668] | 9,979 | [7,819] | 12,219 | [8,029] | 13,375 | [7,986] |
| Secondary care (£) | 17,733 | [9,926] | 17,391 | [9,555] | 18,637 | [12,224] | 15,350 | [6,886] | 16,229 | [7,360] | 17,323 | [9,346] | 16,141 | [7,653] | 16,813 | [7,914] | 16,940 | [8,380] |
| Primary care (£) | 518 | [546] | 511 | [500] | 470 | [455] | 441 | [415] | 515 | [469] | 608 | [545] | 553 | [481] | 602 | [551] | 606 | [595] |
| N(EM) | 3.8 | [3.8] | 2.9 | [2.9] | 2.6 | [2.4] | 1.9 | [2.0] | 2.5 | [2.4] | 3 | [2.9] | 2.6 | [2.2] | 3 | [2.2] | 3 | [2.4] |
| N(ED) | 7.3 | [14.9] | 4.3 | [9.1] | 3.3 | [4.9] | 2.7 | [4.8] | 2.9 | [3.6] | 4.8 | [9.8] | 3.2 | [4.3] | 3.6 | [3.7] | 3.9 | [5.0] |
| N(EL) | 3.4 | [10.9] | 5.2 | [20.2] | 5.9 | [13.2] | 3.4 | [8.5] | 6.7 | [10.9] | 4.2 | [13.4] | 4.1 | [8.8] | 2.4 | [9.1] | 1.7 | [9.2] |
| N(OP) | 12.5 | [15.3] | 13.6 | [17.0] | 15.4 | [16.9] | 13.5 | [14.3] | 21.5 | [19.1] | 13.7 | [15.6] | 16 | [15.1] | 11 | [14.0] | 9 | [15.2] |
| N(GP) | 17.5 | [18.5] | 16.5 | [15.1] | 15.9 | [14.7] | 15.6 | [13.4] | 16.6 | [15.1] | 19.4 | [16.1] | 17.9 | [14.2] | 18 | [15.0] | 18 | [19.1] |
| N(CALIBER) | 18.7 | [8.5] | 19.2 | [9.0] | 17.9 | [7.6] | 14.4 | [7.1] | 17.4 | [7.5] | 21.2 | [8.3] | 19.3 | [7.1] | 22 | [7.5] | 22 | [7.5] |
| N(LTC) | 14.9 | [7.3] | 15.5 | [7.5] | 14.1 | [6.8] | 12.2 | [6.1] | 14.3 | [6.3] | 16.3 | [7.2] | 15.8 | [6.1] | 18.1 | [6.5] | 17.3 | [6.6] |
| N(acute) | 3.8 | [2.7] | 3.7 | [2.9] | 3.9 | [2.8] | 2.3 | [2.4] | 3 | [2.5] | 4.9 | [2.8] | 3.5 | [2.4] | 4 | [2.6] | 4 | [2.5] |
| Disease system | | | | | | | | | | | | | | | | | | |
| Cancers | 1178.0 | [18.1%] | 659.0 | [14.4%] | 1954.0 | [35.8%] | 1613.0 | [23.5%] | 3,251 | [92.5%] | 980 | [34.5%] | 2,913 | [75.7%] | 2,582 | [42.1%] | 2,721 | [21.9%] |
| Circulatory | 4495.0 | [68.9%] | 3942.0 | [85.9%] | 4295.0 | [78.6%] | 5251.0 | [76.6%] | 2,545 | [72.4%] | 2,338 | [82.3%] | 3,365 | [87.4%] | 5,913 | [96.3%] | 11,949 | [96.3%] |
| Digestive | 5137.0 | [78.8%] | 2921.0 | [63.7%] | 3569.0 | [65.3%] | 3947.0 | [57.6%] | 2,074 | [59.0%] | 2,004 | [70.5%] | 2,579 | [67.0%] | 4,322 | [70.4%] | 7,720 | [62.2%] |
| Ear conditions | 1103.0 | [16.9%] | 950.0 | [20.7%] | 1197.0 | [21.9%] | 1611.0 | [23.5%] | 689 | [19.6%] | 819 | [28.8%] | 1,045 | [27.1%] | 2,108 | [34.3%] | 3,951 | [31.9%] |
| Endocrine | 3938.0 | [60.4%] | 3493.0 | [76.1%] | 3224.0 | [59.0%] | 4104.0 | [59.9%] | 2,010 | [57.2%] | 1,770 | [62.3%] | 2,243 | [58.3%] | 3,905 | [63.6%] | 8,262 | [66.6%] |
| Eye conditions | 1,686 | [25.8%] | 2,110 | [46.0%] | 1,720 | [31.5%] | 1,947 | [28.4%] | 1,020 | [29.0%] | 1,533 | [53.9%] | 1,495 | [38.8%] | 3,591 | [58.5%] | 7,135 | [57.5%] |
| Genitourinary | 3,845 | [58.9%] | 3,188 | [69.5%] | 3,470 | [63.5%] | 3,371 | [49.2%] | 1,957 | [55.7%] | 2,385 | [83.9%] | 2,405 | [62.5%] | 4,852 | [79.0%] | 9,981 | [80.5%] |
| Respiratory | 3,887 | [59.6%] | 2,854 | [62.2%] | 3,200 | [58.6%] | 3,078 | [44.9%] | 1,713 | [48.7%] | 1,738 | [61.2%] | 2,224 | [57.8%] | 3,954 | [64.4%] | 7,611 | [61.4%] |
| Haematological/Immunological | 3,317 | [50.9%] | 2,626 | [57.2%] | 2,963 | [54.2%] | 2,327 | [34.0%] | 1,633 | [46.5%] | 1,575 | [55.4%] | 1,871 | [48.6%] | 3,317 | [54.0%] | 6,664 | [53.7%] |
| Infectious Disease | 4,419 | [67.7%] | 3,090 | [67.3%] | 3,889 | [71.2%] | 3,298 | [48.1%] | 2,201 | [62.6%] | 2,013 | [70.8%] | 2,602 | [67.6%] | 4,669 | [76.1%] | 10,075 | [81.2%] |
| Mental Health | 5,119 | [78.5%] | 2,649 | [57.7%] | 2,975 | [54.4%] | 3,312 | [48.3%] | 1,620 | [46.1%] | 1,708 | [60.1%] | 1,925 | [50.0%] | 3,647 | [59.4%] | 8,185 | [66.0%] |

| | | | | | | | | | | | | | | | | | | |
|---|---|---|---|---|---|---|---|---|---|---|---|---|---|---|---|---|---|---|
| Musculoskeletal | 3,770 | [57.8%] | 3,178 | [69.3%] | 3,707 | [67.8%] | 4,907 | [71.6%] | 2,291 | [65.2%] | 2,286 | [80.4%] | 2,984 | [77.5%] | 4,881 | [79.5%] | 9,663 | [77.9%] |
| Neurological | 2,254 | [34.6%] | 1,134 | [24.7%] | 1,515 | [27.7%] | 1,383 | [20.2%] | 678 | [19.3%] | 861 | [30.3%] | 770 | [20.0%] | 1,427 | [23.2%] | 3,524 | [28.4%] |
| Skin conditions | 2,455 | [37.6%] | 1,294 | [28.2%] | 1,659 | [30.4%] | 2,456 | [35.8%] | 925 | [26.3%] | 1,347 | [47.4%] | 1,078 | [28.0%] | 1,815 | [29.6%] | 3,429 | [27.6%] |
| In-year Mortality | 882 | [13.5%] | 704 | [15.3%] | 929 | [17.0%] | 715 | [10.4%] | 1,128 | [32.1%] | 475 | [16.7%] | 1,201 | [31.2%] | 1,568 | [25.5%] | 3,157 | [25.5%] |

<u>Relative Prevalence for Top 20 Diseases</u>

| (1) | | (2) | | (3) | | (4) | | (5) | |
|---|---|---|---|---|---|---|---|---|---|
| Alcoholic liver disease | 6.8 | Sleep apnoea | 2.8 | Chronic fatigue syndrome | 3.8 | Obesity | 1.1 | Secondary malignancy_lymph nodes | 5.4 |
| Portal hypertension | 6.4 | Peptic ulcer disease | 2.7 | Hypersplenism | 2.1 | Irritable bowel syndrome | 1.0 | Primary malignancy_skin | 1.4 |
| Personality disorders | 5.8 | Diabetic eye disease | 1.7 | Secondary thrombocytopaenia | 1.9 | Osteoarthritis (excl spine) | 1.0 | Irritable bowel syndrome | 1.2 |
| Acne | 5.7 | Diabetic neuropathy | 1.5 | Infections of the digestive system | 1.3 | Gastro-oesophageal reflux disease | 1.0 | Infections of the digestive system | 1.0 |
| Cirrhosis | 5.5 | Diabetes | 1.4 | Irritable bowel syndrome | 1.3 | Spondylosis | 1.0 | Thyroid disease | 0.9 |
| Bipolar affective disorder and mania | 5.2 | Unstable angina | 1.4 | Peptic ulcer disease | 1.2 | Oesophagitis and oesophageal ulcer | 1.0 | Obesity | 0.9 |
| Substance misuse | 3.9 | Oesophagitis and oesophageal ulcer | 1.3 | Infections of other or unspecified organs | 1.2 | Stable angina | 0.9 | Anxiety disorders | 0.9 |
| Schizophrenia | 3.9 | Respiratory failure | 1.3 | Spondylosis | 1.2 | Anxiety disorders | 0.9 | Gastro-oesophageal reflux disease | 0.9 |
| Hypersplenism | 3.7 | Gout | 1.2 | Gastro-oesophageal reflux disease | 1.0 | Depression | 0.9 | Pleural effusion | 0.9 |
| Pancreatitis | 3.6 | Iron deficiency anaemia | 1.2 | Other or unspecified infectious organisms | 1.0 | Alcohol misuse | 0.9 | Other anaemias | 0.9 |
| Fatty Liver | 2.8 | Gastritis and duodenitis | 1.2 | Anxiety disorders | 1.0 | Unstable angina | 0.9 | Diverticular disease of intestine | 0.9 |
| Alcohol misuse | 2.7 | Asthma | 1.2 | Asthma | 1.0 | Hyperplasia of prostate | 0.9 | Other or unspecified infectious organisms | 0.9 |
| Epilepsy | 2.3 | Other anaemias | 1.2 | Depression | 1.0 | Hypertension | 0.9 | Secondary thrombocytopaenia | 0.9 |
| Cholelithiasis | 2.2 | Obesity | 1.2 | Bacterial diseases (excl TB) | 1.0 | Diverticular disease of intestine | 0.9 | Peripheral neuropathy | 0.9 |
| Secondary thrombocytopaenia | 1.8 | Chronic kidney disease | 1.2 | Sleep apnoea | 1.0 | Coronary heart disease NOS | 0.8 | Infections of other or unspecified organs | 0.8 |
| Depression | 1.7 | Myocardial infarction | 1.1 | Obesity | 1.0 | Myocardial infarction | 0.8 | Osteoarthritis (excl spine) | 0.8 |
| Anxiety disorders | 1.7 | Heart failure | 1.1 | Gastritis and duodenitis | 1.0 | Asthma | 0.8 | Chronic fatigue syndrome | 0.8 |
| Oesophagitis and oesophageal ulcer | 1.7 | Ischaemic stroke | 1.1 | Acute kidney injury | 0.9 | Primary malignancy_skin | 0.8 | Diaphragmatic hernia | 0.8 |
| Irritable bowel syndrome | 1.6 | Peripheral neuropathy | 1.1 | Diverticular disease of intestine | 0.9 | Hearing loss | 0.8 | Urinary tract infections | 0.8 |
| Chronic fatigue syndrome | 1.6 | Thyroid disease | 1.1 | Other anaemias | 0.9 | Diaphragmatic hernia | 0.8 | Osteoporosis | 0.8 |

| **(6)** | | **(7)** | | **(8)** | | **(9)** | |
|---|---|---|---|---|---|---|---|
| Acne | 1.8 | Primary malignancy_prostate | 5.0 | Primary malignancy_prostate | 1.9 | Other cardiomyopathy | 2.5 |
| Primary malignancy_skin | 1.5 | Secondary malignancy_lymph nodes | 3.0 | Hyperplasia of prostate | 1.8 | Rheumatic valve disorder | 2.3 |
| Spondylosis | 1.5 | Hyperplasia of prostate | 1.4 | Stroke NOS | 1.7 | Secondary pulmonary hypertension | 2.1 |
| Visual impairment and blindness | 1.5 | COPD | 1.4 | Macular degeneration | 1.7 | Left bundle branch block | 2.1 |
| Cholelithiasis | 1.4 | Diverticular disease of intestine | 1.3 | Visual impairment and blindness | 1.6 | Dementia | 2.0 |
| Personality disorders | 1.4 | Pleural effusion | 1.1 | Transient ischaemic attack | 1.6 | Multiple valve disorder | 1.9 |
| Chronic fatigue syndrome | 1.4 | Lower respiratory tract infections | 1.1 | Ischaemic stroke | 1.5 | Atrioventricular blocks | 1.9 |
| Glaucoma | 1.4 | Gout | 1.1 | Glaucoma | 1.5 | Nonrheumatic mitral valve disorders | 1.9 |
| Osteoporosis | 1.3 | Hearing loss | 1.0 | Peripheral arterial disease | 1.5 | Diabetic neuropathy | 1.9 |
| Irritable bowel syndrome | 1.3 | Alcohol misuse | 1.0 | Primary malignancy_skin | 1.5 | Heart failure | 1.7 |
| Macular degeneration | 1.3 | Other or unspecified infectious organisms | 1.0 | Dementia | 1.5 | Macular degeneration | 1.7 |
| Cataract | 1.2 | Gastro-oesophageal reflux disease | 1.0 | Atrioventricular blocks | 1.4 | Right bundle branch block combinations | 1.7 |
| Iron deficiency anaemia | 1.2 | Iron deficiency anaemia | 1.0 | Diaphragmatic hernia | 1.4 | Ischaemic stroke | 1.7 |
| Gastro-oesophageal reflux disease | 1.2 | Hypertension | 1.0 | Cataract | 1.4 | Delirium, not induced by alcohol and other psychoactive substances | 1.7 |
| Asthma | 1.2 | Oesophagitis and oesophageal ulcer | 1.0 | Nonrheumatic aortic valve disorders | 1.4 | Stroke NOS | 1.6 |
| Anxiety disorders | 1.1 | Peripheral neuropathy | 1.0 | Right bundle branch block combinations | 1.4 | Atrial fibrillation | 1.6 |
| Thyroid disease | 1.1 | Diaphragmatic hernia | 1.0 | Atrial fibrillation | 1.4 | Nonrheumatic aortic valve disorders | 1.6 |

| | | | | | | | |
|---|---|---|---|---|---|---|---|
| Other anaemias | 1.1 | Primary malignancy_skin | 1.0 | Hearing loss | 1.4 | Transient ischaemic attack | 1.6 |
| Diaphragmatic hernia | 1.1 | Respiratory failure | 1.0 | Urinary tract infections | 1.4 | Visual impairment and blindness | 1.5 |
| Diabetic neuropathy | 1.1 | Osteoarthritis (excl spine) | 1.0 | Osteoporosis | 1.4 | Myocardial infarction | 1.5 |

**Figure S3 – Disease-system profiles of age-stratified high-cost clusters**

Panel a. Patients aged 18-44 (fuzzy k-medoids, 6 clusters)

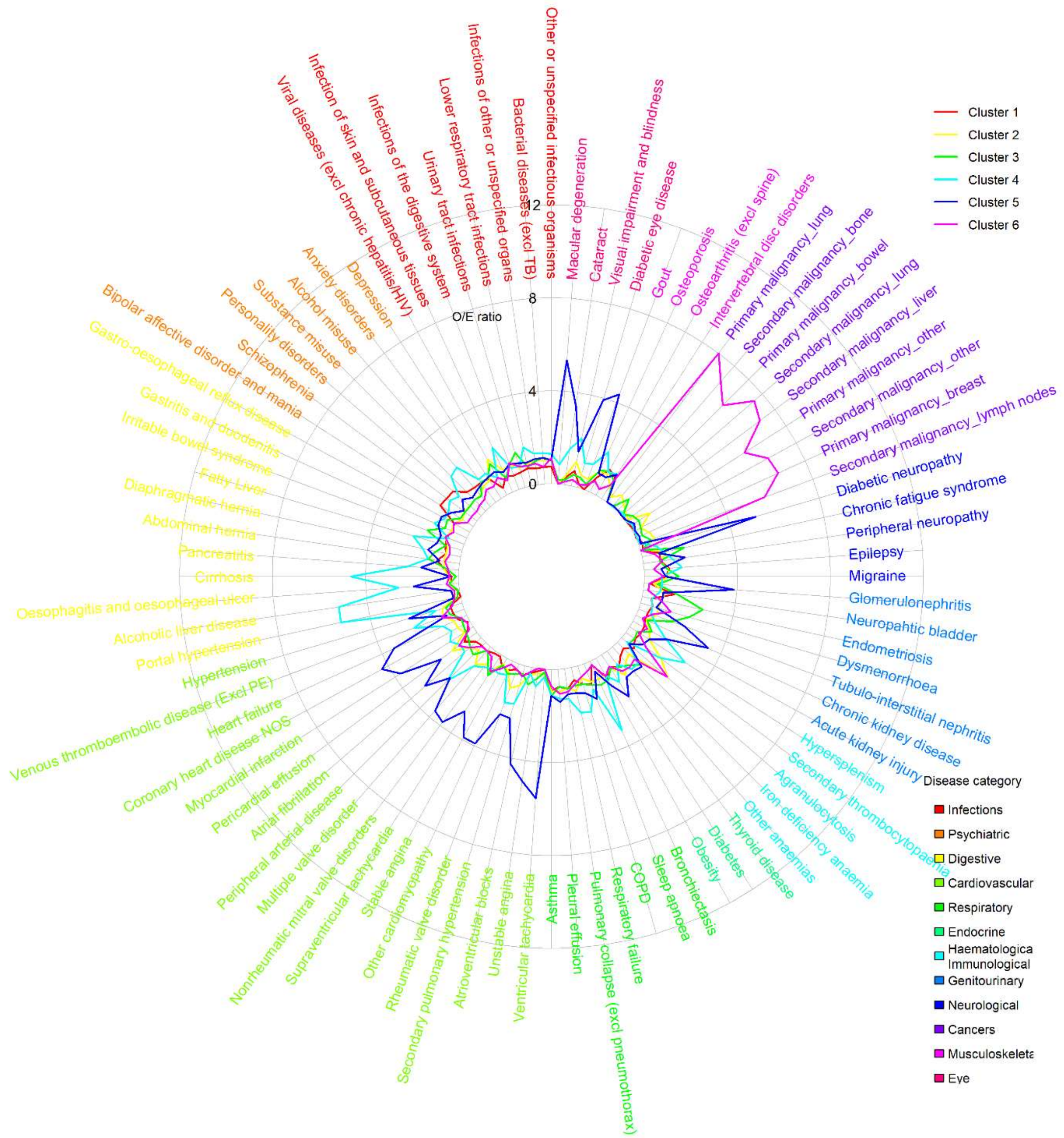

Panel b. Patients aged 45-65 (k-medoids, 6 clusters)

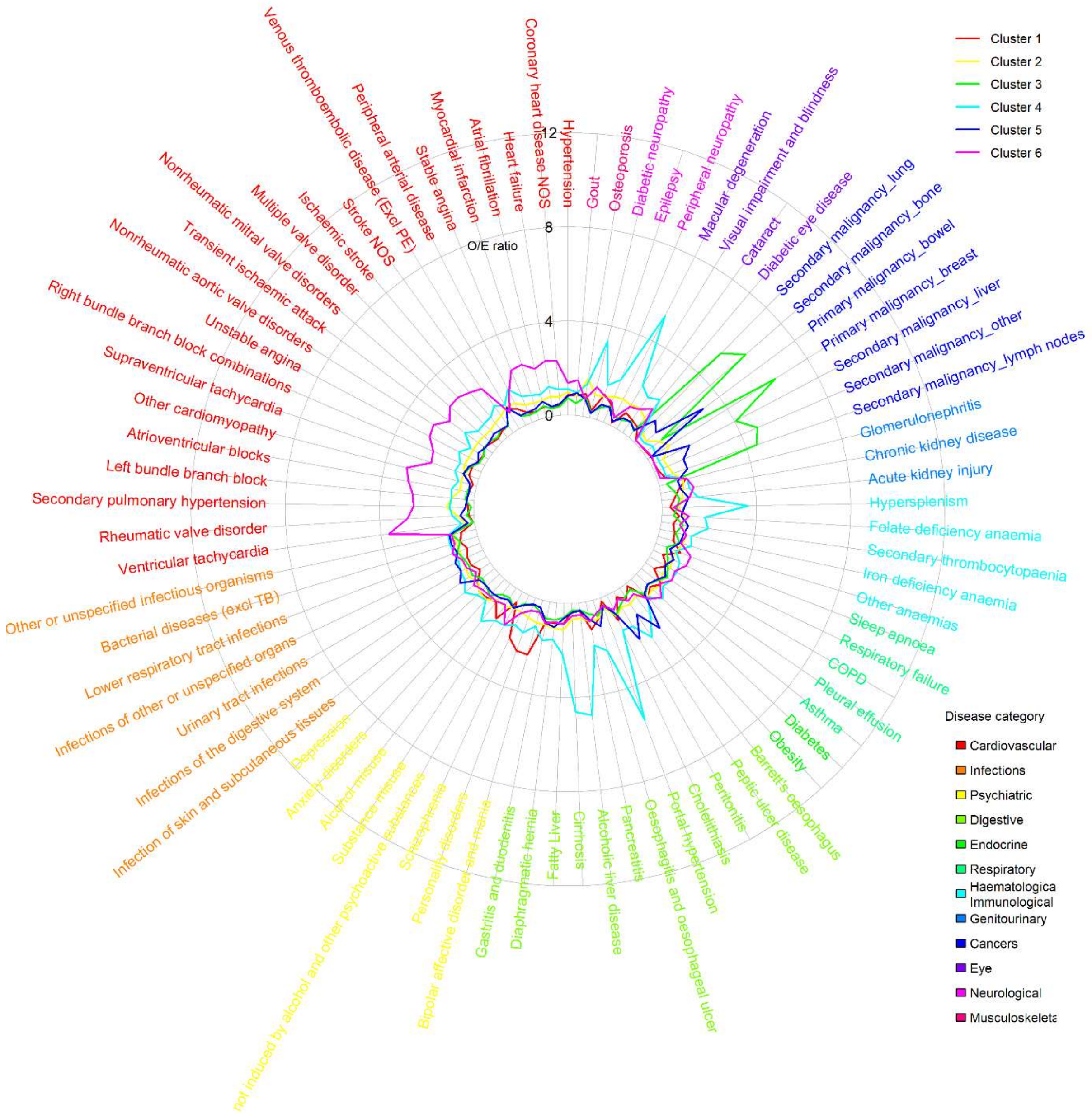

Panel c. Patients aged 66-79 (hierarchical, 4 clusters)

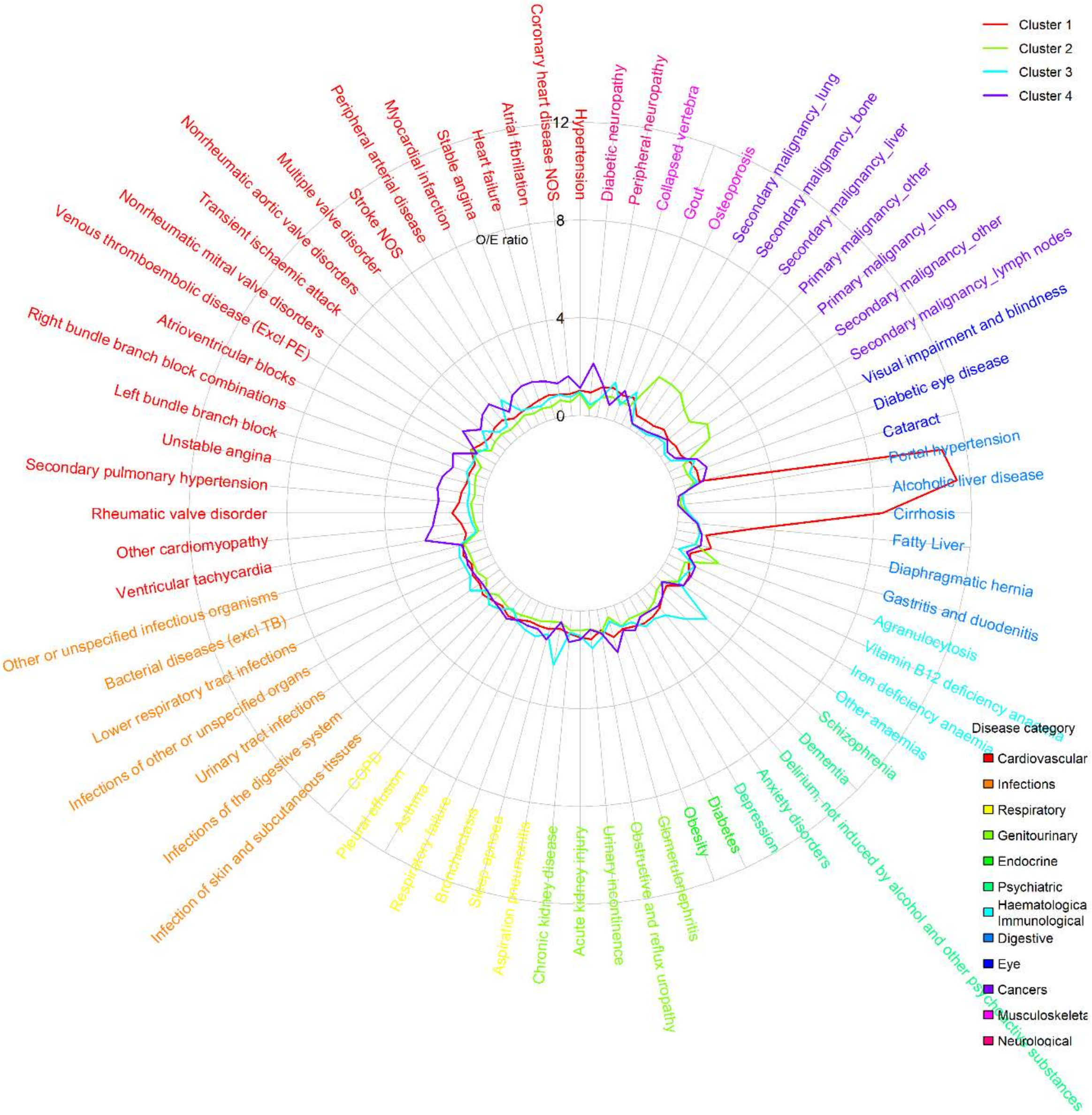

Panel d. Patients aged 80 or over (k-medoids, 6 clusters)

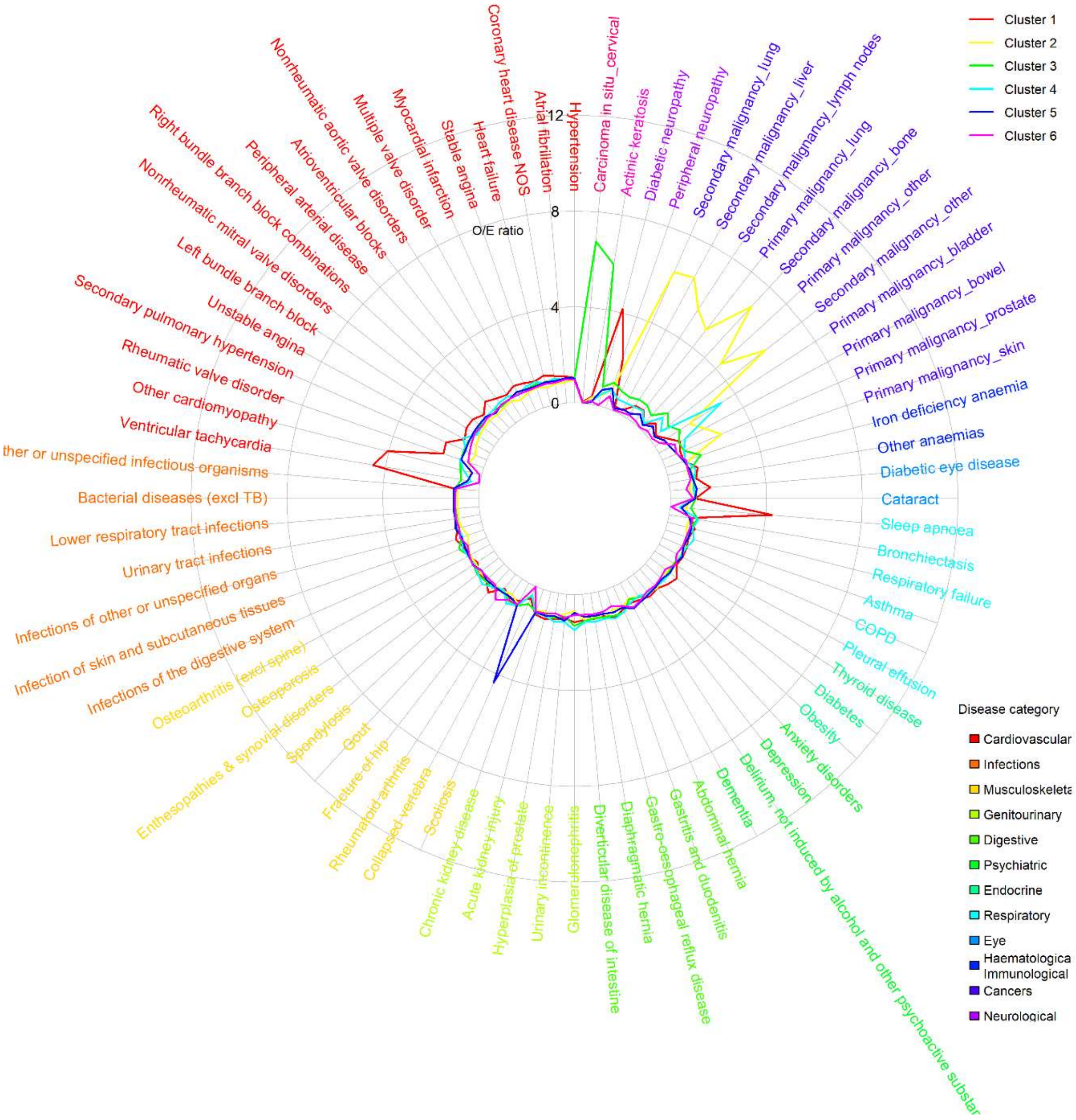

*Note: Relative prevalence is estimated according to relative prevalence to other patients in high-cost patient group*

### Table S5 – Descriptive statistics of age-stratified high-cost clusters, based on 50% random sample

a. Patients aged 18-44 (N=4,710; fuzzy k-medoids, 6 clusters)

| | (1) Mental health and substance misuse | | (2) Immunological and haematological disorders | | (3) Female pelvic pain, chronic fatigue and urogenital conditions | | (4) Alcohol-related liver disease | | (5) Cardiometabolic complications | | (6) Cancer | |
|---|---|---|---|---|---|---|---|---|---|---|---|---|
| | N=1,166 (24.8%) | | N=585 (12.4%) | | N=846 (18.0%) | | N=766 (16.3%) | | N=755 (16.0%) | | N=592 (12.6%) | |
| Age | 31.2 | [7.7] | 32.1 | [7.6] | 32.3 | [7.6] | 33.9 | [7.8] | 34.7 | [7.0] | 35.5 | [6.9] |
| Female | 543 | [46.6%] | 328 | [56.1%] | 563 | [66.5%] | 314 | [41.0%] | 378 | [50.1%] | 378 | [63.9%] |
| Ethnicity | | | | | | | | | | | | |
| White | 904 | [77.5%] | 344 | [58.8%] | 663 | [78.4%] | 627 | [81.9%] | 530 | [70.2%] | 370 | [62.5%] |
| Mixed | 0 | [0.0%] | 66 | [11.3%] | 5 | [0.6%] | 5 | [0.7%] | 9 | [1.2%] | 2 | [0.3%] |
| Asian | 110 | [9.4%] | 36 | [6.2%] | 68 | [8.0%] | 51 | [6.7%] | 99 | [13.1%] | 40 | [6.8%] |
| Black | 139 | [11.9%] | 28 | [4.8%] | 56 | [6.6%] | 37 | [4.8%] | 60 | [7.9%] | 34 | [5.7%] |
| Other | 0 | [0.0%] | 87 | [14.9%] | 3 | [0.4%] | 7 | [0.9%] | 5 | [0.7%] | 10 | [1.7%] |
| unknown | 13 | [1.1%] | 24 | [4.1%] | 51 | [6.0%] | 39 | [5.1%] | 52 | [6.9%] | 136 | [23.0%] |
| IMD quintiles | | | | | | | | | | | | |
| 1 (least deprived) | 174 | [14.9%] | 85 | [14.5%] | 118 | [13.9%] | 80 | [10.4%] | 75 | [9.9%] | 93 | [15.7%] |
| 2 | 159 | [13.6%] | 80 | [13.7%] | 138 | [16.3%] | 106 | [13.8%] | 119 | [15.8%] | 95 | [16.0%] |
| 3 | 223 | [19.1%] | 122 | [20.9%] | 149 | [17.6%] | 134 | [17.5%] | 157 | [20.8%] | 105 | [17.7%] |
| 4 | 285 | [24.4%] | 146 | [25.0%] | 197 | [23.3%] | 174 | [22.7%] | 181 | [24.0%] | 149 | [25.2%] |
| 5 (most deprived) | 325 | [27.9%] | 152 | [26.0%] | 244 | [28.8%] | 272 | [35.5%] | 223 | [29.5%] | 136 | [23.0%] |
| unknown | 0 | [0.0%] | 0 | [0.0%] | 0 | [0.0%] | 0 | [0.0%] | 0 | [0.0%] | 14 | [2.4%] |
| Usage in 2018/19 | | | | | | | | | | | | |
| Total care costs (£) | 18,073 | [11,942] | 21,813 | [14,446] | 17,330 | [10,077] | 19,940 | [11,826] | 19,091 | [11,007] | 18,351 | [12,245] |
| Planned care (£) | 8,130 | [10,969] | 11,047 | [13,776] | 7,324 | [7,967] | 5,827 | [8,086] | 7,895 | [9,918] | 9,674 | [9,241] |
| Unplanned care (£) | 9,827 | [9,701] | 10,817 | [10,451] | 9,984 | [9,460] | 14,113 | [11,672] | 11,198 | [9,501] | 8,766 | [10,433] |
| Secondary care (£) | 17,765 | [11,945] | 21,508 | [14,471] | 16,916 | [10,098] | 19,544 | [11,813] | 18,711 | [11,002] | 17,998 | [12,223] |
| Primary care (£) | 308 | [381] | 305 | [361] | 414 | [448] | 396 | [497] | 380 | [366] | 353 | [390] |
| N(EM) | 3.6 | [4.8] | 3 | [3.7] | 3.8 | [4.6] | 4.1 | [4.3] | 3.9 | [4.7] | 2.8 | [3.5] |
| N(ED) | 9 | [20.9] | 7.2 | [24.3] | 7.6 | [16.9] | 8.5 | [13.3] | 7.6 | [15.4] | 4.2 | [7.5] |
| N(EL) | 4.2 | [9.7] | 6 | [12.4] | 4.2 | [9.0] | 2.7 | [7.4] | 8.2 | [24.3] | 8.6 | [11.2] |
| N(OP) | 13.9 | [16.7] | 17.4 | [18.1] | 14.8 | [17.8] | 11.8 | [17.4] | 17.7 | [17.6] | 26.6 | [21.4] |
| N(GP) | 11.4 | [13.6] | 11.4 | [13.7] | 15.4 | [15.6] | 14.5 | [19.4] | 14.8 | [14.7] | 12.2 | [12.1] |
| N(CALIBER) | 8.2 | [5.6] | 10.8 | [6.3] | 11.7 | [5.9] | 14.8 | [7.6] | 15.1 | [7.5] | 10.4 | [6.2] |
| N(LTC) | 6.4 | [4.6] | 7.9 | [4.9] | 8.6 | [4.8] | 10.8 | [6.4] | 11.7 | [6.2] | 7.7 | [4.6] |
| N(acute) | 1.8 | [2.2] | 2.9 | [2.8] | 3.1 | [2.5] | 4 | [2.8] | 3.3 | [2.6] | 2.7 | [2.6] |
| Disease system | | | | | | | | | | | | |
| Cancers | 18 | [1.5%] | 130 | [22.2%] | 60 | [7.1%] | 16 | [2.1%] | 19 | [2.5%] | 434 | [73.3%] |
| Circulatory | 250 | [21.4%] | 259 | [44.3%] | 316 | [37.4%] | 416 | [54.3%] | 529 | [70.1%] | 172 | [29.1%] |
| Digestive | 457 | [39.2%] | 244 | [41.7%] | 560 | [66.2%] | 473 | [61.7%] | 453 | [60.0%] | 243 | [41.0%] |
| Ear conditions | 25 | [2.1%] | 173 | [29.6%] | 59 | [7.0%] | 61 | [8.0%] | 44 | [5.8%] | 37 | [6.2%] |
| Endocrine | 466 | [40.0%] | 230 | [39.3%] | 378 | [44.7%] | 332 | [43.3%] | 494 | [65.4%] | 232 | [39.2%] |
| Eye conditions | 44 | [3.8%] | 42 | [7.2%] | 39 | [4.6%] | 114 | [14.9%] | 199 | [26.4%] | 13 | [2.2%] |
| Genitourinary | 285 | [24.4%] | 195 | [33.3%] | 432 | [51.1%] | 384 | [50.1%] | 423 | [56.0%] | 176 | [29.7%] |
| Respiratory | 524 | [44.9%] | 310 | [53.0%] | 455 | [53.8%] | 471 | [61.5%] | 422 | [55.9%] | 267 | [45.1%] |
| Haematological/Immunological | 268 | [23.0%] | 250 | [42.7%] | 360 | [42.6%] | 358 | [46.7%] | 333 | [44.1%] | 277 | [46.8%] |
| Infectious Disease | 517 | [44.3%] | 339 | [57.9%] | 519 | [61.3%] | 524 | [68.4%] | 448 | [59.3%] | 335 | [56.6%] |
| Mental Health | 719 | [61.7%] | 324 | [55.4%] | 523 | [61.8%] | 629 | [82.1%] | 452 | [59.9%] | 249 | [42.1%] |
| Musculoskeletal | 311 | [26.7%] | 173 | [29.6%] | 261 | [30.9%] | 351 | [45.8%] | 289 | [38.3%] | 130 | [22.0%] |

| | | | | | | | | | | | | |
|---|---|---|---|---|---|---|---|---|---|---|---|---|
| Neurological | 333 | [28.6%] | 177 | [30.3%] | 302 | [35.7%] | 270 | [35.2%] | 269 | [35.6%] | 137 | [23.1%] |
| Skin conditions | 369 | [31.6%] | 216 | [36.9%] | 386 | [45.6%] | 230 | [30.0%] | 243 | [32.2%] | 183 | [30.9%] |
| In-year Mortality | 26 | [2.2%] | 38 | [6.5%] | 30 | [3.5%] | 67 | [8.7%] | 29 | [3.8%] | 111 | [18.8%] |

Relative Prevalence for Top 20 Diseases

| (1) | | (2) | | (3) | |
|---|---|---|---|---|---|
| Bipolar affective disorder and mania | 1.7 | Agranulocytosis | 2.6 | Endometriosis | 2.7 |
| Schizophrenia | 1.6 | Secondary thrombocytopaenia | 1.9 | Dysmenorrhoea | 2.2 |
| Personality disorders | 1.6 | Viral diseases (excl chronic hepatitis/HIV) | 1.7 | Chronic fatigue syndrome | 1.9 |
| Neuropahtic bladder | 1.3 | Pericardial effusion | 1.3 | Irritable bowel syndrome | 1.7 |
| Osteoarthritis (excl spine) | 1.2 | Thyroid disease | 1.3 | Urinary tract infections | 1.6 |
| Substance misuse | 1.1 | Rheumatic valv=e disorder | 1.1 | Neuropahtic bladder | 1.5 |
| Epilepsy | 1.1 | Infections of the digestive system | 1.1 | Viral diseases (excl chronic hepatitis/HIV) | 1.5 |
| Intervertebral disc disorders | 1.1 | Migraine | 1.1 | Tubulo-interstitial nephritis | 1.5 |
| Alcohol misuse | 1.1 | Infections of other or unspecified organs | 1.1 | Migraine | 1.5 |
| Anxiety disorders | 1.0 | Chronic fatigue syndrome | 1.1 | Iron deficiency anaemia | 1.4 |
| Depression | 1.0 | Infection of skin and subcutaneous tissues | 1.1 | Abdominal hernia | 1.4 |
| Obesity | 1.0 | Visual impairment and blindness | 1.1 | Osteoarthritis (excl spine) | 1.2 |
| Abdominal hernia | 1.0 | Respiratory failure | 1.0 | Bacterial diseases (excl TB) | 1.1 |
| Migraine | 0.9 | Other or unspecified infectious organisms | 1.0 | Infections of the digestive system | 1.1 |
| Sleep apnoea | 0.9 | Primary malignancy_other | 1.0 | Asthma | 1.1 |
| Asthma | 0.9 | Osteoarthritis (excl spine) | 1.0 | Bronchiectasis | 1.1 |
| Pulmonary collapse (excl pneumothorax) | 0.9 | Epilepsy | 1.0 | Diaphragmatic hernia | 1.1 |
| Thyroid disease | 0.9 | Bacterial diseases (excl TB) | 1.0 | Gastro-oesophageal reflux disease | 1.1 |
| COPD | 0.8 | Hypersplenism | 1.0 | Obesity | 1.1 |
| Irritable bowel syndrome | 0.8 | Multiple valve disorder | 1.0 | Anxiety disorders | 1.1 |

| (4) | | (5) | | (6) | |
|---|---|---|---|---|---|
| Portal hypertension | 5.3 | Ventricular tachycardia | 5.6 | Primary malignancy_lung | 8.0 |
| Alcoholic liver disease | 5.2 | Macular degeneration | 5.3 | Secondary malignancy_lung | 7.5 |
| Cirrhosis | 4.6 | Diabetic neuropathy | 5.2 | Secondary malignancy_liver | 7.2 |
| Bronchiectasis | 3.3 | Unstable angina | 4.9 | Secondary malignancy_bone | 7.1 |
| Hypersplenism | 2.8 | Gout | 4.4 | Primary malignancy_breast | 6.7 |
| Oesophagitis and oesophageal ulcer | 2.6 | Coronary heart disease NOS | 4.3 | Secondary malignancy_other | 6.6 |
| Peripheral arterial disease | 2.3 | Atrioventricular blocks | 4.3 | Primary malignancy_bowel | 6.4 |
| Venous thromboembolic disease (Excl PE) | 2.3 | Diabetic eye disease | 3.9 | Primary malignancy_other | 5.9 |
| Alcohol misuse | 2.2 | Other cardiomyopathy | 3.9 | Secondary malignancy_lymph nodes | 5.8 |
| Pancreatitis | 2.2 | Stable angina | 3.9 | Agranulocytosis | 2.6 |
| Fatty Liver | 2.2 | Glomerulonephritis | 3.9 | Dysmenorrhoea | 1.4 |
| Infection of skin and subcutaneous tissues | 2.1 | Nonrheumatic mitral valve disorders | 3.8 | Infections of the digestive system | 1.2 |
| Substance misuse | 2.1 | Myocardial infarction | 3.7 | Other anaemias | 1.1 |
| Visual impairment and blindness | 2.1 | Multiple valve disorder | 3.6 | Other or unspecified infectious organisms | 1.1 |
| COPD | 2.1 | Heart failure | 3.5 | Thyroid disease | 1.1 |
| Respiratory failure | 2.0 | Cataract | 3.4 | Secondary thrombocytopaenia | 1.1 |
| Osteoporosis | 1.9 | Chronic kidney disease | 3.4 | Pleural effusion | 1.0 |
| Secondary thrombocytopaenia | 1.8 | Atrial fibrillation | 3.2 | Iron deficiency anaemia | 1.0 |
| Rheumatic valve disorder | 1.8 | Supraventricular tachycardia | 2.9 | Venous thromboembolic disease (Excl PE) | 1.0 |
| Secondary pulmonary hypertension | 1.7 | Hypertension | 2.4 | Pulmonary collapse (excl pneumothorax) | 1.0 |

b. Patients aged 45-65 (N=12,586; k-medoids, 6 clusters)

| | (1) | | (2) | | (3) | | (4) | | (5) | | (6) | |
|---|---|---|---|---|---|---|---|---|---|---|---|---|
| | Mental health and substance misuse | | Sensory impairment with osteoporosis | | Breast-cancer dominant with widespread secondaries | | Alcohol-related liver disease | | Gastrointestinal cancers | | Cardiovascular disease | |
| | N=2,434 (19.3%) | | N=1,769 (14.1%) | | N=1,365 (10.8%) | | N=1,654 (13.1%) | | N=2,592 (20.6%) | | N=2,772 (22.0%) | |
| Age | 55.5 | [5.9] | 55.9 | [6.0] | 56.5 | [5.8] | 57 | [5.8] | 57.2 | [5.7] | 57.7 | [5.5] |
| Female | 1,207 | [49.6%] | 1,123 | [63.5%] | 959 | [70.3%] | 659 | [39.8%] | 906 | [35.0%] | 870 | [31.4%] |
| Ethnicity | | | | | | | | | | | | |
| White | 2,156 | [88.6%] | 1,312 | [74.2%] | 1,183 | [86.7%] | 1,375 | [83.1%] | 2,156 | [83.2%] | 2,185 | [78.8%] |
| Mixed | 0 | [0.0%] | 0 | [0.0%] | 0 | [0.0%] | 85 | [5.1%] | 0 | [0.0%] | 0 | [0.0%] |
| Asian | 83 | [3.4%] | 125 | [7.1%] | 59 | [4.3%] | 88 | [5.3%] | 106 | [4.1%] | 330 | [11.9%] |
| Black | 182 | [7.5%] | 110 | [6.2%] | 70 | [5.1%] | 59 | [3.6%] | 102 | [3.9%] | 139 | [5.0%] |
| Other | 0 | [0.0%] | 149 | [8.4%] | 1 | [0.1%] | 4 | [0.2%] | 2 | [0.1%] | 0 | [0.0%] |
| unknown | 13 | [0.5%] | 73 | [4.1%] | 52 | [3.8%] | 43 | [2.6%] | 226 | [8.7%] | 118 | [4.3%] |
| IMD quintiles | | | | | | | | | | | | |
| 1 (least deprived) | 402 | [16.5%] | 255 | [14.4%] | 228 | [16.7%] | 207 | [12.5%] | 469 | [18.1%] | 355 | [12.8%] |
| 2 | 377 | [15.5%] | 314 | [17.8%] | 284 | [20.8%] | 252 | [15.2%] | 554 | [21.4%] | 419 | [15.1%] |
| 3 | 500 | [20.5%] | 324 | [18.3%] | 261 | [19.1%] | 286 | [17.3%] | 483 | [18.6%] | 520 | [18.8%] |
| 4 | 568 | [23.3%] | 409 | [23.1%] | 296 | [21.7%] | 391 | [23.6%] | 512 | [19.8%] | 634 | [22.9%] |
| 5 (most deprived) | 587 | [24.1%] | 467 | [26.4%] | 296 | [21.7%] | 518 | [31.3%] | 574 | [22.1%] | 824 | [29.7%] |
| unknown | 0 | [0.0%] | 0 | [0.0%] | 0 | [0.0%] | 0 | [0.0%] | 0 | [0.0%] | 20 | [0.7%] |
| Usage in 2018/19 | | | | | | | | | | | | |
| Total care costs (£) | 16,761 | [9,014] | 18,233 | [10,769] | 16,903 | [7,119] | 19,332 | [10,365] | 19,193 | [11,616] | 18,676 | [10,994] |
| Planned care (£) | 8,689 | [8,422] | 8,411 | [9,081] | 8,552 | [6,298] | 6,456 | [8,635] | 9,800 | [11,121] | 7,474 | [9,586] |
| Unplanned care (£) | 8,075 | [8,896] | 9,827 | [9,854] | 8,339 | [7,458] | 12,892 | [9,761] | 9,406 | [8,478] | 11,218 | [9,845] |
| Secondary care (£) | 16,356 | [9,007] | 17,756 | [10,773] | 16,467 | [7,080] | 18,822 | [10,355] | 18,790 | [11,642] | 18,225 | [10,985] |
| Primary care (£) | 405 | [421] | 476 | [438] | 436 | [412] | 510 | [534] | 403 | [380] | 451 | [412] |
| N(EM) | 2.3 | [3.0] | 2.7 | [3.1] | 2.6 | [2.4] | 3.4 | [3.1] | 2.5 | [2.4] | 2.7 | [2.8] |
| N(ED) | 4.4 | [11.0] | 4.3 | [8.3] | 2.9 | [3.7] | 5.4 | [7.8] | 3.1 | [4.8] | 3.9 | [6.8] |
| N(EL) | 5.1 | [16.1] | 6.7 | [17.6] | 7.6 | [10.4] | 4.3 | [14.9] | 7.5 | [15.6] | 3.7 | [14.7] |
| N(OP) | 15.1 | [16.5] | 17.6 | [17.6] | 25.3 | [19.6] | 15.3 | [18.7] | 19.3 | [18.6] | 13.6 | [16.2] |
| N(GP) | 15 | [14.6] | 17.2 | [15.0] | 14.7 | [13.8] | 17.9 | [18.7] | 14.5 | [13.3] | 16.5 | [15.1] |
| N(CALIBER) | 13.3 | [7.1] | 17.8 | [7.7] | 15.6 | [7.1] | 21.6 | [8.2] | 15.2 | [6.6] | 18.8 | [7.7] |
| N(LTC) | 11.2 | [6.0] | 14.4 | [6.7] | 12.7 | [5.9] | 17.5 | [7.1] | 11.4 | [5.6] | 15.6 | [6.6] |
| N(acute) | 2.1 | [2.3] | 3.4 | [2.7] | 2.8 | [2.4] | 4.1 | [2.7] | 3.8 | [2.7] | 3.2 | [2.7] |
| Disease system | | | | | | | | | | | | |
| Cancers | 224 | [9.2%] | 421 | [23.8%] | 1,251 | [91.6%] | 275 | [16.6%] | 1,392 | [53.7%] | 231 | [8.3%] |
| Circulatory | 1,536 | [63.1%] | 1,304 | [73.7%] | 836 | [61.2%] | 1,329 | [80.4%] | 1,686 | [65.0%] | 2,665 | [96.1%] |
| Digestive | 1,433 | [58.9%] | 1,124 | [63.5%] | 739 | [54.1%] | 1,312 | [79.3%] | 1,744 | [67.3%] | 1,598 | [57.6%] |
| Ear conditions | 301 | [12.4%] | 258 | [14.6%] | 155 | [11.4%] | 254 | [15.4%] | 369 | [14.2%] | 347 | [12.5%] |
| Endocrine | 1,561 | [64.1%] | 1,177 | [66.5%] | 757 | [55.5%] | 1,154 | [69.8%] | 1,400 | [54.0%] | 2,010 | [72.5%] |
| Eye conditions | 389 | [16.0%] | 532 | [30.1%] | 169 | [12.4%] | 654 | [39.5%] | 333 | [12.8%] | 870 | [31.4%] |
| Genitourinary | 896 | [36.8%] | 978 | [55.3%] | 542 | [39.7%] | 1,070 | [64.7%] | 1,340 | [51.7%] | 1,443 | [52.1%] |
| Respiratory | 1,156 | [47.5%] | 1,046 | [59.1%] | 689 | [50.5%] | 1,031 | [62.3%] | 1,378 | [53.2%] | 1,777 | [64.1%] |
| Haematological/Immunological | 759 | [31.2%] | 854 | [48.3%] | 590 | [43.2%] | 1,007 | [60.9%] | 1,238 | [47.8%] | 1,206 | [43.5%] |
| Infectious Disease | 1,095 | [45.0%] | 1,085 | [61.3%] | 836 | [61.2%] | 1,176 | [71.1%] | 1,830 | [70.6%] | 1,748 | [63.1%] |
| Mental Health | 1,531 | [62.9%] | 1,130 | [63.9%] | 666 | [48.8%] | 1,282 | [77.5%] | 1,284 | [49.5%] | 1,723 | [62.2%] |
| Musculoskeletal | 1,639 | [67.3%] | 1,319 | [74.6%] | 803 | [58.8%] | 1,118 | [67.6%] | 1,333 | [51.4%] | 1,597 | [57.6%] |
| Neurological | 761 | [31.3%] | 648 | [36.6%] | 315 | [23.1%] | 831 | [50.2%] | 540 | [20.8%] | 807 | [29.1%] |
| Skin conditions | 646 | [26.5%] | 739 | [41.8%] | 354 | [25.9%] | 549 | [33.2%] | 626 | [24.2%] | 712 | [25.7%] |
| In-year Mortality | 106 | [4.4%] | 166 | [9.4%] | 463 | [33.9%] | 290 | [17.5%] | 458 | [17.7%] | 333 | [12.0%] |

Relative Prevalence for Top 20 Diseases

| (1) | | (2) | | (3) | |
|---|---|---|---|---|---|
| Bipolar affective disorder and mania | 2.4 | Osteoporosis | 1.6 | Primary malignancy_breast | 6.4 |
| Personality disorders | 2.4 | Cataract | 1.3 | Secondary malignancy_bone | 6.0 |
| Schizophrenia | 2.0 | Macular degeneration | 1.2 | Secondary malignancy_lung | 5.3 |
| Substance misuse | 1.5 | Visual impairment and blindness | 1.2 | Secondary malignancy_other | 4.8 |
| Pancreatitis | 1.2 | Anxiety disorders | 1.2 | Secondary malignancy_lymph nodes | 4.5 |
| Cholelithiasis | 1.2 | Peripheral neuropathy | 1.2 | Secondary malignancy_liver | 4.1 |
| Glomerulonephritis | 1.2 | Asthma | 1.2 | Pleural effusion | 1.1 |
| Depression | 1.1 | Iron deficiency anaemia | 1.1 | Venous thromboembolic disease (Excl PE) | 1.1 |
| Epilepsy | 1.1 | Secondary pulmonary hypertension | 1.1 | Other or unspecified infectious organisms | 1.1 |
| Sleep apnoea | 1.1 | Depression | 1.1 | Lower respiratory tract infections | 1.0 |
| Anxiety disorders | 1.1 | Fatty Liver | 1.1 | Primary malignancy_bowel | 1.0 |
| Asthma | 1.1 | Diaphragmatic hernia | 1.1 | Other anaemias | 0.9 |
| Obesity | 1.1 | Other anaemias | 1.1 | Infections of the digestive system | 0.9 |
| Venous thromboembolic disease (Excl PE) | 1.0 | Primary malignancy_breast | 1.1 | Cholelithiasis | 0.9 |
| Alcohol misuse | 1.0 | Infections of the digestive system | 1.1 | Secondary thrombocytopaenia | 0.9 |
| Osteoporosis | 1.0 | Venous thromboembolic disease (Excl PE) | 1.1 | Anxiety disorders | 0.9 |
| Infection of skin and subcutaneous tissues | 0.9 | Glomerulonephritis | 1.1 | COPD | 0.9 |
| Gastritis and duodenitis | 0.9 | Diabetic eye disease | 1.1 | Infections of other or unspecified organs | 0.9 |
| Diaphragmatic hernia | 0.9 | Obesity | 1.0 | Obesity | 0.9 |
| Hypertension | 0.9 | Peripheral arterial disease | 1.0 | Osteoporosis | 0.8 |
| (4) | | (5) | | (6) | |
| Portal hypertension | 5.5 | Primary malignancy_bowel | 3.1 | Ventricular tachycardia | 3.7 |
| Macular degeneration | 5.2 | Barrett's oesophagus | 2.4 | Other cardiomyopathy | 3.1 |
| Alcoholic liver disease | 4.8 | Peritonitis | 2.3 | Atrioventricular blocks | 2.9 |
| Cirrhosis | 4.6 | Secondary malignancy_liver | 1.8 | Rheumatic valve disorder | 2.8 |
| Hypersplenism | 3.6 | Infections of the digestive system | 1.5 | Nonrheumatic aortic valve disorders | 2.7 |
| Diabetic neuropathy | 3.3 | Secondary malignancy_lymph nodes | 1.3 | Multiple valve disorder | 2.7 |
| Peptic ulcer disease | 2.5 | Secondary malignancy_other | 1.3 | Nonrheumatic mitral valve disorders | 2.6 |
| Oesophagitis and oesophageal ulcer | 2.2 | Peptic ulcer disease | 1.3 | Unstable angina | 2.6 |
| Fatty Liver | 2.2 | Infections of other or unspecified organs | 1.3 | Left bundle branch block | 2.6 |
| Visual impairment and blindness | 2.1 | Secondary malignancy_lung | 1.3 | Secondary pulmonary hypertension | 2.6 |
| Diabetic eye disease | 2.1 | Urinary tract infections | 1.2 | Ischaemic stroke | 2.5 |
| Alcohol misuse | 2.0 | Bacterial diseases (excl TB) | 1.2 | Myocardial infarction | 2.5 |
| Secondary thrombocytopaenia | 2.0 | Secondary thrombocytopaenia | 1.2 | Stable angina | 2.4 |
| Peripheral neuropathy | 1.9 | Other or unspecified infectious organisms | 1.2 | Heart failure | 2.4 |
| Pancreatitis | 1.9 | Oesophagitis and oesophageal ulcer | 1.1 | Coronary heart disease NOS | 2.3 |
| Cataract | 1.8 | Acute kidney injury | 1.1 | Stroke NOS | 2.3 |
| Folate deficiency anaemia | 1.8 | Venous thromboembolic disease (Excl PE) | 1.0 | Transient ischaemic attack | 2.3 |
| Barrett's oesophagus | 1.7 | Lower respiratory tract infections | 1.0 | Right bundle branch block combinations | 2.2 |
| Peripheral arterial disease | 1.7 | Diaphragmatic hernia | 1.0 | Atrial fibrillation | 2.1 |
| Peritonitis | 1.7 | Pleural effusion | 1.0 | Supraventricular tachycardia | 2.1 |

c. Patients aged 66-79 (N=17,130; hierarchical, 4 clusters)

| | (1) Alcohol-related liver disease N=1,330 (7.8%) | | (2) Nodal metastases N=5,686 (33.2%) | | (3) Neurodegenerative and frailty N=4,427 (25.8%) | | (4) Cardiovascular disease N=5,687 (33.2%) | |
|---|---|---|---|---|---|---|---|---|
| Age | 72.4 | [4.0] | 72.5 | [3.9] | 73.1 | [4.0] | 73.2 | [3.9] |
| Female | 665 | [50.0%] | 2,727 | [48.0%] | 2,202 | [49.7%] | 1,935 | [34.0%] |
| Ethnicity | | | | | | | | |
| White | 1,204 | [90.5%] | 5,340 | [93.9%] | 3,761 | [85.0%] | 5,027 | [88.4%] |
| Mixed | 0 | [0.0%] | 43 | [0.8%] | 0 | [0.0%] | 0 | [0.0%] |
| Asian | 62 | [4.7%] | 182 | [3.2%] | 120 | [2.7%] | 365 | [6.4%] |
| Black | 28 | [2.1%] | 69 | [1.2%] | 63 | [1.4%] | 260 | [4.6%] |
| Other | 6 | [0.5%] | 1 | [0.0%] | 137 | [3.1%] | 0 | [0.0%] |
| unknown | 30 | [2.3%] | 51 | [0.9%] | 346 | [7.8%] | 35 | [0.6%] |
| IMD quintiles | | | | | | | | |
| 1 (least deprived) | 281 | [21.1%] | 1,286 | [22.6%] | 756 | [17.1%] | 1,003 | [17.6%] |
| 2 | 265 | [19.9%] | 1,200 | [21.1%] | 830 | [18.7%] | 1,199 | [21.1%] |
| 3 | 251 | [18.9%] | 1,163 | [20.5%] | 851 | [19.2%] | 1,143 | [20.1%] |
| 4 | 277 | [20.8%] | 1,049 | [18.4%] | 946 | [21.4%] | 1,159 | [20.4%] |
| 5 (most deprived) | 239 | [18.0%] | 988 | [17.4%] | 1,044 | [23.6%] | 1,183 | [20.8%] |
| unknown | 17 | [1.3%] | 0 | [0.0%] | 0 | [0.0%] | 0 | [0.0%] |
| Usage in 2018/19 | | | | | | | | |
| Total care costs (£) | 17,684 | [8,989] | 16,817 | [7,993] | 17,904 | [8,904] | 17,863 | [9,197] |
| Planned care (£) | 6,799 | [7,663] | 7,968 | [7,896] | 5,216 | [7,490] | 7,163 | [7,861] |
| Unplanned care (£) | 10,894 | [8,638] | 8,854 | [7,495] | 12,683 | [8,598] | 10,721 | [8,997] |
| Secondary care (£) | 17,106 | [8,965] | 16,325 | [7,984] | 17,334 | [8,885] | 17,322 | [9,168] |
| Primary care (£) | 577 | [486] | 492 | [426] | 571 | [588] | 541 | [458] |
| N(EM) | 2.8 | [2.5] | 2.2 | [2.1] | 3 | [2.4] | 2.7 | [2.6] |
| N(ED) | 3.8 | [5.9] | 2.6 | [3.3] | 4.2 | [6.7] | 3.4 | [5.3] |
| N(EL) | 4.1 | [11.0] | 5.3 | [10.4] | 2 | [7.5] | 3.9 | [13.1] |
| N(OP) | 13.8 | [13.4] | 16.9 | [15.6] | 9.9 | [13.2] | 14.6 | [16.0] |
| N(GP) | 19.9 | [16.2] | 17.1 | [14.4] | 17.4 | [18.5] | 19.3 | [15.0] |
| N(CALIBER) | 23.1 | [7.9] | 17.7 | [6.9] | 19.8 | [7.6] | 22.2 | [7.9] |
| N(LTC) | 18.5 | [7.1] | 14.5 | [5.8] | 15.6 | [6.5] | 18.6 | [6.7] |
| N(acute) | 4.6 | [2.7] | 3.1 | [2.6] | 4.2 | [2.7] | 3.6 | [2.7] |
| Disease system | | | | | | | | |
| Cancers | 517 | [38.9%] | 3,494 | [61.4%] | 949 | [21.4%] | 1,783 | [31.4%] |
| Circulatory | 1,218 | [91.6%] | 4,820 | [84.8%] | 4,017 | [90.7%] | 5,509 | [96.9%] |
| Digestive | 1,130 | [85.0%] | 3,896 | [68.5%] | 2,764 | [62.4%] | 3,948 | [69.4%] |
| Ear conditions | 342 | [25.7%] | 1,317 | [23.2%] | 1,019 | [23.0%] | 1,443 | [25.4%] |
| Endocrine | 922 | [69.3%] | 3,501 | [61.6%] | 2,873 | [64.9%] | 4,225 | [74.3%] |
| Eye conditions | 584 | [43.9%] | 1,987 | [34.9%] | 1,727 | [39.0%] | 2,855 | [50.2%] |
| Genitourinary | 972 | [73.1%] | 3,466 | [61.0%] | 3,142 | [71.0%] | 4,273 | [75.1%] |
| Respiratory | 811 | [61.0%] | 2,957 | [52.0%] | 2,703 | [61.1%] | 3,711 | [65.3%] |
| Haematological/Immunological | 784 | [58.9%] | 2,725 | [47.9%] | 2,079 | [47.0%] | 3,095 | [54.4%] |
| Infectious Disease | 918 | [69.0%] | 3,554 | [62.5%] | 3,376 | [76.3%] | 3,877 | [68.2%] |
| Mental Health | 863 | [64.9%] | 2,459 | [43.2%] | 3,089 | [69.8%] | 2,778 | [48.8%] |
| Musculoskeletal | 1,084 | [81.5%] | 4,298 | [75.6%] | 3,417 | [77.2%] | 4,401 | [77.4%] |
| Neurological | 376 | [28.3%] | 927 | [16.3%] | 1,474 | [33.3%] | 1,608 | [28.3%] |
| Skin conditions | 682 | [51.3%] | 1,753 | [30.8%] | 1,303 | [29.4%] | 1,672 | [29.4%] |
| In-year Mortality | 265 | [19.9%] | 1,450 | [25.5%] | 817 | [18.5%] | 1,001 | [17.6%] |

Relative Prevalence for Top 20 Diseases

| (1) | | (2) | |
|---|---|---|---|
| Alcoholic liver disease | 11.5 | Secondary malignancy_bone | 2.4 |
| Portal hypertension | 11.0 | Secondary malignancy_lung | 2.4 |
| Cirrhosis | 8.3 | Secondary malignancy_liver | 2.4 |
| Fatty Liver | 3.2 | Secondary malignancy_other | 2.3 |
| Gastritis and duodenitis | 1.5 | Secondary malignancy_lymph nodes | 2.1 |
| Collapsed vertebra | 1.3 | Agranulocytosis | 2.0 |
| Glomerulonephritis | 1.3 | Primary malignancy_other | 2.0 |
| Peripheral neuropathy | 1.2 | Primary malignancy_lung | 1.8 |
| Rheumatic valve disorder | 1.2 | Venous thromboembolic disease (Excl PE) | 1.1 |
| Diaphragmatic hernia | 1.2 | Infections of the digestive system | 1.1 |
| Depression | 1.2 | Diaphragmatic hernia | 1.0 |
| Osteoporosis | 1.2 | Obstructive and reflux uropathy | 1.0 |
| Infections of the digestive system | 1.2 | Collapsed vertebra | 0.9 |
| Urinary incontinence | 1.2 | Obesity | 0.9 |
| Venous thromboembolic disease (Excl PE) | 1.2 | Other or unspecified infectious organisms | 0.9 |
| Anxiety disorders | 1.2 | Hypertension | 0.9 |
| Other anaemias | 1.1 | Other anaemias | 0.9 |
| Iron deficiency anaemia | 1.1 | Pleural effusion | 0.9 |
| Pleural effusion | 1.1 | Gastritis and duodenitis | 0.9 |
| Gout | 1.1 | Infections of other or unspecified organs | 0.9 |
| (3) | | (4) | |
| Schizophrenia | 2.8 | Ventricular tachycardia | 2.4 |
| Aspiration pneumonitis | 2.3 | Diabetic neuropathy | 2.2 |
| Dementia | 2.1 | Other cardiomyopathy | 2.1 |
| Stroke NOS | 1.7 | Unstable angina | 1.9 |
| Urinary incontinence | 1.5 | Rheumatic valve disorder | 1.9 |
| Collapsed vertebra | 1.5 | Glomerulonephritis | 1.9 |
| Urinary tract infections | 1.5 | Nonrheumatic mitral valve disorders | 1.9 |
| Osteoporosis | 1.5 | Left bundle branch block | 1.8 |
| Delirium, not induced by alcohol and other psychoactive substances | 1.4 | Multiple valve disorder | 1.8 |
| Depression | 1.4 | Secondary pulmonary hypertension | 1.8 |
| Bronchiectasis | 1.4 | Atrioventricular blocks | 1.8 |
| Respiratory failure | 1.3 | Myocardial infarction | 1.7 |
| Infection of skin and subcutaneous tissues | 1.3 | Stable angina | 1.7 |
| Anxiety disorders | 1.3 | Nonrheumatic aortic valve disorders | 1.7 |
| Bacterial diseases (excl TB) | 1.2 | Coronary heart disease NOS | 1.6 |
| Transient ischaemic attack | 1.2 | Heart failure | 1.6 |
| Infections of other or unspecified organs | 1.2 | Peripheral arterial disease | 1.6 |
| Lower respiratory tract infections | 1.2 | Right bundle branch block combinations | 1.5 |
| Obstructive and reflux uropathy | 1.1 | Diabetic eye disease | 1.5 |
| Visual impairment and blindness | 1.1 | Atrial fibrillation | 1.4 |

d. Patients aged 80 or over (N=17,752; k-medoids, 6 clusters)

| | (1) Cardiovascular disease N=2,754 (15.5%) | | (2) Systemic metastases N=2,037 (11.5%) | | (3) Cervical and skin cancers N=2,587 (14.6%) | | (4) Urogenital and gastrointestinal cancers N=1,970 (11.1%) | | (5) Scoliosis and dementia N=2,905 (16.4%) | | (6) Hip fracture and dementia N=5,499 (31.0%) | |
|---|---|---|---|---|---|---|---|---|---|---|---|---|
| Age | 85.7 | [4.5] | 85.8 | [4.4] | 85.9 | [4.5] | 86.3 | [4.6] | 86.8 | [4.7] | 87.5 | [4.9] |
| Female | 1,293 | [46.9%] | 1,022 | [50.2%] | 1,352 | [52.3%] | 1,051 | [53.4%] | 1,630 | [56.1%] | 3,241 | [58.9%] |
| Ethnicity | | | | | | | | | | | | |
| White | 2,499 | [90.7%] | 1,948 | [95.6%] | 2,353 | [91.0%] | 1,645 | [83.5%] | 2,316 | [79.7%] | 5,485 | [99.7%] |
| Mixed | 0 | [0.0%] | 27 | [1.3%] | 0 | [0.0%] | 0 | [0.0%] | 0 | [0.0%] | 0 | [0.0%] |
| Asian | 87 | [3.2%] | 27 | [1.3%] | 53 | [2.0%] | 295 | [15.0%] | 66 | [2.3%] | 12 | [0.2%] |
| Black | 125 | [4.5%] | 35 | [1.7%] | 47 | [1.8%] | 22 | [1.1%] | 204 | [7.0%] | 2 | [0.0%] |
| Other | 3 | [0.1%] | 0 | [0.0%] | 119 | [4.6%] | 1 | [0.1%] | 0 | [0.0%] | 0 | [0.0%] |
| unknown | 40 | [1.5%] | 0 | [0.0%] | 15 | [0.6%] | 7 | [0.4%] | 319 | [11.0%] | 0 | [0.0%] |
| IMD quintiles | | | | | | | | | | | | |
| 1 (least deprived) | 592 | [21.5%] | 514 | [25.2%] | 591 | [22.8%] | 409 | [20.8%] | 613 | [21.1%] | 1,302 | [23.7%] |
| 2 | 601 | [21.8%] | 450 | [22.1%] | 560 | [21.6%] | 409 | [20.8%] | 610 | [21.0%] | 1,261 | [22.9%] |
| 3 | 513 | [18.6%] | 417 | [20.5%] | 546 | [21.1%] | 408 | [20.7%] | 549 | [18.9%] | 1,096 | [19.9%] |
| 4 | 552 | [20.0%] | 353 | [17.3%] | 462 | [17.9%] | 343 | [17.4%] | 559 | [19.2%] | 991 | [18.0%] |
| 5 (most deprived) | 496 | [18.0%] | 303 | [14.9%] | 428 | [16.5%] | 373 | [18.9%] | 574 | [19.8%] | 849 | [15.4%] |
| unknown | 0 | [0.0%] | 0 | [0.0%] | 0 | [0.0%] | 28 | [1.4%] | 0 | [0.0%] | 0 | [0.0%] |
| Usage in 2018/19 | | | | | | | | | | | | |
| Total care costs (£) | 17,075 | [7,229] | 16,182 | [6,201] | 17,081 | [7,502] | 16,644 | [6,726] | 16,971 | [7,009] | 15,912 | [6,147] |
| Planned care (£) | 3,661 | [4,951] | 4,136 | [5,205] | 4,247 | [6,274] | 3,571 | [4,977] | 2,901 | [4,484] | 3,188 | [4,922] |
| Unplanned care (£) | 13,413 | [7,728] | 12,038 | [6,852] | 12,826 | [7,041] | 13,071 | [7,175] | 14,066 | [7,090] | 12,716 | [6,628] |
| Secondary care (£) | 16,350 | [7,216] | 15,506 | [6,178] | 16,393 | [7,494] | 15,982 | [6,698] | 16,326 | [6,996] | 15,287 | [6,127] |
| Primary care (£) | 725 | [706] | 675 | [575] | 689 | [573] | 663 | [609] | 645 | [589] | 625 | [593] |
| N(EM) | 3.1 | [2.1] | 2.8 | [1.9] | 3 | [2.0] | 3.1 | [2.1] | 3.2 | [2.0] | 2.8 | [1.8] |
| N(ED) | 4 | [4.0] | 3.6 | [4.5] | 3.9 | [3.4] | 3.9 | [5.3] | 4.1 | [3.8] | 3.5 | [3.1] |
| N(EL) | 1.3 | [5.7] | 2 | [7.1] | 2.6 | [9.1] | 1.9 | [10.8] | 1.1 | [9.8] | 0.8 | [4.2] |
| N(OP) | 9.2 | [14.1] | 10.4 | [13.1] | 10.3 | [14.1] | 8.7 | [13.9] | 6.5 | [10.5] | 6.4 | [11.4] |
| N(GP) | 20.9 | [24.5] | 18.8 | [14.8] | 20.1 | [15.8] | 19 | [17.3] | 17 | [14.6] | 16.7 | [16.6] |
| N(CALIBER) | 24.7 | [7.0] | 21.9 | [6.6] | 24.1 | [7.1] | 23.7 | [7.3] | 22.5 | [6.9] | 19.7 | [6.5] |
| N(LTC) | 20.1 | [6.1] | 18 | [5.8] | 19.2 | [6.3] | 19.2 | [6.6] | 17.8 | [6.1] | 15.6 | [5.7] |
| N(acute) | 4.6 | [2.6] | 3.9 | [2.5] | 4.9 | [2.6] | 4.6 | [2.6] | 4.7 | [2.5] | 4.1 | [2.5] |
| Disease system | | | | | | | | | | | | |
| Cancers | 845 | [30.7%] | 1,544 | [75.8%] | 1,358 | [52.5%] | 773 | [39.2%] | 765 | [26.3%] | 1,352 | [24.6%] |
| Circulatory | 2,711 | [98.4%] | 1,959 | [96.2%] | 2,508 | [96.9%] | 1,925 | [97.7%] | 2,847 | [98.0%] | 5,281 | [96.0%] |
| Digestive | 2,061 | [74.8%] | 1,382 | [67.8%] | 2,020 | [78.1%] | 1,473 | [74.8%] | 1,949 | [67.1%] | 3,270 | [59.5%] |
| Ear conditions | 1,073 | [39.0%] | 870 | [42.7%] | 1,139 | [44.0%] | 845 | [42.9%] | 1,162 | [40.0%] | 2,180 | [39.6%] |
| Endocrine | 2,065 | [75.0%] | 1,218 | [59.8%] | 1,640 | [63.4%] | 1,298 | [65.9%] | 1,852 | [63.8%] | 3,165 | [57.6%] |
| Eye conditions | 1,913 | [69.5%] | 1,222 | [60.0%] | 1,764 | [68.2%] | 1,358 | [68.9%] | 2,004 | [69.0%] | 3,425 | [62.3%] |
| Genitourinary | 2,475 | [89.9%] | 1,699 | [83.4%] | 2,286 | [88.4%] | 1,763 | [89.5%] | 2,554 | [87.9%] | 4,660 | [84.7%] |
| Respiratory | 1,737 | [63.1%] | 1,153 | [56.6%] | 1,577 | [61.0%] | 1,299 | [65.9%] | 1,800 | [62.0%] | 2,985 | [54.3%] |
| Haematological/Immunological | 1,732 | [62.9%] | 1,068 | [52.4%] | 1,598 | [61.8%] | 1,200 | [60.9%] | 1,677 | [57.7%] | 2,835 | [51.6%] |
| Infectious Disease | 2,203 | [80.0%] | 1,518 | [74.5%] | 2,053 | [79.4%] | 1,598 | [81.1%] | 2,430 | [83.6%] | 4,277 | [77.8%] |
| Mental Health | 1,812 | [65.8%] | 1,185 | [58.2%] | 1,565 | [60.5%] | 1,193 | [60.6%] | 2,091 | [72.0%] | 3,593 | [65.3%] |
| Musculoskeletal | 2,362 | [85.8%] | 1,724 | [84.6%] | 2,233 | [86.3%] | 1,675 | [85.0%] | 2,467 | [84.9%] | 4,683 | [85.2%] |
| Neurological | 890 | [32.3%] | 506 | [24.8%] | 631 | [24.4%] | 306 | [15.5%] | 881 | [30.3%] | 557 | [10.1%] |
| Skin conditions | 1,075 | [39.0%] | 604 | [29.7%] | 1,053 | [40.7%] | 969 | [49.2%] | 758 | [26.1%] | 1,336 | [24.3%] |
| In-year Mortality | 707 | [25.7%] | 716 | [35.1%] | 751 | [29.0%] | 550 | [27.9%] | 865 | [29.8%] | 1,520 | [27.6%] |

Relative Prevalence for Top 20 Diseases

| (1) | | (2) | | (3) | |
|---|---|---|---|---|---|
| Ventricular tachycardia | 4.5 | Secondary malignancy_bone | 6.9 | Carcinoma in situ_cervical | 6.8 |
| Sleep apnoea | 4.3 | Secondary malignancy_liver | 6.5 | Actinic keratosis | 5.9 |
| Diabetic neuropathy | 4.2 | Secondary malignancy_lung | 6.3 | Primary malignancy_skin | 1.6 |
| Other cardiomyopathy | 4.0 | Secondary malignancy_other | 6.1 | Glomerulonephritis | 1.3 |
| Peripheral neuropathy | 2.3 | Secondary malignancy_lymph nodes | 5.4 | Primary malignancy_other | 1.3 |
| Secondary pulmonary hypertension | 1.8 | Primary malignancy_lung | 4.9 | Infections of the digestive system | 1.3 |
| Rheumatic valve disorder | 1.8 | Primary malignancy_other | 4.3 | Primary malignancy_bladder | 1.2 |
| Diabetic eye disease | 1.7 | Primary malignancy_prostate | 2.7 | Bronchiectasis | 1.2 |
| Peripheral arterial disease | 1.5 | Primary malignancy_bowel | 2.2 | Gastritis and duodenitis | 1.2 |
| Diabetes | 1.4 | Primary malignancy_bladder | 1.7 | Abdominal hernia | 1.2 |
| Left bundle branch block | 1.4 | Gastro-oesophageal reflux disease | 1.1 | Enthesopathies & synovial disorders | 1.1 |
| Nonrheumatic mitral valve disorders | 1.4 | Anxiety disorders | 1.1 | Diverticular disease of intestine | 1.1 |
| Multiple valve disorder | 1.3 | Diverticular disease of intestine | 1.1 | Peripheral neuropathy | 1.1 |
| Gout | 1.3 | Primary malignancy_skin | 1.1 | Gastro-oesophageal reflux disease | 1.1 |
| Atrioventricular blocks | 1.3 | Hyperplasia of prostate | 1.1 | Rheumatoid arthritis | 1.1 |
| Heart failure | 1.3 | COPD | 1.0 | Pleural effusion | 1.1 |
| Iron deficiency anaemia | 1.3 | Pleural effusion | 1.0 | Collapsed vertebra | 1.1 |
| Obesity | 1.3 | Enthesopathies & synovial disorders | 1.0 | Coronary heart disease NOS | 1.1 |
| Myocardial infarction | 1.3 | Diaphragmatic hernia | 1.0 | Spondylosis | 1.1 |
| Infection of skin and subcutaneous tissues | 1.2 | Collapsed vertebra | 1.0 | Diaphragmatic hernia | 1.1 |
| (4) | | (5) | | (6) | |
| Primary malignancy_bladder | 3.3 | Scoliosis | 4.4 | Fracture of hip | 1.4 |
| Glomerulonephritis | 1.5 | Dementia | 1.2 | Dementia | 1.2 |
| Unstable angina | 1.3 | Urinary incontinence | 1.1 | Osteoporosis | 1.1 |
| Gastritis and duodenitis | 1.3 | Diabetic eye disease | 1.1 | Delirium, not induced by alcohol and other psychoactive substances | 1.1 |
| Asthma | 1.3 | Infections of other or unspecified organs | 1.1 | Rheumatoid arthritis | 1.0 |
| Spondylosis | 1.3 | Delirium, not induced by alcohol and other psychoactive substances | 1.1 | Infection of skin and subcutaneous tissues | 1.0 |
| Primary malignancy_bowel | 1.3 | Pleural effusion | 1.1 | Collapsed vertebra | 1.0 |
| Bronchiectasis | 1.2 | Lower respiratory tract infections | 1.1 | Urinary tract infections | 1.0 |
| Rheumatoid arthritis | 1.2 | Multiple valve disorder | 1.1 | Thyroid disease | 1.0 |
| Hyperplasia of prostate | 1.2 | Other or unspecified infectious organisms | 1.1 | Bronchiectasis | 1.0 |
| Diaphragmatic hernia | 1.2 | Osteoporosis | 1.1 | Lower respiratory tract infections | 1.0 |
| Iron deficiency anaemia | 1.2 | Collapsed vertebra | 1.0 | Bacterial diseases (excl TB) | 1.0 |
| Gastro-oesophageal reflux disease | 1.2 | Urinary tract infections | 1.0 | Chronic kidney disease | 1.0 |
| Urinary incontinence | 1.2 | Nonrheumatic mitral valve disorders | 1.0 | Osteoarthritis (excl spine) | 1.0 |
| Diverticular disease of intestine | 1.2 | Bacterial diseases (excl TB) | 1.0 | Atrial fibrillation | 1.0 |
| Abdominal hernia | 1.2 | Right bundle branch block combinations | 1.0 | Urinary incontinence | 1.0 |
| Stable angina | 1.1 | Depression | 1.0 | Other or unspecified infectious organisms | 1.0 |
| Infections of the digestive system | 1.1 | Atrial fibrillation | 1.0 | Left bundle branch block | 1.0 |
| Nonrheumatic aortic valve disorders | 1.1 | Diabetes | 1.0 | Hypertension | 1.0 |
| Myocardial infarction | 1.1 | Left bundle branch block | 1.0 | Respiratory failure | 1.0 |

**Figure S4 – Disease-System Profiles of in-year decedent high-cost clusters (optimal hierarchical 4-cluster solution)**

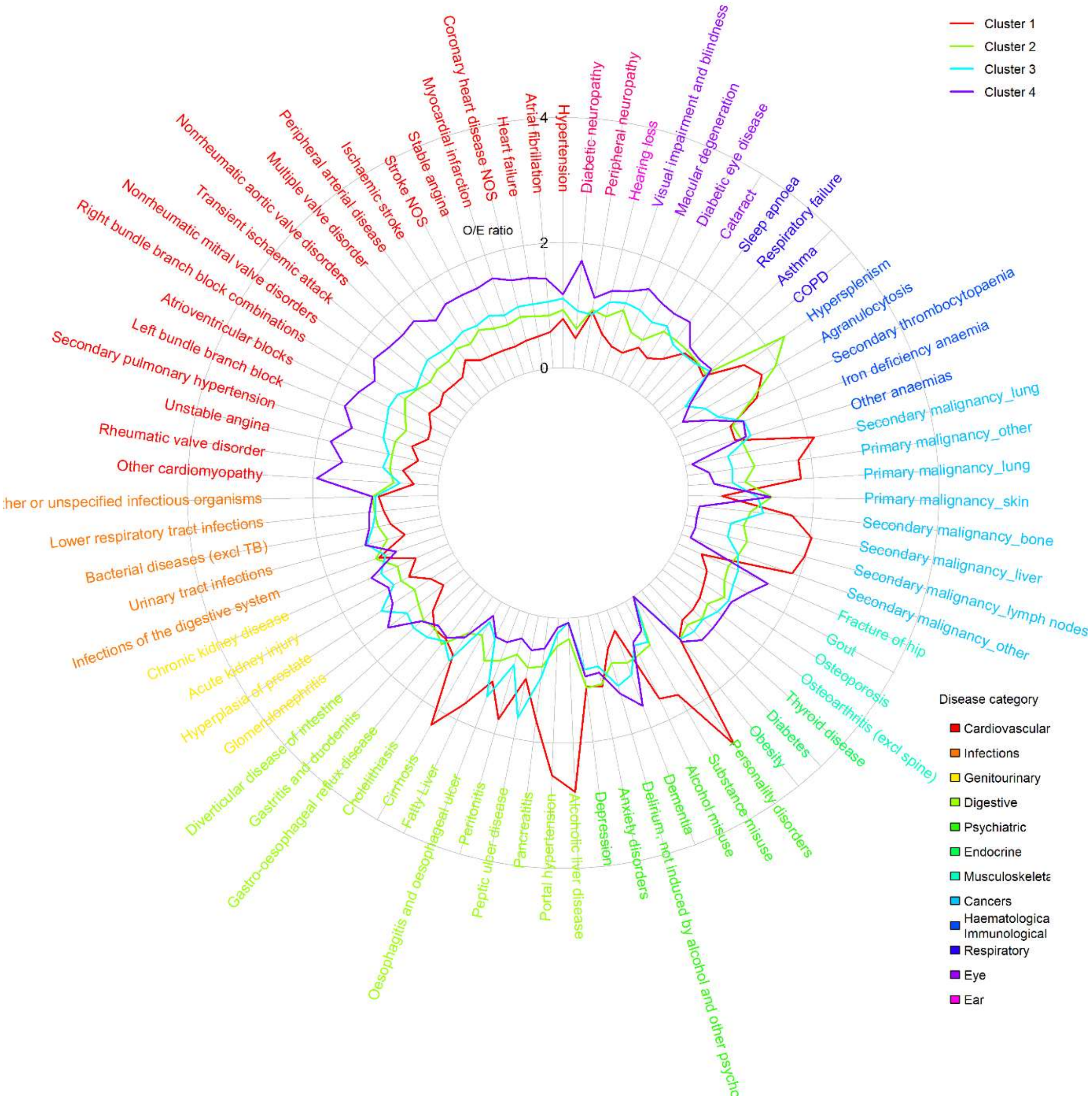

*Note: Relative prevalence is estimated according to relative prevalence to other patients in high-cost patient group*

**Table S6 – Descriptive statistics of in-year decedent high-cost clusters, based on 50% random sample (N=10,759; optimal hierarchical 4-cluster solution)**

| | (1) Personality disorders, liver disease, and metastatic lung cancer N=3,616 (33.6%) | | (2) Immunological and haematological disorders N=1,166 (10.8%) | | (3) Upper GI and prostate multimorbidity N=2,383 (22.1%) | | (4) Cardiovascular disease and dementia N=3,594 (33.4%) | |
|---|---|---|---|---|---|---|---|---|
| Age | 67.5 | [14.6] | 73.7 | [13.3] | 79.7 | [10.8] | 83.1 | [9.5] |
| Female | 1,892 | [52.3%] | 551 | [47.3%] | 946 | [39.7%] | 1,654 | [46.0%] |
| Ethnicity | | | | | | | | |
| White | 3,037 | [84.0%] | 987 | [84.6%] | 2,032 | [85.3%] | 3,418 | [95.1%] |
| Mixed | 0 | [0.0%] | 0 | [0.0%] | 36 | [1.5%] | 0 | [0.0%] |
| Asian | 188 | [5.2%] | 43 | [3.7%] | 79 | [3.3%] | 133 | [3.7%] |
| Black | 102 | [2.8%] | 13 | [1.1%] | 204 | [8.6%] | 20 | [0.6%] |
| Other | 5 | [0.1%] | 102 | [8.7%] | 0 | [0.0%] | 0 | [0.0%] |
| unknown | 284 | [7.9%] | 21 | [1.8%] | 32 | [1.3%] | 23 | [0.6%] |
| IMD quintiles | | | | | | | | |
| 1 (least deprived) | 678 | [18.8%] | 212 | [18.2%] | 460 | [19.3%] | 715 | [19.9%] |
| 2 | 678 | [18.8%] | 264 | [22.6%] | 464 | [19.5%] | 762 | [21.2%] |
| 3 | 651 | [18.0%] | 236 | [20.2%] | 454 | [19.1%] | 712 | [19.8%] |
| 4 | 740 | [20.5%] | 211 | [18.1%] | 513 | [21.5%] | 682 | [19.0%] |
| 5 (most deprived) | 869 | [24.0%] | 243 | [20.8%] | 492 | [20.6%] | 707 | [19.7%] |
| unknown | 0 | [0.0%] | 0 | [0.0%] | 0 | [0.0%] | 16 | [0.4%] |
| Usage in 2018/19 | | | | | | | | |
| Total care costs (£) | 17,969 | [9,297] | 18,248 | [9,745] | 17,611 | [7,762] | 17,296 | [7,744] |
| Planned care (£) | 4,918 | [7,583] | 4,354 | [7,723] | 3,210 | [5,193] | 2,737 | [5,126] |
| Unplanned care (£) | 13,052 | [7,607] | 13,909 | [7,385] | 14,388 | [7,053] | 14,564 | [7,156] |
| Secondary care (£) | 17,452 | [9,301] | 17,707 | [9,765] | 17,031 | [7,751] | 16,696 | [7,706] |
| Primary care (£) | 517 | [473] | 541 | [482] | 579 | [548] | 600 | [583] |
| N(EM) | 3.1 | [2.2] | 3 | [1.9] | 3.1 | [1.9] | 3 | [1.8] |
| N(ED) | 3.3 | [4.2] | 3.3 | [3.4] | 3.3 | [3.0] | 3.3 | [2.9] |
| N(EL) | 3.7 | [9.0] | 3.3 | [9.5] | 2.3 | [10.4] | 1.3 | [7.4] |
| N(OP) | 11.4 | [13.5] | 9.9 | [12.2] | 7.9 | [11.4] | 6.2 | [11.9] |
| N(GP) | 14.4 | [12.3] | 14.8 | [12.5] | 15.6 | [13.4] | 15.5 | [14.5] |
| N(CALIBER) | 19.2 | [7.1] | 22.2 | [7.4] | 23.1 | [7.1] | 23.7 | [7.3] |
| N(LTC) | 15 | [6.2] | 17.6 | [6.6] | 18 | [6.2] | 18.8 | [6.6] |
| N(acute) | 4.2 | [2.6] | 4.7 | [2.4] | 5.1 | [2.6] | 4.9 | [2.3] |
| Disease system | | | | | | | | |
| Cancers | 2,190 | [60.6%] | 697 | [59.8%] | 1,200 | [50.4%] | 1,159 | [32.2%] |
| Circulatory | 2,954 | [81.7%] | 1,012 | [86.8%] | 2,273 | [95.4%] | 3,518 | [97.9%] |
| Digestive | 2,416 | [66.8%] | 794 | [68.1%] | 1,732 | [72.7%] | 2,275 | [63.3%] |
| Ear conditions | 686 | [19.0%] | 311 | [26.7%] | 764 | [32.1%] | 1,310 | [36.4%] |
| Endocrine | 2,014 | [55.7%] | 705 | [60.5%] | 1,543 | [64.8%] | 2,435 | [67.8%] |
| Eye conditions | 1,058 | [29.3%] | 579 | [49.7%] | 1,302 | [54.6%] | 2,286 | [63.6%] |
| Genitourinary | 2,247 | [62.1%] | 866 | [74.3%] | 2,091 | [87.7%] | 3,226 | [89.8%] |
| Respiratory | 2,430 | [67.2%] | 840 | [72.0%] | 1,644 | [69.0%] | 2,580 | [71.8%] |
| Haematological/Immunological | 1,978 | [54.7%] | 719 | [61.7%] | 1,498 | [62.9%] | 2,091 | [58.2%] |
| Infectious Disease | 2,893 | [80.0%] | 1,017 | [87.2%] | 2,055 | [86.2%] | 3,196 | [88.9%] |
| Mental Health | 2,025 | [56.0%] | 751 | [64.4%] | 1,442 | [60.5%] | 2,449 | [68.1%] |
| Musculoskeletal | 2,249 | [62.2%] | 839 | [72.0%] | 1,886 | [79.1%] | 2,919 | [81.2%] |
| Neurological | 730 | [20.2%] | 361 | [31.0%] | 441 | [18.5%] | 937 | [26.1%] |
| Skin conditions | 1,035 | [28.6%] | 419 | [35.9%] | 656 | [27.5%] | 1,103 | [30.7%] |

Relative Prevalence for Top 20 Diseases

| (1) | | (2) | |
|---|---|---|---|
| Personality disorders | 2.9 | Hypersplenism | 2.3 |
| Alcoholic liver disease | 2.8 | Agranulocytosis | 1.9 |
| Portal hypertension | 2.5 | Secondary thrombocytopaenia | 1.3 |
| Cirrhosis | 2.3 | Primary malignancy_skin | 1.3 |
| Secondary malignancy_lung | 2.1 | Infections of the digestive system | 1.2 |
| Secondary malignancy_liver | 2.0 | Depression | 1.1 |
| Secondary malignancy_lymph nodes | 2.0 | Anxiety disorders | 1.1 |
| Secondary malignancy_other | 1.9 | Gastro-oesophageal reflux disease | 1.1 |
| Primary malignancy_lung | 1.8 | Primary malignancy_other | 1.1 |
| Primary malignancy_other | 1.8 | Visual impairment and blindness | 1.1 |
| Peritonitis | 1.8 | Osteoporosis | 1.1 |
| Fatty Liver | 1.7 | Other or unspecified infectious organisms | 1.0 |
| Substance misuse | 1.7 | Cataract | 1.0 |
| Agranulocytosis | 1.7 | Secondary malignancy_lymph nodes | 1.0 |
| Secondary malignancy_bone | 1.7 | Lower respiratory tract infections | 1.0 |
| Pancreatitis | 1.6 | Sleep apnoea | 1.0 |
| Alcohol misuse | 1.6 | Respiratory failure | 1.0 |
| Hypersplenism | 1.5 | Diverticular disease of intestine | 1.0 |
| Secondary thrombocytopaenia | 1.4 | Secondary malignancy_lung | 1.0 |
| Oesophagitis and oesophageal ulcer | 1.2 | Secondary malignancy_bone | 1.0 |
| (3) | | (4) | |
| Peptic ulcer disease | 1.7 | Other cardiomyopathy | 1.9 |
| Oesophagitis and oesophageal ulcer | 1.5 | Unstable angina | 1.8 |
| Hyperplasia of prostate | 1.5 | Left bundle branch block | 1.8 |
| Urinary tract infections | 1.2 | Diabetic neuropathy | 1.7 |
| Cholelithiasis | 1.2 | Nonrheumatic mitral valve disorders | 1.7 |
| Diverticular disease of intestine | 1.2 | Atrioventricular blocks | 1.6 |
| Secondary malignancy_bone | 1.2 | Multiple valve disorder | 1.6 |
| Delirium, not induced by alcohol and other psychoactive substances | 1.2 | Dementia | 1.6 |
| Visual impairment and blindness | 1.2 | Transient ischaemic attack | 1.6 |
| Chronic kidney disease | 1.2 | Myocardial infarction | 1.6 |
| Macular degeneration | 1.2 | Nonrheumatic aortic valve disorders | 1.6 |
| Gastritis and duodenitis | 1.2 | Fracture of hip | 1.6 |
| Osteoporosis | 1.1 | Rheumatic valve disorder | 1.6 |
| Glomerulonephritis | 1.1 | Ischaemic stroke | 1.6 |
| Hearing loss | 1.1 | Stroke NOS | 1.5 |
| Cataract | 1.1 | Stable angina | 1.5 |
| Other anaemias | 1.1 | Macular degeneration | 1.5 |
| Osteoarthritis (excl spine) | 1.1 | Glomerulonephritis | 1.5 |
| Hypertension | 1.1 | Secondary pulmonary hypertension | 1.5 |
| Iron deficiency anaemia | 1.1 | Right bundle branch block combinations | 1.5 |

**Figure S5 – Disease-System Profiles of Unplanned-Care High-Cost Clusters (optimal K-medoids 6-Cluster Solution)**

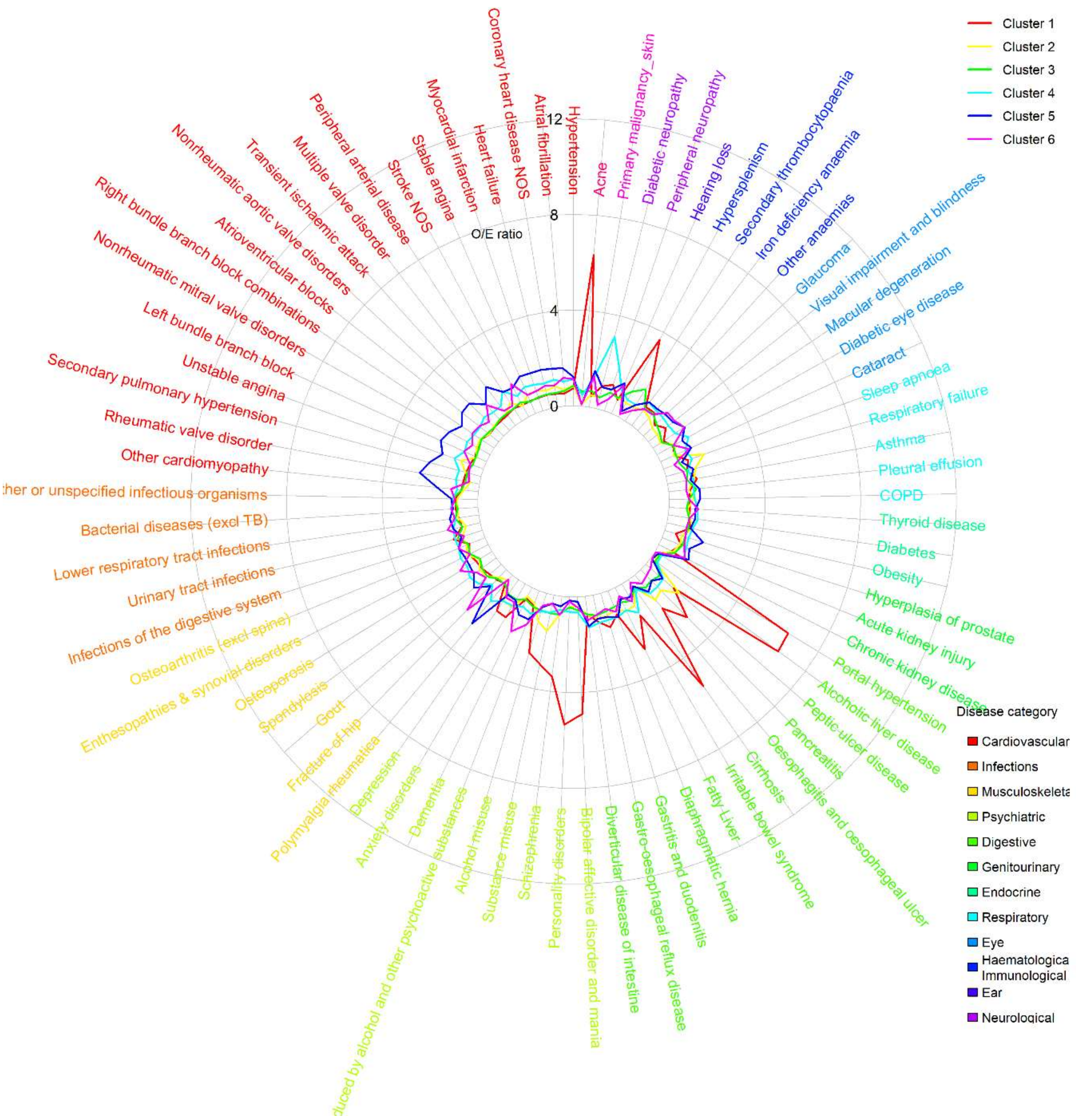

*Note: Relative prevalence is estimated according to relative prevalence to other patients in high-cost patient group*

### Table S7 – Descriptive statistics of unplanned care high-cost clusters, based on 50% random sample (N=52,206; optimal k-medoids 6-cluster solution)

| | (1) | | (2) | | (3) | | (4) | | (5) | | (6) | |
|---|---|---|---|---|---|---|---|---|---|---|---|---|
| | Liver disease & mental health | | Sleep apnoea, gastro-reflux and diabetes | | Immunological and haematological disorders | | Diabetic and vascular complications | | Polymyalgia and cardiovascular disease | | Hip fracture, cognitive, and sensory impairment | |
| | N=6,703 (12.8%) | | N=8,461 (16.2%) | | N=7,860 (15.1%) | | N=8,533 (16.3%) | | N=10,845 (20.8%) | | N=9,804 (18.8%) | |
| Age | 57.9 | [19.5] | 64.8 | [17.7] | 67.8 | [17.3] | 73.3 | [15.1] | 81.3 | [10.0] | 82.2 | [10.8] |
| Female | 3,639 | [54.3%] | 3,748 | [44.3%] | 3,591 | [45.7%] | 4,510 | [52.9%] | 4,267 | [39.3%] | 6,258 | [63.8%] |
| Ethnicity | | | | | | | | | | | | |
| White | 5,817 | [86.8%] | 7,590 | [89.7%] | 4,779 | [60.8%] | 7,804 | [91.5%] | 10,256 | [94.6%] | 9,371 | [95.6%] |
| Mixed | 0 | [0.0%] | 0 | [0.0%] | 236 | [3.0%] | 0 | [0.0%] | 1 | [0.0%] | 0 | [0.0%] |
| Asian | 367 | [5.5%] | 777 | [9.2%] | 187 | [2.4%] | 374 | [4.4%] | 405 | [3.7%] | 255 | [2.6%] |
| Black | 290 | [4.3%] | 72 | [0.9%] | 908 | [11.6%] | 215 | [2.5%] | 144 | [1.3%] | 157 | [1.6%] |
| Other | 8 | [0.1%] | 0 | [0.0%] | 495 | [6.3%] | 8 | [0.1%] | 0 | [0.0%] | 0 | [0.0%] |
| unknown | 221 | [3.3%] | 22 | [0.3%] | 1,255 | [16.0%] | 132 | [1.5%] | 39 | [0.4%] | 21 | [0.2%] |
| IMD quintiles | | | | | | | | | | | | |
| 1 (least deprived) | 935 | [13.9%] | 1,346 | [15.9%] | 1,373 | [17.5%] | 1,599 | [18.7%] | 2,264 | [20.9%] | 1,983 | [20.2%] |
| 2 | 1,100 | [16.4%] | 1,507 | [17.8%] | 1,338 | [17.0%] | 1,670 | [19.6%] | 2,207 | [20.4%] | 2,136 | [21.8%] |
| 3 | 1,247 | [18.6%] | 1,552 | [18.3%] | 1,444 | [18.4%] | 1,621 | [19.0%] | 2,089 | [19.3%] | 2,014 | [20.5%] |
| 4 | 1,544 | [23.0%] | 1,818 | [21.5%] | 1,837 | [23.4%] | 1,750 | [20.5%] | 2,014 | [18.6%] | 1,814 | [18.5%] |
| 5 (most deprived) | 1,877 | [28.0%] | 2,238 | [26.5%] | 1,868 | [23.8%] | 1,893 | [22.2%] | 2,199 | [20.3%] | 1,857 | [18.9%] |
| unknown | 0 | [0.0%] | 0 | [0.0%] | 0 | [0.0%] | 0 | [0.0%] | 72 | [0.7%] | 0 | [0.0%] |
| Usage in 2018/19 | | | | | | | | | | | | |
| Total care costs (£) | 16,298 | [9,987] | 14,666 | [8,220] | 16,590 | [10,607] | 16,394 | [9,421] | 15,469 | [8,075] | 14,154 | [7,154] |
| Planned care (£) | 2,600 | [5,278] | 2,317 | [4,497] | 3,662 | [7,312] | 2,991 | [5,464] | 2,194 | [3,889] | 1,603 | [3,014] |
| Unplanned care (£) | 13,708 | [8,112] | 12,350 | [6,544] | 12,951 | [7,032] | 13,412 | [7,275] | 13,276 | [6,830] | 12,549 | [6,233] |
| Secondary care (£) | 15,781 | [9,944] | 14,215 | [8,178] | 16,120 | [10,590] | 15,796 | [9,377] | 14,872 | [8,009] | 13,603 | [7,095] |
| Primary care (£) | 517 | [506] | 451 | [439] | 471 | [456] | 597 | [536] | 597 | [540] | 551 | [529] |
| N(EM) | 4.1 | [4.1] | 3 | [2.8] | 3.2 | [2.5] | 3.3 | [2.4] | 3 | [2.0] | 2.7 | [1.8] |
| N(ED) | 8.1 | [14.5] | 4.5 | [9.8] | 3.9 | [5.6] | 4.4 | [5.7] | 3.9 | [4.6] | 3.6 | [3.7] |
| N(EL) | 2 | [10.4] | 1.4 | [7.3] | 3.3 | [9.7] | 2.6 | [12.9] | 1.3 | [9.2] | 0.6 | [3.8] |
| N(OP) | 10 | [15.8] | 8.9 | [13.2] | 12.5 | [15.8] | 11.1 | [15.7] | 8.2 | [11.9] | 6.2 | [12.1] |
| N(GP) | 16.9 | [15.8] | 15.1 | [13.8] | 14.3 | [13.0] | 18.9 | [16.3] | 17.6 | [15.0] | 14.9 | [13.4] |
| N(CALIBER) | 19.5 | [8.4] | 16.1 | [7.9] | 17.7 | [7.2] | 22.6 | [7.9] | 23.2 | [7.2] | 18.9 | [6.6] |
| N(LTC) | 15.5 | [7.3] | 12.7 | [6.7] | 13.9 | [6.3] | 18.1 | [6.9] | 18.8 | [6.3] | 14.8 | [5.8] |
| N(acute) | 4 | [2.7] | 3.5 | [2.6] | 3.8 | [2.5] | 4.5 | [2.6] | 4.3 | [2.5] | 4.1 | [2.4] |
| Disease system | | | | | | | | | | | | |
| Cancers | 1,098 | [16.4%] | 1,080 | [12.8%] | 4,905 | [62.4%] | 2,614 | [30.6%] | 3,558 | [32.8%] | 2,521 | [25.7%] |
| Circulatory | 4,765 | [71.1%] | 6,629 | [78.3%] | 6,218 | [79.1%] | 7,828 | [91.7%] | 10,633 | [98.0%] | 9,209 | [93.9%] |
| Digestive | 5,092 | [76.0%] | 5,367 | [63.4%] | 4,551 | [57.9%] | 6,271 | [73.5%] | 7,473 | [68.9%] | 5,593 | [57.0%] |
| Ear conditions | 1,195 | [17.8%] | 1,749 | [20.7%] | 1,574 | [20.0%] | 2,489 | [29.2%] | 3,943 | [36.4%] | 3,261 | [33.3%] |
| Endocrine | 4,033 | [60.2%] | 5,136 | [60.7%] | 4,432 | [56.4%] | 6,059 | [71.0%] | 7,122 | [65.7%] | 5,771 | [58.9%] |
| Eye conditions | 2,293 | [34.2%] | 2,668 | [31.5%] | 2,640 | [33.6%] | 4,554 | [53.4%] | 6,690 | [61.7%] | 5,439 | [55.5%] |
| Genitourinary | 4,321 | [64.5%] | 4,556 | [53.8%] | 4,922 | [62.6%] | 6,892 | [80.8%] | 9,167 | [84.5%] | 7,694 | [78.5%] |
| Respiratory | 4,019 | [60.0%] | 4,863 | [57.5%] | 4,667 | [59.4%] | 5,198 | [60.9%] | 7,281 | [67.1%] | 4,941 | [50.4%] |
| Haematological/Immunological | 3,521 | [52.5%] | 3,236 | [38.2%] | 4,075 | [51.8%] | 4,747 | [55.6%] | 6,114 | [56.4%] | 4,722 | [48.2%] |
| Infectious Disease | 4,878 | [72.8%] | 5,687 | [67.2%] | 5,956 | [75.8%] | 6,812 | [79.8%] | 8,765 | [80.8%] | 7,604 | [77.6%] |
| Mental Health | 5,381 | [80.3%] | 4,945 | [58.4%] | 4,057 | [51.6%] | 5,522 | [64.7%] | 6,669 | [61.5%] | 7,123 | [72.7%] |
| Musculoskeletal | 4,040 | [60.3%] | 5,332 | [63.0%] | 4,780 | [60.8%] | 7,030 | [82.4%] | 8,694 | [80.2%] | 7,941 | [81.0%] |
| Neurological | 2,360 | [35.2%] | 1,689 | [20.0%] | 1,528 | [19.4%] | 3,228 | [37.8%] | 2,551 | [23.5%] | 2,010 | [20.5%] |
| Skin conditions | 2,533 | [37.8%] | 2,250 | [26.6%] | 2,008 | [25.5%] | 3,226 | [37.8%] | 3,597 | [33.2%] | 2,670 | [27.2%] |
| In-year Mortality | 1,218 | [18.2%] | 1,366 | [16.1%] | 3,275 | [41.7%] | 1,880 | [22.0%] | 3,487 | [32.2%] | 2,433 | [24.8%] |

Relative Prevalence for Top 20 Diseases

| (1) | | (2) | | (3) | |
|---|---|---|---|---|---|
| Alcoholic liver disease | 6.6 | Peptic ulcer disease | 1.9 | Secondary thrombocytopaenia | 1.6 |
| Portal hypertension | 6.5 | Sleep apnoea | 1.8 | Hypersplenism | 1.2 |
| Acne | 6.3 | Substance misuse | 1.5 | Pleural effusion | 1.2 |
| Cirrhosis | 5.4 | Oesophagitis and oesophageal ulcer | 1.3 | Infections of the digestive system | 1.1 |
| Personality disorders | 5.3 | Respiratory failure | 1.3 | Other or unspecified infectious organisms | 1.0 |
| Bipolar affective disorder and mania | 4.9 | Alcohol misuse | 1.2 | Lower respiratory tract infections | 1.0 |
| Hypersplenism | 3.7 | Pancreatitis | 1.2 | Other anaemias | 1.0 |
| Schizophrenia | 3.4 | Asthma | 1.1 | Obesity | 0.9 |
| Substance misuse | 2.9 | Anxiety disorders | 1.1 | Respiratory failure | 0.9 |
| Fatty Liver | 2.9 | Obesity | 1.1 | Acute kidney injury | 0.9 |
| Pancreatitis | 2.8 | Depression | 1.1 | Hyperplasia of prostate | 0.9 |
| Alcohol misuse | 2.6 | Fatty Liver | 1.1 | Iron deficiency anaemia | 0.9 |
| Secondary thrombocytopaenia | 2.1 | Irritable bowel syndrome | 1.1 | Diabetes | 0.9 |
| Oesophagitis and oesophageal ulcer | 1.8 | COPD | 1.0 | Enthesopathies & synovial disorders | 0.9 |
| Anxiety disorders | 1.6 | Unstable angina | 1.0 | Gastro-oesophageal reflux disease | 0.9 |
| Depression | 1.6 | Gastro-oesophageal reflux disease | 1.0 | Bacterial diseases (excl TB) | 0.8 |
| Peptic ulcer disease | 1.5 | Gastritis and duodenitis | 1.0 | Hypertension | 0.8 |
| Irritable bowel syndrome | 1.5 | Enthesopathies & synovial disorders | 1.0 | Asthma | 0.8 |
| Gastritis and duodenitis | 1.5 | Diabetes | 0.9 | COPD | 0.8 |
| Infections of the digestive system | 1.3 | Peripheral arterial disease | 0.9 | Irritable bowel syndrome | 0.8 |
| (4) | | (5) | | (6) | |
| Diabetic neuropathy | 3.1 | Polymyalgia rheumatica | 2.6 | Fracture of hip | 2.4 |
| Peripheral neuropathy | 1.9 | Rheumatic valve disorder | 2.5 | Dementia | 2.0 |
| Diabetic eye disease | 1.5 | Secondary pulmonary hypertension | 2.2 | Macular degeneration | 1.6 |
| Peripheral arterial disease | 1.5 | Left bundle branch block | 2.1 | Osteoporosis | 1.6 |
| Polymyalgia rheumatica | 1.4 | Multiple valve disorder | 2.0 | Stroke NOS | 1.5 |
| Irritable bowel syndrome | 1.4 | Nonrheumatic aortic valve disorders | 2.0 | Delirium, not induced by alcohol and other psychoactive substances | 1.5 |
| Spondylosis | 1.4 | Atrioventricular blocks | 2.0 | Transient ischaemic attack | 1.4 |
| Diverticular disease of intestine | 1.3 | Nonrheumatic mitral valve disorders | 2.0 | Glaucoma | 1.4 |
| Unstable angina | 1.3 | Unstable angina | 1.8 | Visual impairment and blindness | 1.4 |
| Enthesopathies & synovial disorders | 1.3 | Other cardiomyopathy | 1.7 | Urinary tract infections | 1.4 |
| Sleep apnoea | 1.3 | Stable angina | 1.7 | Primary malignancy_skin | 1.3 |
| Diabetes | 1.2 | Hyperplasia of prostate | 1.7 | Hearing loss | 1.3 |
| Gout | 1.2 | Heart failure | 1.7 | Cataract | 1.3 |
| Primary malignancy_skin | 1.2 | Myocardial infarction | 1.7 | Thyroid disease | 1.2 |
| Iron deficiency anaemia | 1.2 | Right bundle branch block combinations | 1.6 | Chronic kidney disease | 1.2 |
| Thyroid disease | 1.2 | Coronary heart disease NOS | 1.6 | Osteoarthritis (excl spine) | 1.2 |
| Osteoporosis | 1.2 | Atrial fibrillation | 1.6 | Atrial fibrillation | 1.2 |
| Infections of the digestive system | 1.2 | Macular degeneration | 1.6 | Other cardiomyopathy | 1.1 |
| Stable angina | 1.2 | Primary malignancy_skin | 1.5 | Hypertension | 1.1 |
| Gastro-oesophageal reflux disease | 1.2 | Gout | 1.5 | Bacterial diseases (excl TB) | 1.1 |

#### Table S8 – Cross-Method Comparison of Optimal Clustering

| Hierarchical, 9 clusters (optimal) | K-medoids, 6 clusters | Fuzzy K-medoids, 7 clusters |
|---|---|---|
| **Liver disease & mental health** | ***Matched*** (alcoholic liver disease, portal hypertension, acne, personality disorders, cirrhosis, bipolar affective disorder and mania, substance misuse, schizophrenia, chronic fatigue syndrome, hypersplenism, pancreatitis, fatty liver, alcohol misuse, epilepsy, cholelithiasis) | ***Matched*** (alcoholic liver disease, portal hypertension, personality disorders, cirrhosis, acne, bipolar affective disorder and mania, substance misuse, schizophrenia, hypersplenism, pancreatitis, chronic fatigue syndrome, fatty liver, alcohol misuse, epilepsy, cholelithiasis) |
| **Sleep apnoea, gastro-reflux and diabetes** | ***Dropped, embedded within a novel cluster "Cardiometabolic disease"*** *(rheumatic valve disorder, diabetic neuropathy/eye disease, secondary pulmonary hypertension, cardiomyopathy, sleep apnoea, and peptic ulcer disease).* | ***Dropped, embedded within novel cluster "Cardiometabolic disease"*** *(rheumatic valve disorder, cardiomyopathy, diabetic neuropathy/eye disease, secondary pulmonary hypertension, nonrheumatic mitral valve, multiple valve disorder, heart failure, and sleep apnoea).* |
| **Chronic fatigue, immunological and haematological disorders** | *Dropped, embedded within "Nodal metastases" cluster (hypersplenism, secondary thrombocytopaenia, chronic fatigue syndrome embedded).* | ***Matched*** *(hypersplenism, chronic fatigue syndrome, acne, secondary thrombocytopaenia, and irritable bowel syndrome)* |
| **Low multimorbidity** | ***Matched*** *(highest O/E prevalence of all diseases was 1.1).* | ***Matched*** *(highest O/E prevalence was 1.2 for peptic ulcer disease, all others below 1.2).* |
| **Nodal metastases** | ***Matched*** *(nodal metastases, hypersplenism, skin cancer, and secondary thrombocytopaenia).* | ***Matched*** *(nodal metastases, skin cancer).* |
| **Complex multimorbidity** | *Dropped* | *Dropped* |
| **Prostate cancer (with nodal metastasis)** | ***Matched*** *(prostate primary malignancy, secondary malignancy in lymph node, and hyperplasia of prostate)* | ***Matched*** *(prostate primary malignancy and hyperplasia of prostate)* |
| **Prostate, stroke & sensory impairment** | *Dropped, split between "Prostate cancer (with nodal metastasis)" and "Dementia, cardiovascular disease, and sensory impairment"* | *Dropped, split between "Prostate cancer (with nodal metastasis)" and "Dementia, cardiovascular disease, and sensory impairment"* |
| **Cardiovascular disease and dementia** | ***Reproduced with more dementia and sensory impairment diagnoses "Dementia, cardiovascular disease, and sensory impairment"*** *(dementia, left bundle branch blocks, macular degeneration, ischaemic stroke, atrioventricular blocks, delirium, stroke, nonrheumatic aortic valve disorder, multiple valve disorder, and visual impairment & blindness)* | ***Reproduced with more dementia and sensory impairment diagnoses "Dementia, cardiovascular disease, and sensory impairment"*** *(dementia, macular degeneration, delirium, visual impairment & blindness, left bundle branch block, atrioventricular blocks, glaucoma, urinary tract infections, osteoporosis, transient ischaemic attack, ischaemic stroke, stroke, cataract, valve disorders, hearing loss)* |

### Table S9 – Descriptive statistics of high-cost clusters from k-medoids and fuzzy k-medoids, based on 50% random sample (N=52,175)

a. K-medoids, k=6

| | (1) Liver disease & mental health | | (2) Low multimorbidity | | (3) Nodal metastases | | (4) Cardiometabolic disease | | (5) Prostate cancer (with nodal metastasis) | | (6) Dementia, cardiovascular disease, and sensory impairment | |
|---|---|---|---|---|---|---|---|---|---|---|---|---|
| | N=8,014 (15.4%) | | N=7,916 (15.2%) | | N=8,304 (15.9%) | | N=7,196 (13.8%) | | N=9,983 (19.1%) | | N=10,762 (20.6%) | |
| Age | 55.9 | [18.3] | 65.6 | [15.7] | 67.2 | [17.3] | 70.2 | [15.5] | 75.3 | [11.9] | 81 | [11.3] |
| Female | 3,960 | [49.4%] | 3,847 | [48.6%] | 5,329 | [64.2%] | 3,270 | [45.4%] | 3,404 | [34.1%] | 5,536 | [51.4%] |
| Ethnicity | | | | | | | | | | | | |
| White | 6,735 | [84.0%] | 7,656 | [96.7%] | 6,608 | [79.6%] | 5,047 | [70.1%] | 9,414 | [94.3%] | 9,920 | [92.2%] |
| Mixed | 0 | [0.0%] | 0 | [0.0%] | 1 | [0.0%] | 240 | [3.3%] | 1 | [0.0%] | 0 | [0.0%] |
| Asian | 391 | [4.9%] | 172 | [2.2%] | 330 | [4.0%] | 1,149 | [16.0%] | 237 | [2.4%] | 185 | [1.7%] |
| Black | 460 | [5.7%] | 68 | [0.9%] | 319 | [3.8%] | 657 | [9.1%] | 257 | [2.6%] | 110 | [1.0%] |
| Other | 3 | [0.0%] | 0 | [0.0%] | 12 | [0.1%] | 0 | [0.0%] | 0 | [0.0%] | 520 | [4.8%] |
| unknown | 425 | [5.3%] | 20 | [0.3%] | 1,034 | [12.5%] | 103 | [1.4%] | 74 | [0.7%] | 27 | [0.3%] |
| IMD quintiles | | | | | | | | | | | | |
| 1 (least deprived) | 1,159 | [14.5%] | 1,709 | [21.6%] | 1,709 | [20.6%] | 1,036 | [14.4%] | 2,121 | [21.2%] | 2,154 | [20.0%] |
| 2 | 1,251 | [15.6%] | 1,686 | [21.3%] | 1,693 | [20.4%] | 1,240 | [17.2%] | 2,059 | [20.6%] | 2,351 | [21.8%] |
| 3 | 1,469 | [18.3%] | 1,706 | [21.6%] | 1,659 | [20.0%] | 1,397 | [19.4%] | 1,913 | [19.2%] | 2,057 | [19.1%] |
| 4 | 1,849 | [23.1%] | 1,438 | [18.2%] | 1,650 | [19.9%] | 1,708 | [23.7%] | 1,907 | [19.1%] | 2,080 | [19.3%] |
| 5 (most deprived) | 2,286 | [28.5%] | 1,377 | [17.4%] | 1,593 | [19.2%] | 1,815 | [25.2%] | 1,904 | [19.1%] | 2,120 | [19.7%] |
| unknown | 0 | [0.0%] | 0 | [0.0%] | 0 | [0.0%] | 0 | [0.0%] | 79 | [0.8%] | 0 | [0.0%] |
| Usage in 2018/19 | | | | | | | | | | | | |
| Total care costs (£) | 18,209 | [10,185] | 16,798 | [9,261] | 17,716 | [9,406] | 18,568 | [9,780] | 17,061 | [7,712] | 16,952 | [7,582] |
| Planned care (£) | 6,018 | [8,246] | 8,632 | [9,251] | 7,601 | [8,785] | 5,794 | [7,998] | 6,006 | [6,874] | 3,912 | [6,004] |
| Unplanned care (£) | 12,183 | [9,514] | 8,174 | [7,930] | 10,124 | [8,218] | 12,786 | [9,187] | 11,051 | [7,944] | 13,036 | [7,638] |
| Secondary care (£) | 17,701 | [10,184] | 16,352 | [9,285] | 17,189 | [9,415] | 17,990 | [9,767] | 16,495 | [7,709] | 16,358 | [7,555] |
| Primary care (£) | 508 | [532] | 446 | [443] | 527 | [491] | 578 | [555] | 566 | [509] | 595 | [579] |
| N(EM) | 3.6 | [3.7] | 2 | [2.1] | 2.7 | [2.6] | 3.1 | [2.9] | 2.7 | [2.1] | 2.9 | [2.1] |
| N(ED) | 6.9 | [14.6] | 2.7 | [4.9] | 3.7 | [6.6] | 4.1 | [6.5] | 3.3 | [4.0] | 3.8 | [4.2] |
| N(EL) | 3.6 | [11.6] | 4.1 | [9.8] | 5.6 | [12.0] | 4.3 | [18.2] | 3.4 | [9.0] | 1.5 | [8.0] |
| N(OP) | 12.8 | [16.0] | 14 | [15.2] | 17.1 | [17.3] | 13.8 | [18.2] | 13.5 | [14.9] | 8.3 | [13.3] |
| N(GP) | 17.2 | [18.0] | 15.5 | [13.9] | 17.1 | [15.2] | 19.1 | [19.0] | 17.7 | [14.6] | 16.9 | [17.0] |
| N(CALIBER) | 18.3 | [8.5] | 14.7 | [7.4] | 18.6 | [8.1] | 22.5 | [8.3] | 20.4 | [7.4] | 20.8 | [7.3] |
| N(LTC) | 14.5 | [7.3] | 12.2 | [6.4] | 14.8 | [6.9] | 18.3 | [7.1] | 16.6 | [6.4] | 16.6 | [6.4] |
| N(acute) | 3.7 | [2.8] | 2.5 | [2.5] | 3.9 | [2.8] | 4.2 | [2.8] | 3.8 | [2.6] | 4.2 | [2.5] |
| Disease system | | | | | | | | | | | | |
| Cancers | 1,359 | [17.0%] | 1,784 | [22.5%] | 4,962 | [59.8%] | 1,167 | [16.2%] | 6,038 | [60.5%] | 2,541 | [23.6%] |
| Circulatory | 5,560 | [69.4%] | 5,853 | [73.9%] | 6,442 | [77.6%] | 6,825 | [94.8%] | 9,114 | [91.3%] | 10,299 | [95.7%] |
| Digestive | 5,958 | [74.3%] | 4,740 | [59.9%] | 5,295 | [63.8%] | 4,821 | [67.0%] | 6,874 | [68.9%] | 6,585 | [61.2%] |
| Ear conditions | 1,331 | [16.6%] | 1,577 | [19.9%] | 1,963 | [23.6%] | 1,728 | [24.0%] | 3,247 | [32.5%] | 3,627 | [33.7%] |
| Endocrine | 4,881 | [60.9%] | 4,691 | [59.3%] | 4,833 | [58.2%] | 5,752 | [79.9%] | 6,005 | [60.2%] | 6,787 | [63.1%] |
| Eye conditions | 2,079 | [25.9%] | 2,196 | [27.7%] | 3,223 | [38.8%] | 3,983 | [55.4%] | 4,550 | [45.6%] | 6,206 | [57.7%] |
| Genitourinary | 4,832 | [60.3%] | 3,950 | [49.9%] | 5,492 | [66.1%] | 5,606 | [77.9%] | 6,976 | [69.9%] | 8,598 | [79.9%] |
| Respiratory | 4,736 | [59.1%] | 3,739 | [47.2%] | 4,570 | [55.0%] | 4,941 | [68.7%] | 6,039 | [60.5%] | 6,234 | [57.9%] |
| Haematological/Immunological | 3,978 | [49.6%] | 3,055 | [38.6%] | 4,315 | [52.0%] | 4,320 | [60.0%] | 5,075 | [50.8%] | 5,550 | [51.6%] |
| Infectious Disease | 5,391 | [67.3%] | 4,088 | [51.6%] | 5,581 | [67.2%] | 5,526 | [76.8%] | 7,149 | [71.6%] | 8,521 | [79.2%] |
| Mental Health | 6,042 | [75.4%] | 3,927 | [49.6%] | 4,415 | [53.2%] | 4,142 | [57.6%] | 5,406 | [54.2%] | 7,208 | [67.0%] |
| Musculoskeletal | 4,663 | [58.2%] | 5,742 | [72.5%] | 5,840 | [70.3%] | 5,190 | [72.1%] | 7,655 | [76.7%] | 8,577 | [79.7%] |
| Neurological | 2,851 | [35.6%] | 1,734 | [21.9%] | 1,993 | [24.0%] | 2,328 | [32.4%] | 2,110 | [21.1%] | 2,530 | [23.5%] |

| | | | | | | | | | | | | |
|---|---|---|---|---|---|---|---|---|---|---|---|---|
| Skin conditions | 2,865 | [35.7%] | 2,779 | [35.1%] | 2,874 | [34.6%] | 2,128 | [29.6%] | 2,944 | [29.5%] | 2,868 | [26.6%] |
| In-year Mortality | 998 | [12.5%] | 853 | [10.8%] | 1,892 | [22.8%] | 1,452 | [20.2%] | 2,859 | [28.6%] | 2,705 | [25.1%] |

<u>Relative Prevalence for Top 20 Diseases</u>

| (1) | | (2) | | (3) | |
|---|---|---|---|---|---|
| Alcoholic liver disease | 5.6 | Irritable bowel syndrome | 1.1 | Secondary malignancy_lymph nodes | 2.5 |
| Portal hypertension | 5.3 | Spondylosis | 1.1 | Hypersplenism | 1.4 |
| Acne | 4.9 | Secondary thrombocytopaenia | 1.1 | Primary malignancy_skin | 1.3 |
| Personality disorders | 4.9 | Osteoarthritis (excl spine) | 1.1 | Secondary thrombocytopaenia | 1.2 |
| Cirrhosis | 4.6 | Obesity | 1.0 | Irritable bowel syndrome | 1.2 |
| Bipolar affective disorder and mania | 4.4 | Gastro-oesophageal reflux disease | 1.0 | Infections of the digestive system | 1.1 |
| Substance misuse | 3.5 | Anxiety disorders | 1.0 | Chronic fatigue syndrome | 1.0 |
| Schizophrenia | 3.4 | Depression | 0.9 | Spondylosis | 1.0 |
| Chronic fatigue syndrome | 3.4 | Asthma | 0.9 | Osteoporosis | 1.0 |
| Hypersplenism | 3.3 | Oesophagitis and oesophageal ulcer | 0.9 | Gastro-oesophageal reflux disease | 1.0 |
| Pancreatitis | 3.1 | Alcohol misuse | 0.9 | Anxiety disorders | 1.0 |
| Fatty Liver | 2.6 | Diverticular disease of intestine | 0.9 | Acne | 1.0 |
| Alcohol misuse | 2.3 | Peripheral neuropathy | 0.9 | Cholelithiasis | 1.0 |
| Epilepsy | 2.1 | Hypertension | 0.8 | Thyroid disease | 1.0 |
| Cholelithiasis | 2.0 | Thyroid disease | 0.8 | Other anaemias | 1.0 |
| Secondary thrombocytopaenia | 1.7 | Diaphragmatic hernia | 0.8 | Infections of other or unspecified organs | 1.0 |
| Depression | 1.6 | Infections of the digestive system | 0.8 | Pleural effusion | 1.0 |
| Irritable bowel syndrome | 1.6 | Hyperplasia of prostate | 0.8 | Glaucoma | 1.0 |
| Anxiety disorders | 1.6 | Primary malignancy_skin | 0.8 | Diverticular disease of intestine | 1.0 |
| Oesophagitis and oesophageal ulcer | 1.5 | Gout | 0.8 | Other or unspecified infectious organisms | 0.9 |
| (4) | | (5) | | (6) | |
| Rheumatic valve disorder | 3.6 | Primary malignancy_prostate | 3.2 | Dementia | 2.2 |
| Diabetic neuropathy | 3.6 | Secondary malignancy_lymph nodes | 1.9 | Left bundle branch block | 2.0 |
| Secondary pulmonary hypertension | 3.0 | Hyperplasia of prostate | 1.6 | Macular degeneration | 1.8 |
| Other cardiomyopathy | 2.8 | Diverticular disease of intestine | 1.3 | Ischaemic stroke | 1.8 |
| Sleep apnoea | 2.4 | COPD | 1.3 | Atrioventricular blocks | 1.8 |
| Peptic ulcer disease | 2.2 | Diaphragmatic hernia | 1.2 | Delirium, not induced by alcohol and other psychoactive substances | 1.8 |
| Diabetic eye disease | 2.2 | Primary malignancy_skin | 1.2 | Stroke NOS | 1.7 |
| Nonrheumatic mitral valve disorders | 1.9 | Oesophagitis and oesophageal ulcer | 1.2 | Transient ischaemic attack | 1.7 |
| Multiple valve disorder | 1.9 | Hearing loss | 1.2 | Nonrheumatic aortic valve disorders | 1.6 |
| Peripheral neuropathy | 1.8 | Gout | 1.2 | Multiple valve disorder | 1.6 |
| Heart failure | 1.8 | Stroke NOS | 1.2 | Visual impairment and blindness | 1.6 |
| Peripheral arterial disease | 1.7 | Atrial fibrillation | 1.1 | Right bundle branch block combinations | 1.6 |
| Unstable angina | 1.7 | Macular degeneration | 1.1 | Urinary tract infections | 1.5 |
| Diabetes | 1.7 | Lower respiratory tract infections | 1.1 | Atrial fibrillation | 1.5 |
| Myocardial infarction | 1.6 | Cataract | 1.1 | Osteoporosis | 1.5 |
| Respiratory failure | 1.6 | Transient ischaemic attack | 1.1 | Heart failure | 1.5 |
| Coronary heart disease NOS | 1.5 | Glaucoma | 1.1 | Myocardial infarction | 1.5 |
| Infection of skin and subcutaneous tissues | 1.5 | Ischaemic stroke | 1.1 | Unstable angina | 1.5 |
| Right bundle branch block combinations | 1.5 | Pleural effusion | 1.1 | Nonrheumatic mitral valve disorders | 1.5 |
| Stable angina | 1.5 | Hypertension | 1.1 | Cataract | 1.5 |

b. Fuzzy k-medoids, k=7

| | (1) Liver disease & mental health N=7,644 (14.7%) | | (2) Low multimorbidity N=9,399 (18.0%) | | (3) Chronic fatigue, immunological and haematological disorders N=5,723 (11.0%) | | (4) Nodal metastases N=7,044 (13.5%) | | (5) Cardiometabolic disease N=7,779 (14.9%) | | (6) Prostate cancer (with nodal metastasis) N=6,202 (11.9%) | | (7) Dementia, cardiovascular disease, and sensory impairment N=8,384 (16.1%) | |
|---|---|---|---|---|---|---|---|---|---|---|---|---|---|---|
| Age | 56.3 | [18.1] | 66.1 | [15.9] | 66.9 | [18.6] | 69.2 | [13.6] | 72.3 | [15.0] | 77.2 | [11.4] | 82.4 | [11.2] |
| Female | 3,790 | [49.6%] | 4,319 | [46.0%] | 3,599 | [62.9%] | 3,798 | [53.9%] | 3,587 | [46.1%] | 1,746 | [28.2%] | 4,507 | [53.8%] |
| Ethnicity | | | | | | | | | | | | | | |
| White | 6,415 | [83.9%] | 9,140 | [97.2%] | 4,054 | [70.8%] | 6,504 | [92.3%] | 5,830 | [74.9%] | 5,818 | [93.8%] | 7,619 | [90.9%] |
| Mixed | 0 | [0.0%] | 0 | [0.0%] | 1 | [0.0%] | 0 | [0.0%] | 240 | [3.1%] | 1 | [0.0%] | 0 | [0.0%] |
| Asian | 387 | [5.1%] | 154 | [1.6%] | 219 | [3.8%] | 245 | [3.5%] | 1,182 | [15.2%] | 145 | [2.3%] | 132 | [1.6%] |
| Black | 661 | [8.6%] | 99 | [1.1%] | 230 | [4.0%] | 175 | [2.5%] | 430 | [5.5%] | 183 | [3.0%] | 93 | [1.1%] |
| Other | 3 | [0.0%] | 0 | [0.0%] | 12 | [0.2%] | 0 | [0.0%] | 0 | [0.0%] | 0 | [0.0%] | 520 | [6.2%] |
| unknown | 178 | [2.3%] | 6 | [0.1%] | 1,207 | [21.1%] | 120 | [1.7%] | 97 | [1.2%] | 55 | [0.9%] | 20 | [0.2%] |
| IMD quintiles | | | | | | | | | | | | | | |
| 1 (least deprived) | 1,069 | [14.0%] | 1,938 | [20.6%] | 1,132 | [19.8%] | 1,449 | [20.6%] | 1,169 | [15.0%] | 1,356 | [21.9%] | 1,775 | [21.2%] |
| 2 | 1,176 | [15.4%] | 1,983 | [21.1%] | 1,092 | [19.1%] | 1,510 | [21.4%] | 1,394 | [17.9%] | 1,283 | [20.7%] | 1,842 | [22.0%] |
| 3 | 1,372 | [17.9%] | 1,950 | [20.7%] | 1,109 | [19.4%] | 1,435 | [20.4%] | 1,519 | [19.5%] | 1,206 | [19.4%] | 1,610 | [19.2%] |
| 4 | 1,811 | [23.7%] | 1,750 | [18.6%] | 1,198 | [20.9%] | 1,322 | [18.8%] | 1,782 | [22.9%] | 1,170 | [18.9%] | 1,599 | [19.1%] |
| 5 (most deprived) | 2,216 | [29.0%] | 1,778 | [18.9%] | 1,192 | [20.8%] | 1,328 | [18.9%] | 1,915 | [24.6%] | 1,108 | [17.9%] | 1,558 | [18.6%] |
| unknown | 0 | [0.0%] | 0 | [0.0%] | 0 | [0.0%] | 0 | [0.0%] | 0 | [0.0%] | 79 | [1.3%] | 0 | [0.0%] |
| Usage in 2018/19 | | | | | | | | | | | | | | |
| Total care costs (£) | 18,280 | [10,176] | 16,750 | [9,208] | 18,132 | [10,138] | 16,685 | [7,479] | 18,471 | [9,366] | 17,275 | [7,938] | 17,069 | [7,702] |
| Planned care (£) | 5,939 | [8,230] | 8,074 | [9,059] | 7,386 | [9,493] | 7,443 | [7,170] | 5,333 | [7,481] | 5,560 | [6,846] | 3,752 | [5,987] |
| Unplanned care (£) | 12,326 | [9,520] | 8,684 | [8,042] | 10,761 | [8,454] | 9,241 | [7,524] | 13,149 | [8,964] | 11,708 | [8,103] | 13,313 | [7,623] |
| Secondary care (£) | 17,766 | [10,176] | 16,311 | [9,222] | 17,590 | [10,165] | 16,168 | [7,465] | 17,873 | [9,349] | 16,688 | [7,941] | 16,461 | [7,682] |
| Primary care (£) | 514 | [539] | 439 | [434] | 542 | [505] | 517 | [457] | 598 | [561] | 587 | [538] | 608 | [599] |
| N(EM) | 3.7 | [3.8] | 2.1 | [2.2] | 3 | [2.6] | 3 | [2.2] | 3.2 | [2.7] | 2.7 | [2.1] | 3 | [2.1] |
| N(ED) | 7 | [14.9] | 2.8 | [5.2] | 4.2 | [7.8] | 2.9 | [3.5] | 4.1 | [6.2] | 3.4 | [3.7] | 3.9 | [4.0] |
| N(EL) | 3.7 | [12.7] | 3.8 | [9.5] | 4.9 | [12.9] | 5.5 | [9.8] | 3.8 | [16.9] | 2.6 | [8.6] | 1.5 | [8.2] |
| N(OP) | 12.6 | [15.9] | 13.2 | [14.9] | 15 | [15.9] | 18 | [17.1] | 13 | [18.0] | 11.9 | [14.1] | 8.3 | [13.7] |
| N(GP) | 17.4 | [18.3] | 15.2 | [13.8] | 18 | [15.5] | 17 | [14.0] | 19.4 | [18.7] | 18.2 | [15.2] | 16.8 | [17.6] |
| N(CALIBER) | 18.4 | [8.4] | 14.8 | [7.1] | 19.3 | [8.3] | 18.2 | [7.4] | 23.2 | [8.0] | 21.3 | [7.5] | 21 | [7.2] |
| N(LTC) | 14.7 | [7.2] | 12.2 | [6.2] | 15 | [7.1] | 15 | [6.3] | 18.8 | [6.9] | 17.4 | [6.5] | 16.7 | [6.5] |
| N(acute) | 3.8 | [2.7] | 2.6 | [2.5] | 4 | [2.8] | 3 | [2.5] | 4.4 | [2.7] | 3.8 | [2.6] | 4.4 | [2.5] |
| Disease system | | | | | | | | | | | | | | |
| Cancers | 1296.0 | [17.0%] | 1932.0 | [20.6%] | 2102.0 | [36.7%] | 5716.0 | [81.1%] | 1293.0 | [16.6%] | 3325.0 | [53.6%] | 2187.0 | [26.1%] |
| Circulatory | 5391.0 | [70.5%] | 7214.0 | [76.8%] | 4622.0 | [80.8%] | 5597.0 | [79.5%] | 7475.0 | [96.1%] | 5820.0 | [93.8%] | 7974.0 | [95.1%] |
| Digestive | 5715.0 | [74.8%] | 5590.0 | [59.5%] | 3839.0 | [67.1%] | 4470.0 | [63.5%] | 5187.0 | [66.7%] | 4332.0 | [69.8%] | 5140.0 | [61.3%] |
| Ear conditions | 1271.0 | [16.6%] | 2073.0 | [22.1%] | 1456.0 | [25.4%] | 1578.0 | [22.4%] | 1964.0 | [25.2%] | 2051.0 | [33.1%] | 3080.0 | [36.7%] |
| Endocrine | 4712.0 | [61.6%] | 5607.0 | [59.7%] | 3419.0 | [59.7%] | 4083.0 | [58.0%] | 6161.0 | [79.2%] | 3833.0 | [61.8%] | 5134.0 | [61.2%] |
| Eye conditions | 2,049 | [26.8%] | 2,648 | [28.2%] | 2,481 | [43.4%] | 2,273 | [32.3%] | 4,446 | [57.2%] | 3,123 | [50.4%] | 5,217 | [62.2%] |
| Genitourinary | 4,763 | [62.3%] | 4,768 | [50.7%] | 4,199 | [73.4%] | 3,915 | [55.6%] | 6,199 | [79.7%] | 4,729 | [76.2%] | 6,881 | [82.1%] |
| Respiratory | 4,514 | [59.1%] | 4,678 | [49.8%] | 3,326 | [58.1%] | 3,808 | [54.1%] | 5,498 | [70.7%] | 3,866 | [62.3%] | 4,569 | [54.5%] |
| Haematological/Immunological | 3,856 | [50.4%] | 3,528 | [37.5%] | 3,086 | [53.9%] | 3,347 | [47.5%] | 4,842 | [62.2%] | 3,274 | [52.8%] | 4,360 | [52.0%] |
| Infectious Disease | 5,165 | [67.6%] | 4,983 | [53.0%] | 3,942 | [68.9%] | 4,615 | [65.5%] | 6,234 | [80.1%] | 4,513 | [72.8%] | 6,804 | [81.2%] |
| Mental Health | 5,812 | [76.0%] | 4,736 | [50.4%] | 3,277 | [57.3%] | 3,340 | [47.4%] | 4,571 | [58.8%] | 3,538 | [57.0%] | 5,866 | [70.0%] |
| Musculoskeletal | 4,496 | [58.8%] | 6,532 | [69.5%] | 4,158 | [72.7%] | 4,886 | [69.4%] | 5,736 | [73.7%] | 5,013 | [80.8%] | 6,846 | [81.7%] |

| | | | | | | | | | | | | | | |
|---|---|---|---|---|---|---|---|---|---|---|---|---|---|---|
| Neurological | 2,668 | [34.9%] | 1,934 | [20.6%] | 1,558 | [27.2%] | 1,416 | [20.1%] | 2,404 | [30.9%] | 1,411 | [22.8%] | 2,155 | [25.7%] |
| Skin conditions | 2,736 | [35.8%] | 2,622 | [27.9%] | 2,268 | [39.6%] | 2,334 | [33.1%] | 2,315 | [29.8%] | 1,824 | [29.4%] | 2,359 | [28.1%] |
| In-year Mortality | 965 | [12.6%] | 1,135 | [12.1%] | 973 | [17.0%] | 2,201 | [31.2%] | 1,766 | [22.7%] | 1,622 | [26.2%] | 2,097 | [25.0%] |

Relative Prevalence for Top 20 Diseases

| (1) | | (2) | | (3) | | (4) | |
|---|---|---|---|---|---|---|---|
| Alcoholic liver disease | 5.8 | Peptic ulcer disease | 1.2 | Hypersplenism | 1.8 | Secondary malignancy_lymph nodes | 4.1 |
| Portal hypertension | 5.6 | Oesophagitis and oesophageal ulcer | 1.1 | Chronic fatigue syndrome | 1.7 | Primary malignancy_skin | 1.2 |
| Personality disorders | 5.1 | Obesity | 1.0 | Acne | 1.7 | Diverticular disease of intestine | 1.1 |
| Cirrhosis | 4.8 | Irritable bowel syndrome | 1.0 | Secondary thrombocytopaenia | 1.3 | Pleural effusion | 1.1 |
| Acne | 4.7 | Gastro-oesophageal reflux disease | 1.0 | Irritable bowel syndrome | 1.3 | Irritable bowel syndrome | 1.0 |
| Bipolar affective disorder and mania | 4.6 | Osteoarthritis (excl spine) | 1.0 | Cholelithiasis | 1.2 | COPD | 1.0 |
| Substance misuse | 3.6 | Secondary thrombocytopaenia | 1.0 | Spondylosis | 1.2 | Spondylosis | 1.0 |
| Schizophrenia | 3.5 | Sleep apnoea | 1.0 | Osteoporosis | 1.1 | Infections of the digestive system | 1.0 |
| Hypersplenism | 3.3 | Depression | 0.9 | Infections of the digestive system | 1.1 | Other or unspecified infectious organisms | 1.0 |
| Pancreatitis | 3.2 | Ischaemic stroke | 0.9 | Primary malignancy_skin | 1.1 | Gastro-oesophageal reflux disease | 1.0 |
| Chronic fatigue syndrome | 2.7 | Anxiety disorders | 0.9 | Glaucoma | 1.1 | Obesity | 1.0 |
| Fatty Liver | 2.6 | Alcohol misuse | 0.9 | Visual impairment and blindness | 1.1 | Peripheral neuropathy | 0.9 |
| Alcohol misuse | 2.4 | Spondylosis | 0.9 | Gastro-oesophageal reflux disease | 1.1 | Lower respiratory tract infections | 0.9 |
| Epilepsy | 2.1 | Asthma | 0.9 | Anxiety disorders | 1.1 | Diaphragmatic hernia | 0.9 |
| Cholelithiasis | 2.0 | Stable angina | 0.9 | Iron deficiency anaemia | 1.1 | Oesophagitis and oesophageal ulcer | 0.9 |
| Secondary thrombocytopaenia | 1.7 | Unstable angina | 0.9 | Asthma | 1.0 | Osteoarthritis (excl spine) | 0.9 |
| Irritable bowel syndrome | 1.6 | Hypertension | 0.9 | Infections of other or unspecified organs | 1.0 | Other anaemias | 0.9 |
| Depression | 1.6 | Diverticular disease of intestine | 0.9 | Other anaemias | 1.0 | Peptic ulcer disease | 0.9 |
| Anxiety disorders | 1.6 | Hyperplasia of prostate | 0.8 | Thyroid disease | 1.0 | Iron deficiency anaemia | 0.9 |
| Oesophagitis and oesophageal ulcer | 1.5 | Epilepsy | 0.8 | Bacterial diseases (excl TB) | 1.0 | Thyroid disease | 0.9 |
| **(5)** | | **(6)** | | **(7)** | | | |
| Rheumatic valve disorder | 3.6 | Primary malignancy_prostate | 4.6 | Dementia | 2.6 | | |
| Other cardiomyopathy | 3.6 | Hyperplasia of prostate | 1.9 | Macular degeneration | 2.3 | | |
| Diabetic neuropathy | 3.4 | Stroke NOS | 1.5 | Delirium, not induced by alcohol and other psychoactive substances | 2.0 | | |
| Secondary pulmonary hypertension | 3.4 | Diaphragmatic hernia | 1.4 | Visual impairment and blindness | 2.0 | | |
| Nonrheumatic mitral valve disorders | 2.3 | Ischaemic stroke | 1.4 | Left bundle branch block | 2.0 | | |
| Multiple valve disorder | 2.2 | Primary malignancy_skin | 1.4 | Atrioventricular blocks | 1.9 | | |
| Diabetic eye disease | 2.2 | Gout | 1.3 | Glaucoma | 1.8 | | |
| Heart failure | 2.0 | Transient ischaemic attack | 1.3 | Urinary tract infections | 1.7 | | |
| Sleep apnoea | 1.9 | Hearing loss | 1.3 | Osteoporosis | 1.7 | | |
| Left bundle branch block | 1.8 | Right bundle branch block combinations | 1.3 | Transient ischaemic attack | 1.7 | | |
| Respiratory failure | 1.8 | Urinary tract infections | 1.3 | Ischaemic stroke | 1.6 | | |
| Peripheral arterial disease | 1.8 | Oesophagitis and oesophageal ulcer | 1.3 | Stroke NOS | 1.6 | | |
| Peripheral neuropathy | 1.8 | Atrioventricular blocks | 1.3 | Cataract | 1.6 | | |
| Myocardial infarction | 1.7 | Atrial fibrillation | 1.3 | Right bundle branch block combinations | 1.6 | | |
| Unstable angina | 1.7 | Cataract | 1.3 | Nonrheumatic aortic valve disorders | 1.5 | | |
| Infection of skin and subcutaneous tissues | 1.7 | Diverticular disease of intestine | 1.2 | Hearing loss | 1.5 | | |
| Coronary heart disease NOS | 1.6 | Macular degeneration | 1.2 | Primary malignancy_skin | 1.5 | | |
| Diabetes | 1.6 | Dementia | 1.2 | Atrial fibrillation | 1.5 | | |
| Right bundle branch block combinations | 1.6 | Delirium, not induced by alcohol and other psychoactive substances | 1.2 | Chronic kidney disease | 1.4 | | |
| Atrioventricular blocks | 1.6 | Chronic kidney disease | 1.2 | Multiple valve disorder | 1.4 | | |

### Table S10 – Cross-Sample Comparison of Optimal Clustering

| Random sample 1 (hierarchical, 9 clusters) | Random sample 2 (fuzzy k-medoids, 4 clusters) | Cluster features |
|---|---|---|
| **Liver disease & mental health** | ***Matched*** *(alcoholic liver disease, portal hypertension, personality disorders, cirrhosis, bipolar affective disorder and mania, hypersplenism, substance misuse, schizophrenia, pancreatitis)* | • Mean age: 57.4; most minor ethnicity (non-white: 1,924 [15.1%] of 12,761); living in most derived area (IMD 4 or 5: 6,129 [48.0%])<br>• Highest total care costs (£17,853); Most emergency admissions (3) and ED attendances (5.4) |
| **Sleep apnoea, gastro-reflux and diabetes** | *Dropped, peptic ulcer disease and diabetes embedded within other clusters* | |
| **Chronic fatigue, immunological and haematological disorders** | *Dropped, chronic fatigue embedded within "Nodal metastases and chronic fatigue"* | |
| **Low multimorbidity** | *Dropped* | |
| **Nodal metastases** | ***Reproduced with more chronic fatigue diagnoses "Nodal metastases and chronic fatigue"*** *(nodal metastases, chronic fatigue syndrome, tubule-interstitial nephritis, and irritable bowel syndrome)* | • Mean age: 66.8<br>• Highest planned care costs (£7,516; 5.5 elective admissions; 17.1 outpatient visits)<br>• Highest mortality (2,986 [25.3%] of 11,815) |
| **Complex multimorbidity** | *Dropped, multimorbidity burden splits across "Dementia and cardiovascular disease" and "Prostate and cardiometabolic complications"* | |
| **Prostate cancer (with nodal metastasis)** | ***Reproduced with hyperplasia embedded and more diabetes complications and cardiovascular disease "Prostate and cardiometabolic complications"*** *(diabetic neuropathy, prostate primary malignancy, peripheral arterial disease, peripheral neuropathy, diabetic eye disease, hyperplasia of prostate, tubule-interstitial nephritis, cardiomyopathy, angina, gout, myocardial infarction, CHD).* | • Mean age: 74.6<br>• High total care costs (£17,743); and most GP appointments (19.2)<br>• Most multimorbid (22.4 diagnoses) |
| **Prostate, stroke & sensory impairment** | *Dropped, split to "Prostate and cardiometabolic complications" (prostate component embedded); and "Dementia and cardiovascular disease" (stroke and macular degeneration embedded)* | |
| **Cardiovascular disease & dementia** | ***Reproduced with higher relative prevalence of dementia diagnoses "Dementia and cardiovascular disease"*** *(dementia, stroke, delirium, macular degeneration, rheumatic valve disorder, transient ischaemic attach, left bundle branch block, atrioventricular blocks, secondary pulmonary hypertension, nonrheumatic mitral valve disorders, multiple valve disorder, atrial fibrillation, right bundle branch block combinations, heart failure)* | • Mean age: 79.3; living in least derived area (IMD 4 or 5: 6,007 [38.5%] of 15,584)<br>• Highest unplanned care costs (£12,717) and lowest planned care costs (£4,297) |

**Table S11 – Descriptive statistics of high-cost clusters from 50% random sample 2 (N=52,195; optimal fuzzy k-medoids solution)**

| | (1) Liver disease & mental health | | (2) Nodal metastases and chronic fatigue | | (3) Prostate and cardiometabolic complications | | (4) Dementia and cardiovascular disease | |
|---|---|---|---|---|---|---|---|---|
| | N=12,761 (24.4%) | | N=11,815 (22.6%) | | N=12,035 (23.1%) | | N=15,584 (29.9%) | |
| Age | 57.4 | [17.8] | 66.8 | [16.2] | 74.6 | [12.6] | 79.3 | [11.9] |
| Female | 6,335 | [49.6%] | 6,787 | [57.4%] | 4,000 | [33.2%] | 8,233 | [52.8%] |
| Ethnicity | | | | | | | | |
| White | 10,641 | [83.4%] | 9,210 | [78.0%] | 11,034 | [91.7%] | 14,511 | [93.1%] |
| Mixed | 0 | [0.0%] | 2 | [0.0%] | 1 | [0.0%] | 239 | [1.5%] |
| Asian | 916 | [7.2%] | 414 | [3.5%] | 542 | [4.5%] | 593 | [3.8%] |
| Black | 1,001 | [7.8%] | 381 | [3.2%] | 314 | [2.6%] | 175 | [1.1%] |
| Other | 7 | [0.1%] | 528 | [4.5%] | 0 | [0.0%] | 0 | [0.0%] |
| unknown | 196 | [1.5%] | 1,280 | [10.8%] | 144 | [1.2%] | 66 | [0.4%] |
| IMD quintiles | | | | | | | | |
| 1 (least deprived) | 2,137 | [16.7%] | 2,349 | [19.9%] | 2,301 | [19.1%] | 3,109 | [19.9%] |
| 2 | 2,145 | [16.8%] | 2,288 | [19.4%] | 2,568 | [21.3%] | 3,289 | [21.1%] |
| 3 | 2,350 | [18.4%] | 2,289 | [19.4%] | 2,388 | [19.8%] | 3,179 | [20.4%] |
| 4 | 2,861 | [22.4%] | 2,461 | [20.8%] | 2,366 | [19.7%] | 2,939 | [18.9%] |
| 5 (most deprived) | 3,268 | [25.6%] | 2,428 | [20.6%] | 2,333 | [19.4%] | 3,068 | [19.7%] |
| unknown | 0 | [0.0%] | 0 | [0.0%] | 79 | [0.7%] | 0 | [0.0%] |
| Usage in 2018/19 | | | | | | | | |
| Total care costs (£) | 17,853 | [10,226] | 17,525 | [9,256] | 17,743 | [8,556] | 17,018 | [7,791] |
| Planned care (£) | 7,293 | [9,282] | 7,516 | [8,600] | 6,205 | [7,423] | 4,297 | [6,383] |
| Unplanned care (£) | 10,565 | [9,394] | 10,023 | [8,220] | 11,538 | [8,530] | 12,717 | [7,879] |
| Secondary care (£) | 17,395 | [10,234] | 17,007 | [9,261] | 17,160 | [8,537] | 16,425 | [7,772] |
| Primary care (£) | 458 | [472] | 519 | [492] | 583 | [547] | 593 | [558] |
| N(EM) | 3 | [3.7] | 2.7 | [2.7] | 2.8 | [2.4] | 2.9 | [2.3] |
| N(ED) | 5.4 | [12.7] | 3.6 | [6.6] | 3.7 | [4.6] | 3.8 | [4.7] |
| N(EL) | 4.2 | [12.9] | 5.5 | [12.3] | 3.9 | [14.7] | 1.4 | [7.0] |
| N(OP) | 13.7 | [15.9] | 17.1 | [17.8] | 14.1 | [16.5] | 8.5 | [13.1] |
| N(GP) | 15.8 | [15.4] | 16.9 | [15.1] | 19.2 | [18.7] | 17.2 | [14.9] |
| N(CALIBER) | 15.9 | [8.2] | 18.3 | [7.8] | 22.4 | [7.9] | 20.6 | [7.5] |
| N(LTCs) | 12.7 | [6.9] | 14.6 | [6.7] | 18.5 | [6.8] | 16.5 | [6.4] |
| N(acute) | 3.2 | [2.7] | 3.7 | [2.7] | 3.9 | [2.6] | 4.1 | [2.6] |
| Disease system | | | | | | | | |
| Cancers | 2,326 | [18.2%] | 6,997 | [59.2%] | 5,356 | [44.5%] | 3,374 | [21.7%] |
| Circulatory | 8,511 | [66.7%] | 9,167 | [77.6%] | 11,382 | [94.6%] | 14,976 | [96.1%] |
| Digestive | 8,609 | [67.5%] | 7,379 | [62.5%] | 8,530 | [70.9%] | 9,578 | [61.5%] |
| Ear conditions | 2,309 | [18.1%] | 2,627 | [22.2%] | 3,706 | [30.8%] | 5,044 | [32.4%] |
| Endocrine | 7,395 | [58.0%] | 6,993 | [59.2%] | 8,393 | [69.7%] | 10,103 | [64.8%] |
| Eye conditions | 3,046 | [23.9%] | 4,239 | [35.9%] | 6,607 | [54.9%] | 8,428 | [54.1%] |
| Genitourinary | 6,693 | [52.4%] | 7,402 | [62.6%] | 9,298 | [77.3%] | 11,980 | [76.9%] |
| Respiratory | 6,860 | [53.8%] | 6,632 | [56.1%] | 7,753 | [64.4%] | 9,003 | [57.8%] |
| Haematological/Immunological | 5,755 | [45.1%] | 5,845 | [49.5%] | 6,812 | [56.6%] | 7,969 | [51.1%] |
| Infectious Disease | 7,759 | [60.8%] | 7,954 | [67.3%] | 8,708 | [72.4%] | 11,930 | [76.6%] |
| Mental Health | 8,312 | [65.1%] | 6,111 | [51.7%] | 6,628 | [55.1%] | 10,105 | [64.8%] |
| Musculoskeletal | 7,625 | [59.8%] | 8,143 | [68.9%] | 9,549 | [79.3%] | 12,488 | [80.1%] |
| Neurological | 3,543 | [27.8%] | 2,896 | [24.5%] | 3,536 | [29.4%] | 3,406 | [21.9%] |
| Skin conditions | 4,306 | [33.7%] | 4,220 | [35.7%] | 3,667 | [30.5%] | 4,380 | [28.1%] |
| In-year Mortality | 1,448 | [11.3%] | 2,986 | [25.3%] | 2,757 | [22.9%] | 3,568 | [22.9%] |

Relative Prevalence for Top 20 Diseases

| (1) | | (2) | |
|---|---|---|---|
| Alcoholic liver disease | 3.5 | Secondary malignancy_lymph nodes | 2.6 |
| Portal hypertension | 3.4 | Chronic fatigue syndrome | 2.6 |
| Personality disorders | 3.1 | Tubulo-interstitial nephritis | 1.5 |
| Cirrhosis | 3.0 | Irritable bowel syndrome | 1.2 |
| Bipolar affective disorder and mania | 2.8 | Secondary thrombocytopaenia | 1.1 |
| Hypersplenism | 2.4 | Hypersplenism | 1.0 |
| Substance misuse | 2.3 | Infections of the digestive system | 1.0 |
| Schizophrenia | 2.3 | Gastro-oesophageal reflux disease | 1.0 |
| Pancreatitis | 2.0 | Cholelithiasis | 1.0 |
| Fatty Liver | 1.9 | Pleural effusion | 1.0 |
| Alcohol misuse | 1.8 | Spondylosis | 1.0 |
| Epilepsy | 1.7 | Anxiety disorders | 1.0 |
| Secondary thrombocytopaenia | 1.5 | Other or unspecified infectious organisms | 1.0 |
| Irritable bowel syndrome | 1.4 | Obesity | 1.0 |
| Anxiety disorders | 1.3 | Primary malignancy_skin | 1.0 |
| Depression | 1.3 | Asthma | 1.0 |
| Cholelithiasis | 1.3 | Other anaemias | 0.9 |
| Oesophagitis and oesophageal ulcer | 1.2 | Peripheral neuropathy | 0.9 |
| Peptic ulcer disease | 1.2 | Depression | 0.9 |
| Gastritis and duodenitis | 1.1 | Fatty Liver | 0.9 |
| (3) | | (4) | |
| Diabetic neuropathy | 2.8 | Dementia | 1.9 |
| Primary malignancy_prostate | 2.8 | Ischaemic stroke | 1.7 |
| Peripheral arterial disease | 1.7 | Stroke NOS | 1.6 |
| Peripheral neuropathy | 1.7 | Delirium, not induced by alcohol and other psychoactive substances | 1.6 |
| Diabetic eye disease | 1.6 | Macular degeneration | 1.6 |
| Hyperplasia of prostate | 1.6 | Rheumatic valve disorder | 1.6 |
| Tubulo-interstitial nephritis | 1.5 | Transient ischaemic attack | 1.6 |
| Other cardiomyopathy | 1.5 | Left bundle branch block | 1.6 |
| Unstable angina | 1.5 | Atrioventricular blocks | 1.5 |
| Stable angina | 1.4 | Secondary pulmonary hypertension | 1.5 |
| Gout | 1.4 | Nonrheumatic mitral valve disorders | 1.5 |
| Myocardial infarction | 1.4 | Multiple valve disorder | 1.5 |
| Coronary heart disease NOS | 1.4 | Atrial fibrillation | 1.5 |
| Left bundle branch block | 1.4 | Right bundle branch block combinations | 1.5 |
| Nonrheumatic aortic valve disorders | 1.4 | Heart failure | 1.5 |
| Atrioventricular blocks | 1.4 | Osteoporosis | 1.4 |
| Heart failure | 1.4 | Nonrheumatic aortic valve disorders | 1.4 |
| Secondary pulmonary hypertension | 1.4 | Other cardiomyopathy | 1.4 |
| Primary malignancy_skin | 1.4 | Myocardial infarction | 1.4 |
| Multiple valve disorder | 1.3 | Urinary tract infections | 1.4 |
